# SEEDNet: Covariate-free multi-country settlement-level epidemiological estimates datasets for network analysis

**DOI:** 10.1101/2025.02.26.25322963

**Authors:** Amir Hossein Darooneh, Jean-Luc Kortenaar, Céline M. Goulart, Katie McLaughlin, Sean P. Cornelius, Diego G. Bassani

**Affiliations:** Centre for Global Child Health, The Hospital for Sick Children, Toronto, M5G 0A4, Canada; Department of Physics, University of Zanjan, Zanjan, 45371-38791, Iran; Dalla Lana School of Public Health, University of Toronto, Toronto, M5T 3M7, Canada; Department of Epidemiology, Biostatistics and Occupational Health, McGill University, Montreal, H3A 1Y7, Canada; Department of Physics, Toronto Metropolitan University, Toronto, M5B 2K3, Canada; Department of Paediatrics, University of Toronto, Toronto, M5T 3M7, Canada; ISI Foundation, Turin, 10126, Italy

## Abstract

The study of population health through network science holds high promise, but data sources that allow complete representation of populations are limited in low-and middle-income countries. Large national health surveys designed to gather nationally representative health and development data are promising data sources but are not designed to produce small-area estimates of health indicators. Methods for producing these from national surveys tend to rely on varied covariate data sources and are computationally demanding, limiting their use for network representations of populations. To reduce the sources of measurement error and allow efficient multi-country representation of populations as networks of human settlements here, we present SEEDNet (Settlement-level Epidemiological Estimates Datasets for Network Analysis)^1^, a data library of multi-country representations of population health across human settlements. Our covariate-free method uses georeferenced national surveys to produce SAEs of health indicators through local inverse-distance weighted interpolation and includes an algorithm for the comprehensive identification of population settlements of all sizes across the globe. Our estimates are cross-validated against those obtained using a Bayesian Geostatistical Model. The method is fully automated, requiring a single standard georeferenced survey data source for mapping populations, eliminating the need for indicator or country-specific covariate selection by investigators. Computational efficiency is achieved by restricting computation to human-occupied areas and by adopting a logical aggregation of estimates into the complete range of settlement sizes. Standardized georeferenced settlement-level datasets for 15 indicators and 10 countries were validated in this paper, as well as the novel method to identify settlements. SEEDNet^1^ is a specialized library of nodes that can serve as a basis for network representations of population health in low-and middle-income countries.

## Background & Summary

The study of population health through the network representation of human settlements that are connected and interact requires the availability of high-quality data sources^2^ and complete representation of populations^3^. Especially in low-and middle-income settings, population health data availability is vital in public health decisions^4^ and health surveys are essential tools for gathering information about the health status of populations, monitoring progress in implementing programs and policies, evaluating their impact, and ensuring accountability^5^. These surveys are important potential sources of data for network representations of populations, especially given their public availability, inclusion of geo-referenced data, and standardization across multiple countries and over time^6^. They have been important sources of information for researchers, healthcare providers, policymakers, and public health officials, receiving large investments of time and resources over decades^7^ and being conducted in regular intervals in most low-and middle-income countries.

The results produced by surveys are usually representative of large geographical areas such as states/provinces and regions^6^. Probabilistic sampling proportional to the population of the sampling frame area results in sparse coverage for small-area inferences and corresponding data gaps for some areas of each country^8^. Estimates for smaller areas are often desired for decision-making^9,10^, and would be necessary for the study of population health through network representations. Multiple methods have been proposed to generate more fine-grained estimates within the survey design and sample size constraints^11^, but post-hoc disaggregation of data invariably further reduces the precision of the estimates. These losses of inferential power are especially taxing on indicators measured within narrow age groups, within short time windows, or other selected sub-samples of respondents, such as those who experienced a specific outcome during the weeks or months preceding data collection^11–13^. Approaches to circumvent some of these issues and achieve reliable small-area estimates are primarily based on the assumption that the value of a health indicator for any point in time and space can be predicted by a combination of time and space-varying covariates^8^, as long as these are available at the desired resolution^11^. Covariates for the area-level predictive models are identified from datasets with high space and time granularity^14,15^ and interpolation is used to smooth estimates, with varying degrees of computational complexity^8,11–13^.

While it may be argued that these methods provide statistically acceptable predictions of the value of health indicators in a country, their computation requires input from a large number of covariates from heterogeneous sources^16,17^, each of which is measured with varying uncertainty. In addition, separate regression-based covariate selection exercises must be conducted for each country, time-point, and indicator, often resulting in different prediction models for each combination of these and often requiring investigator input throughout^14,15,18–20^. To improve the precision of the estimates across non-sampled areas of the grid, census data and spatial random effects are incorporated, sometimes reducing comparability across settings. Data quality and comparability is also impacted by the design of the data collection or sensing methodology, the processing of measurement error estimates, methodological choices of the upstream geospatial estimation techniques used, as well as variability in time and space resolution. All of these strategies inject a high degree of uncertainty into the resulting estimates, some of which are impossible to ascertain given multiple and variable potential sources. The use of diverse input data sources and methods for producing network representations of populations impacts node attributes^2^ but are difficult to account for, affecting the comparability of representations across countries and time. In addition, the computational time required to produce estimates using these methods is substantial^21^. This is in part because the estimation of high-resolution maps of health indicators typically is conducted for all areas of the grid, even though many areas are not occupied by human settlements^22^. Therefore, even though complete network representations of populations require defining the areas (nodes) for which demographic and health indicators (node attributes) would be estimated, producing high-resolution maps of health indicators for unoccupied areas of the grid is not necessary.

To overcome existing methods’ limitations and efficiently produce meaningful representations of health indicators across countries we propose an alternative approach that (i) adopts the population settlement (a community of people living in a place) as a standardized real-world minimal aggregation unit (node) and (ii) reduces sources of variation through covariate-free estimation. The method enables the estimation of small-area population health indicators for multiple countries using empirical standardized survey data as inputs and is particularly well-suited for automated tasks. Its technical validation and a library of maps of indicators across the full range of identifiable human settlements for multiple countries are presented here. These representations comprehensively expose the underlying complex network structure of populations, enabling their study through network science approaches. Our multi-country library of georeferenced nodes (settlements) is the first to capture the full scale of settlement sizes in low-and middle-income countries and serves as the foundation for developing more elaborate real-world network representations of populations.

## Methods

### Data Sources

#### Health Indicators

The Demographic and Health Surveys (DHS) program implements standard health surveys across multiple low-and middle-income countries^23^. The DHS are cross-sectional, standardized, and nationally representative surveys that use multi-stage probabilistic sampling of individuals residing in sampled households (secondary sampling units) from selected areas (primary sampling units), hereafter referred to as *survey clusters*^24^. The publicly available spatial datasets for Angola-2015^25^, Benin-2017^26^, Cambodia-2014^27^, Gabon-2012^28^, Malawi-2015^29^, Mali-2018^30^, Mozambique-2011^31^, Nigeria-2013^32^, Senegal-2019^33^ and Zambia-2018^34^ were used in the technical validation. The DHS uses a two-stage stratified cluster-randomized design, typically stratified at the regional level and by rural/urban populations, as defined by each country. In the first stage, census-based enumeration areas (*survey clusters*) are sampled with probability proportional to the *survey cluster* size (number of households). In the second stage, a predetermined number of households are systematically selected from each sampled *survey clusters*. All eligible women 15-49 years of age (with some variability across surveys^35^) in the selected households are interviewed. The DHS questionnaires cover various topics, but our validation focuses primarily on selected indicators covering the maternal and child health continuum that are routinely collected across most countries, as well as structural indicators capturing information about infrastructure (Access to Electricity and to Improved Water Sources). Geographic information such as longitude and latitude for each *survey cluster* is recorded; however, to safeguard the privacy of respondents, each pair of geographical coordinates made available for each cluster is randomly displaced^36,37^. The geographic coordinates for selected clusters may be removed in special situations, such as during armed conflicts. The procedures and questionnaires for standard DHS surveys are reviewed and approved by the ICF Institutional Review Board (IRB) and country-specific DHS survey protocols are reviewed by the ICF IRB and by the IRB in the host country. The surveys comply with the U.S. Department of Health and Human Services regulations for the protection of human subjects (45 CFR 46) and reviews by the host country IRB ensures compliance with local laws and norms. The study is also registered under the number 1000081879 at the Research and Ethics Board of the The Hospital for Sick Children.

Datasets^38^ used in this work were downloaded from the DHS Program through their API^35^. The data file of each country includes several standard health indicators collected at the household or individual level through questionnaires. Household and individual-level data from these files were pooled to obtain estimates of health indicators at the cluster level. Survey weights that account for the survey design and sampling probabilities were used to estimate the indicator values at the different administrative levels of the country during the validation steps. *Survey cluster* with missing geographical coordinates were excluded from these analyses. This issue affected six of the ten countries assessed, and within these, it was limited to less than 5% of the *survey clusters* (Gabon-2012, 1.2%; Zambia-2018, 1.8%; Nigeria-2013, 0.8%; Mali-2018, 4.9%; Benin-2017, 2.7%; Mozambique-2011, 0.2%). Angola-2015, Malawi-2015, Cambodia-2014, and Senegal-2019 had no clusters excluded.

Due to the computational time required for validation, the method was developed and validated on 10 DHS surveys and 15 indicators. Nevertheless, the methods were developed to be scaled, prioritizing simplified and standardized data input requirements and the use of open-source datasets. They do not require country-specific model fitting, allowing users to easily expand the pipeline to multiple countries and indicators as these become available. Currently, estimates for 39 countries are available in the SEEDNet library^1^, including the 10 surveys that are part of this technical validation. Some countries have multiple surveys covering multiple years for a total of 54 surveys at the time of submission.

The DHS survey datasets and the corresponding geo-referenced files necessary to reproduce our study from the raw original data are openly available but require user registration for access. Requests to access the data can be made through the DHS program website. At the time of writing this manuscript, access to the remaining input datasets was available without requiring registration.

#### Country and administrative area polygons

The Database of Global Administrative Areas (GADM)^39^ was used as the source for the polygon boundaries for all countries and corresponding levels of sub-national administrative divisions. Datasets are available from GADM in a vector format^40^ and contain the geographic coordinates of the boundary points, which encode one or more closed polygons defining the country. GADM was selected as it provides centralized and standardized Level 2 administrative boundaries for all countries without use restrictions for academic purposes. Although the DHS files include information about the administrative area (region) to which a sampled cluster belongs, these regions do not necessarily correspond to the administrative sub-divisions in the GADM datasets. When such discrepancies were present, information for the integration was obtained from the DHS reports.

#### Settlement identification

The Global Human Settlement (GHS) Model Grid (GHS-SMOD) datasets^41^ for the years 2010 and 2015 with 1 km^2^ resolution were used in this study to identify population settlement boundaries based on the Degree of Urbanization Level 2 classification of grid cells. These datasets^41^ describe human presence worldwide and over time and are available through the *G*lobal Human Settlement Layer (GHSL) project, supported by the European Commission, Joint Research Center, and Directorate-General for Regional and Urban Policy. In our dataset, settlements were defined using the GHS-SMOD R2023A (Settlement model) spatial raster and spatial entities classified by porting the Degree of Urbanization (DEGURBA) model to each of the 1 km^2^ grid cells in order to classify them according to population density, size, and contiguity using the detailed Level 2 classification, resulting in 8 settlement typologies^42^. Details of the spatial data mining techniques used to process the satellite data to produce the built-up maps, population density maps, and settlement maps for multiple times (epochs), resolutions, and coordinate systems are described in detail elsewhere^42,43^. The procedures used in our study to identify settlements were developed to ensure the identification of smaller settlements that would not be classified within rural cluster entities, which are the smallest units with boundaries in the GHSL products. This enabled us to build upon recent developments in this field^44^ and expand the scale of the sizes of human settlements covered by our datasets. The procedures and methods are described in detail below and the accompanying code.

#### Settlement population

To estimate the size of the populations of each settlement, we obtained population count estimates produced by GHSL with a resolution of 30 arc seconds^45^, or 1 km^2^ for the same year the survey was conducted^46^. GHSL produces spatial raster datasets showing the distribution of the residential population per cell between 1975 and 2020 in 5-year intervals and projections to 2025 and 2030. The estimates are based on CIESIN Gridded Population of the World version 4.11 (GPWv4.11)^47^ and are disaggregated into grid cells from census or administrative units. It uses the GHSL global layer for each time-point (epoch) to incorporate the distribution, volume, and classification of built-up areas^48^. In the validation steps of this work, we employed standardized high-resolution raster file^49^ (World Mollweide; EPSG:54009) for 2015—the nearest common year across all 10 surveys—to minimize sources of variability. For the expanded dataset library^1^, the population in the year nearest to each DHS survey^38^ is used.

### Mapping *survey clusters* and creating the DHS spatial sample

A DHS *survey cluster* is a randomly selected area, typically from census records, composed of between 100-300 households and from which a sample of 25 to 30 households are selected to be interviewed by survey data collectors. We assigned a *dominance zone* to each DHS cluster to define the threshold for neighborship. This zone includes all geographical points (1 × 1 km^2^ units from the grid map of the country) closer to its location than to the location of the next nearest neighboring *survey cluster*. This dominance zone definition is based on the generally accepted assumption that the value of a health indicator at any given point in space is more influenced by the values of indicators in nearby locations, with the nearest empirically observed locations (neighboring *survey clusters*) exerting a stronger influence than farthest ones^50^. To delineate the dominance zones using the coordinates of the *survey clusters*, we drew polygonal zones using the DHS cluster coordinates as the seeding points for radial tessellation in the Euclidean plane (Voronoi tessellation). In panel A of Fig. 1, we show a map of Angola with colored zones corresponding to the resulting Voronoi cells. The position of the *survey clusters* is indicated by blue dots at the center of each polygon. The georeferenced *survey clusters* are also used as the representative points of the Voronoi cells. For each pair of neighboring cells, these representative points (centroids) are connected to create a network of *survey clusters* using Delaunay triangulation. The *survey clusters* act as nodes in the network, and links indicate adjacent cells. This DHS cluster network is spatially embedded, as the location of nodes is also significant beyond their topology (the links). In panel B of Fig. 1, we plot the georeferenced *survey clusters* for the DHS survey conducted in 2015 in Angola.

**Figure 1.**
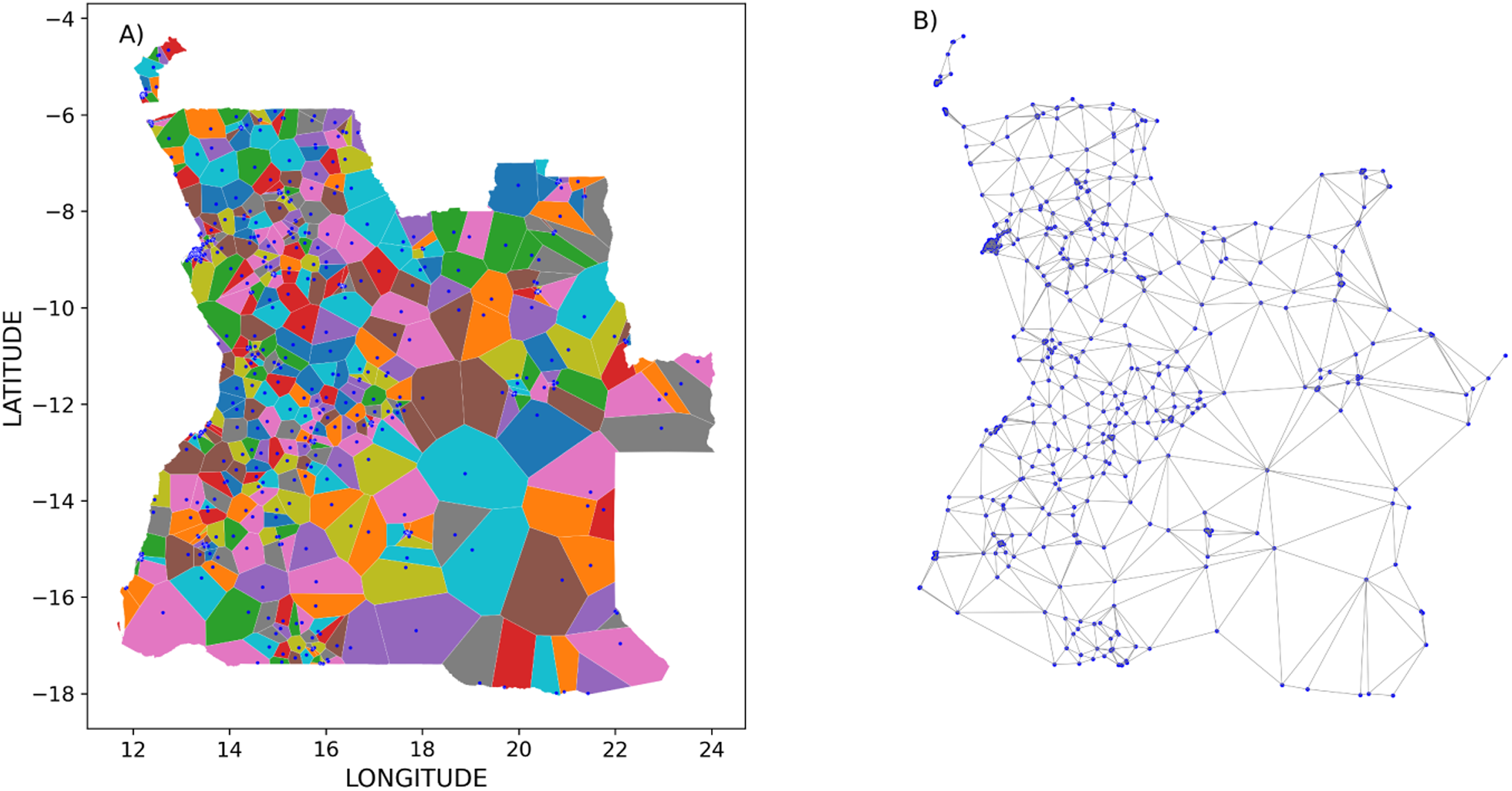
A) Panel A displays the Voronoi cells for Angola in 2015, with blue points indicating the locations of *survey clusters*. Each polygon is uniquely colored for ease of distinction. B) The corresponding spatial distribution of the *survey clusters* included in the Angola-2015 survey can be seen in Panel B.

### Local Inverse Distance Weighting Interpolation

We used Local Inverse Distance Weighting Interpolation to produce high-resolution base maps of occupied areas (settlements) of health indicators based on DHS survey data. Health indicators often exhibit spatial correlations, and proximity, shared resources, and community characteristics contribute to these dependencies. Assuming spatial correlation^51^, interpolation methods are used for predicting health indicators at unknown points based on known values from scattered data. These methods take into account the spatial relationships between areas and can provide insights into the health status of the population in areas for which direct observation is not available. Inverse distance weighting (IDW) interpolation is a method used in geographic information systems (GIS) and other fields to estimate unknown values at a particular location based on values from surrounding known points^50,52^. In IDW interpolation, the estimated value at a given location is calculated as a weighted average of the known values, where the weights are inversely proportional to the distances between the interpolated (unknown) point and the interpolating (known) points. The closer the known points are to the unknown point, the more weight they carry in the interpolation. IDW interpolation is used for a variety of applications, such as creating digital elevation models, predicting air quality, and estimating soil properties^53–56^. This is based on the first law of geography^57^ and computationally simple while also agnostic to the underlying data distribution. Importantly, it assumes a smooth surface between known points and is sensitive to the selection of weighting parameters, all of which can affect the accuracy of the interpolation results^58^. The local aspect of IDW (LIDW) refers to the fact that the weighting is only applied to a subset of the nearest known points rather than all the known points in the dataset^52,58^. Typically, determining a threshold value for assessing neighborship is necessary. However, we adopt a parameter-free approach by utilizing Voronoi tessellation. This allows for greater spatial resolution in the interpolation results, as distant points have less influence on the estimated values. The equation can be written as:

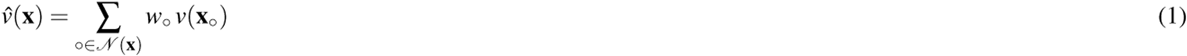

Where 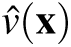 is the estimated value for the health indicator at point **x** and *v*(**x**_◦_) is the known value of the health indicator at the neighboring cluster 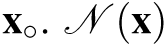 represents the set of neighboring cluster points for the point **x**. The weight *w*_◦_ is proportional to the inverse distance of the point **x** to the cluster location **x**_◦_, and can be written as:

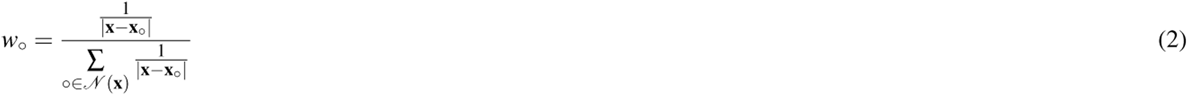

here |**x** − **x**_◦_| stands for geographical (Euclidean) distance between **x** and **x**_◦_.

Figure 2 illustrates one step of the estimation for one grid location **x** (black square in the example), where the neighbors of location **x** in a given Voronoi cell include the representative point (empirically observed in the DHS survey) of its own Voronoi cell (marked with a hollow circle within the central cell), as well as the representative points of adjacent cells (hollow circles within the adjacent cells). All empirically observed points have fixed positions. The adjacency between cells is determined using the DHS cluster sample and is present when there is a direct link between two nodes. The linear distances between the interpolated pixel and interpolating point are used to calculate the weights used in each interpolation.

**Figure 2.**
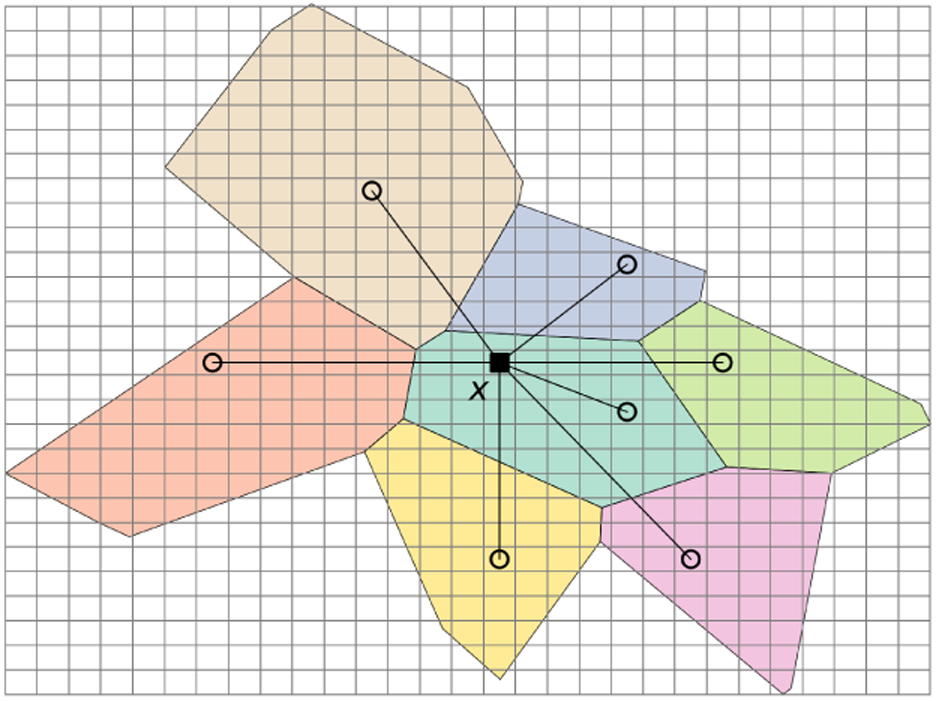
Local inverse distance weighting interpolation (LIDW): the value of an indicator in *x* is calculated based on the known values of the indicator at the *survey clusters* in its neighboring cells (represented by the hollow circles in the neighboring cells). The neighboring Voronoi cells are colored solely for ease of identification. Choosing a resolution is necessary to produce a high-resolution image in practice.

In practice, we divide a country map into pixels of a given resolution (1 km^2^), using the high-resolution global gridded data produced by GHSL^42^. As the value of indicators at the pixel that contains the location of a DHS cluster is known, and because this point is the representative point of a Voronoi cell, we can allocate all pixels with known values (from the DHS survey) to a cell. The value of each health indicator is estimated for all remaining unknown pixels within the settlement polygon (i.e. within the boundaries of a settlement polygon, identified as described in detail below). The neighboring unknown pixels of any pixel contain information about the originating point of its own Voronoi cell as well as from the originating points of neighbouring cells. Figure 3 shows the high-resolution image for the prevalence of underweight children in Angola in 2015 produced using this method. As described above, estimates are only produced for areas occupied by populations.

**Figure 3.**
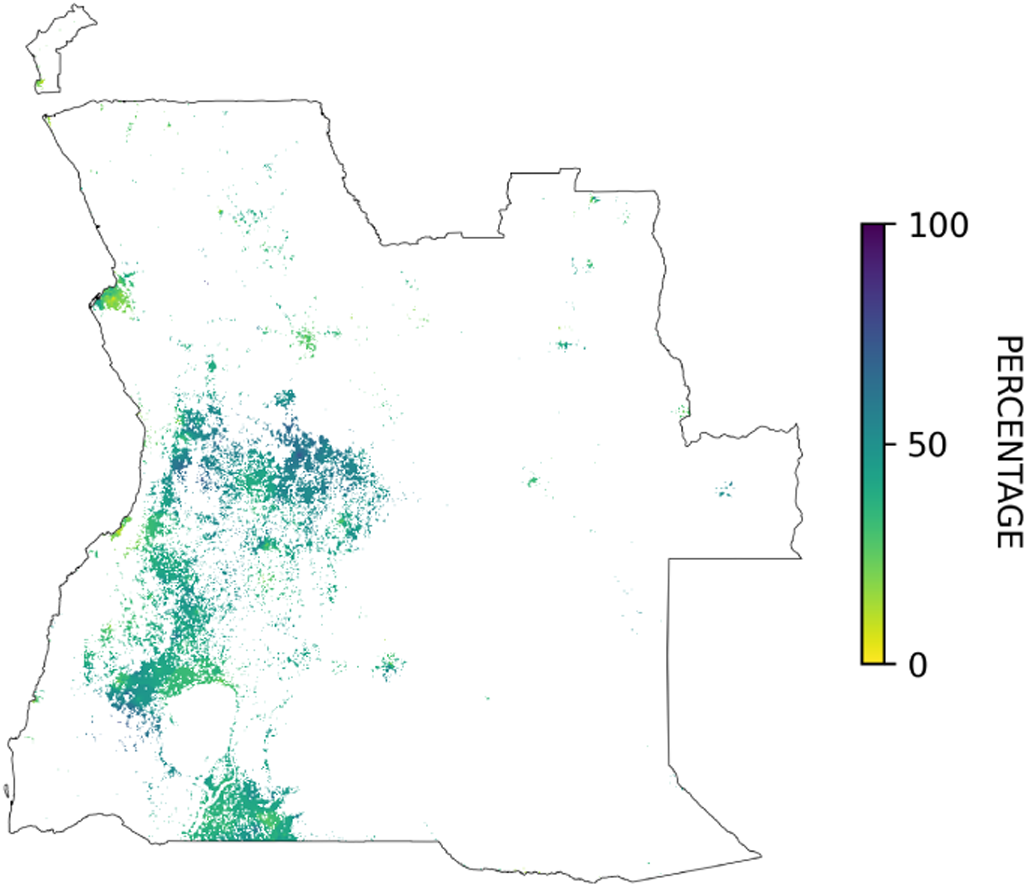
High-resolution image for the percentage of stunted children (<-2 SD height-for-age z-scores) in Angola in 2015 produced using LIDW interpolation based on the *survey clusters*.

### Identifying Population Settlements

The GHSL-SMOD dataset contains high-resolution information about human settlements worldwide (gridded). It relies on heterogeneous data, including streams of fine-scale satellite imagery, census data, and crowd-sourced or volunteered geographic information sources. GHSL has recently implemented a new spatial data mining method to produce the final raster grid of human presence on the planet^42^. One of the classes (city, dense town, suburb, village, dispersed rural area, uninhabited area, and water-covered surface) is assigned to each pixel in the earth map^59^. This dataset is publicly available at 1 km^2^ resolution using the Mollweide coordinate reference system. In our study, the geographic location data was re-projected into the World Geodetic System (WGS84) to obtain latitude and longitude information compatible with those used in the DHS surveys.

To identify each country’s settlements, the world’s raster file was cropped using the polygon that defines the country borders (defined by GADM^40^). Identifying the boundaries of human settlements remains a complex problem with no accepted standard solution. In this paper, to identify the boundaries, we converted the raster file (GHS-SMOD L2 spatial raster dataset) of the country in which pixels are classified using the typologies included in the GHS-SMOD grid to a binary image where each pixel with a value of 1 represents an area classified as inhabited by GHS-SMOD^42^ (codes 12-30) and all other areas as 0. Mostly uninhabited areas (very low-density rural grid cells, code 11 in the GHSL-SMOD) and water-covered areas (water grid cells, code 10 in the GHSL-SMOD) are not added to settlement polygons. The shared edge of a pixel with a value of 1 and a pixel with a value of 0 is considered a piece of the settlement boundary, and the area contained within all contiguous boundaries corresponds to a settlement. This is shown using different colors for the areas near (and including) Luanda, Angola, in Figure 4. This technique works well when defining settlements for most countries. Still, for example, in contiguously densely populated areas it results in a small number of very large settlements, which in some situations could cover large parts of the country.

**Figure 4.**
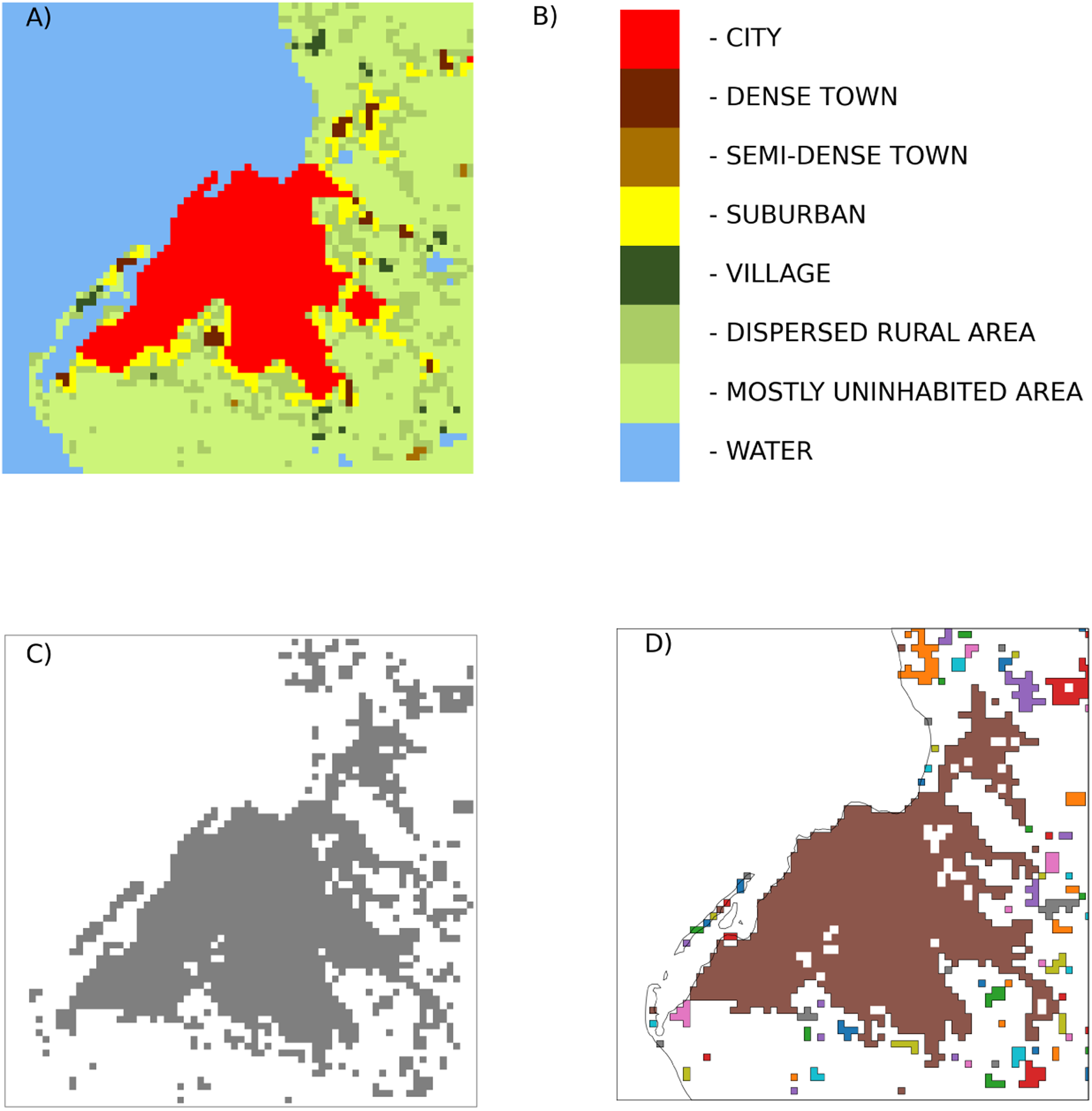
Luanda, Angola, and surrounding settlements in 2015 (the year of the DHS survey). Color is used to distinguish the boundaries of settlements. Panel A shows the GHSL-SMOD L2 spatial raster dataset with the eight classification levels shown in Panel B. Each pixel measures approximately 1 km^2^. Panel C shows the converted raster file used to identify the settlement boundaries. Polygonization was done using the Geospatial Data Abstraction Library. Panel D uses different colors to identify the individual settlements obtained using this approach.

Polygonization was performed using the Geospatial Data Abstraction Library (GDAL), an open-source library for reading and writing raster and vector geospatial data formats^60^. It is widely used in Geographic Information Systems (GIS) and supports various formats and operations. GDAL offers a unified data model for raster and vector data, making it a flexible tool for handling and analyzing geospatial data. The GDAL polygonization tool was used to identify settlement boundaries from binary image files. Supplementary Figure 1 shows the resulting settlement maps for each of the ten countries. We compared the population captured within the settlement polygons to the estimate of the country’s population and observed our settlement polygons captured over 96% of the estimated population of the country (Table 1).

**Table 1.** Validation of the settlement identification method.

| Country-Year | Number of Settlements | Population (000) | Population within Settlements (000) | Population captured (%) | Population not captured (%) |
| --- | --- | --- | --- | --- | --- |
| Angola-2015 | 14,212 | 27,990 | 26,441 | 94.5% | 5.5% |
| Benin-2017 | 3,280 | 10,903 | 10,486 | 96.2% | 3.8% |
| Cambodia-2014 | 3,563 | 15,395 | 15,175 | 98.6% | 1.4% |
| Gabon-2012 | 789 | 1,978 | 1,948 | 98.5% | 1.5% |
| Malawi-2015 | 1,278 | 16,920 | 16,809 | 99.3% | 0.7% |
| Mali-2018 | 14,270 | 18,118 | 16,789 | 92.7% | 7.3% |
| Mozambique-2011 | 13,507 | 26,779 | 24,402 | 91.1% | 8.9% |
| Nigeria-2013 | 32,911 | 183,483 | 179,440 | 97.8% | 2.2% |
| Senegal-2019 | 5,982 | 14,239 | 13,789 | 96.8% | 3.2% |
| Zambia-2018 | 11,643 | 16,278 | 14,499 | 89.1% | 10.9% |
| Technical Validation Sample | 101,435 | 332,086 | 319,779 | 96.3% | 3.7% |

### Estimation of Health Indicators for a Settlement

Two approaches are used to estimate the value of a health indicator for a given settlement. The first approach relies on the indicator values from the high-resolution map, where the values of the pixels within each settlement are estimated using the LIDW method described above. This approach is easily implemented, using the health indicator high-resolution image (Figure 3) and averaging the values of the pixels within the boundaries as shown in the example in Figure 5. The second approach uses information from the clusters sampled in the DHS survey and extrapolates it to settlements that intersect with their displacement buffer circles. Buffer circles for each *survey cluster* are used because the exact longitude and latitude of the cluster are displaced in the pubic datasets to preserve respondent anonymity as described previously. As a result, in our estimation procedures the sampled units (survey clusters) are represented as circles with radii that correspond to the maximum expected displacement, which varies between 2 and 5 km (for urban and rural areas, respectively). A random 1% of the sample is displaced by 10 km but the maximum size of the radii was constrained to 5km in our analysis. Each circle may overlap with the boundaries of one or more settlements. When there are more than one *survey clusters* that overlap with the settlement boundaries, their values are pooled using the DHS weights that account for the multi-stage survey design, probability of selection, and non-response. Estimates produced using the second approach, which is limited to a subset of settlements, are used for validation of the LIDW method described in the section Leave-One-Out Cross-Validation of Out-Of-Sample *predictions* for Settlements.

**Figure 5.**
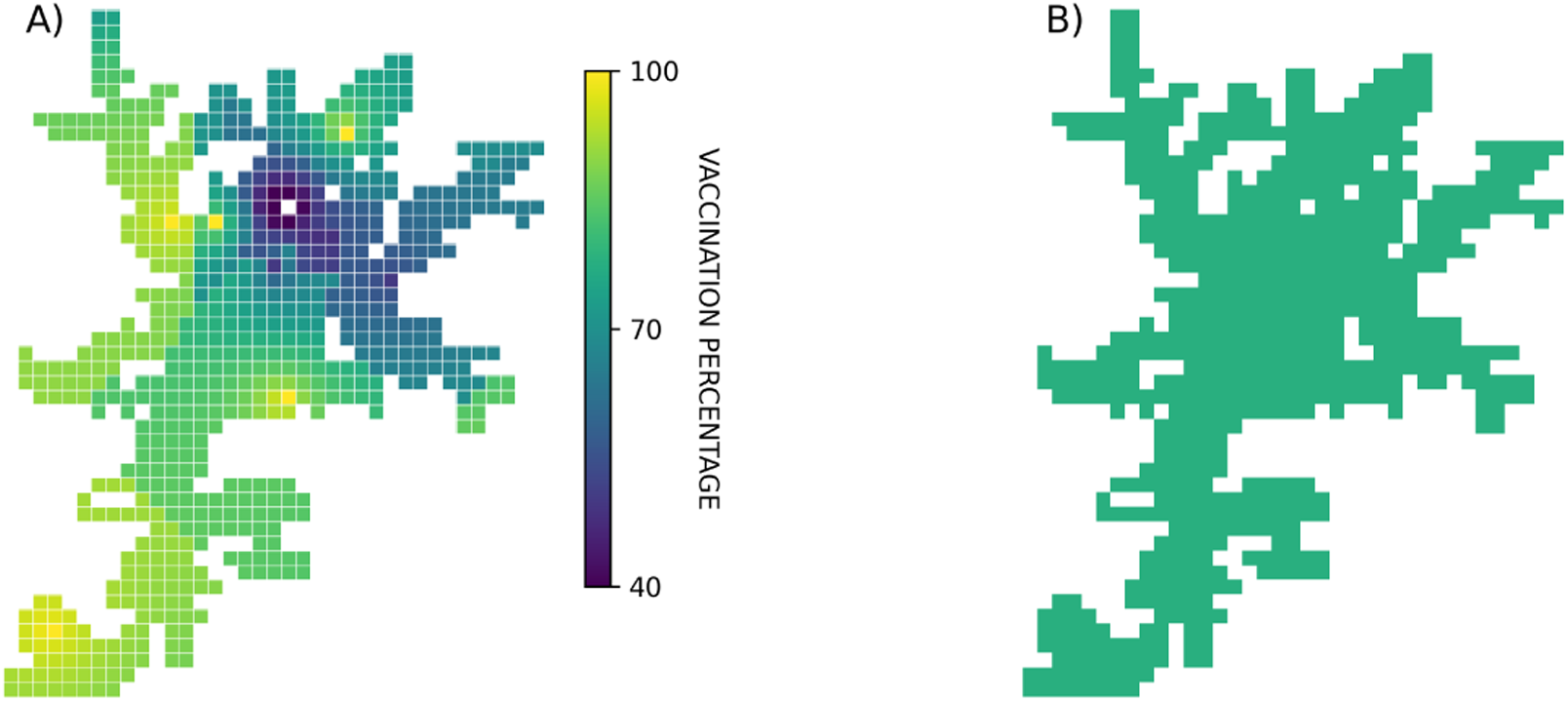
A) High-resolution (each pixel is 1 km^2^) image of vaccination coverage within the borders of one settlement. B) Averaged values within settlement boundaries.

### Validation approaches

During the early stages of method development, we limited the assessments of the validity and reliability of the estimates to measles vaccine coverage among children under five years of age in three countries: Cambodia-2014, Mozambique-2011, and Nigeria-2013. The choice of indicator, definition, age group, and countries intended to match published small-area estimates available from WorldPop. These public datasets are well-described and have been validated^20^. Briefly, WorldPop estimates are based on predictive models that use a set of covariates selected from a pool of 40 candidate covariates from nearly 20 different data sources. Covariates are identified using non-spatial binomial Generalized Linear Models (GLM) fit separately for each country and four age groups (0-8, 9-11, 12-23, and 24-59 months) and for each indicator (in this case, measles immunization)^20^. Covariates are georeferenced and include measures of urbanization, remoteness, poverty, and population density, among others, which vary by indicator being estimated. The small-area estimates are based on these external (i.e. non-DHS) covariates, predicted through a Bayesian Geostatistical Model (BGM) implemented via Markov Chain Monte Carlo (MCMC) methods. Resulting maps and details of the BGM methodology can be found elsewhere^20^. Our preliminary objective was to assess the feasibility of producing comparable aggregated estimates without relying on external covariates and with minimum input data sources.

### LIDW Leave-One-Out Cross-Validation using the *survey clusters*

Following this initial assessment, we expanded the sample to include 15 indicators. These are defined in detail in the Supplementary Table 1. The validation sample is composed of 10 DHS surveys: Angola-2015^25^, Benin-2017^26^, Cambodia-2014^27^, Gabon-2012^28^, Malawi-2015^29^, Mali-2018^30^, Mozambique-2011^31^, Nigeria-2013^32^, Senegal-2019^33^ and Zambia-2018^34^.

Interpolation results were assessed using leave-one-out cross-validation. The number of folds equaled the number of data points, in our case, the centroids of the Voronoi cells, corresponding to the *survey clusters*. We iteratively removed one data point from the dataset and utilized the remaining points to predict the value of the removed point, corresponding to a 1 km^2^ cell. This process was repeated for all data points in the dataset. Metrics used to evaluate the similarity between these lists are explained in the Supplementary Material (Statistical Measures of Error).

### Leave-One-Out Cross-Validation of Out-Of-Sample *predictions* for DHS-overlapping Settlements

To assess the quality of the out-of-sample settlement level estimates, we used the subset of settlements with at least one DHS cluster buffer zone overlapping or within its boundaries. For these settlements, which were deemed to have more *direct* estimates given their proximity to a sampled survey unit, after recording the value of their indicators estimated using the full LIDW model, we identified which *survey clusters*’ buffer zones overlapped with the settlement boundaries and removed those *survey clusters* from the sample used for the estimation. After removing these clusters, the remaining clusters were used to produce an out-of-sample estimate of the value of the indicators for each settlements whose original values included *survey clusters* located inside their boundaries or within a maximum of 5km from the settlement, one at a time. The process was repeated by removing one set of clusters corresponding to one settlement from the estimation sample in each round (one per settlement). Full model indicator values (or nearly-directly estimated values of indicators, hereafter referred to as *direct* estimates) were compared to the values obtained using the reduced model (with the *survey clusters* removed). These out-of-sample estimates are hereafter referred to as *predicted* estimates. Values for all pixels within the settlement boundaries were estimated using the LIDW method detailed above, and their values were aggregated, producing a new *predicted* LIDW estimate for that settlement. Results were assessed using Root Mean Square Distance (RMSD), Mean Absolute Difference (MAD), and 95% Prediction Interval Coverage (P95%) as well as the Ratio of *predicted* to *direct* estimates and their difference (*predicted* minus *direct* estimate) by settlement. This cross-validation allowed us to assess the performance of the out-of-sample estimates using a simulation in which the indicator values for the set of settlements (for which the original LIDW estimates include *survey clusters* whose buffer zones are within or overlap with the settlement boundaries) are re-estimated as if that settlement did not have co-incident *survey clusters*.

### K-fold validation

Ten random samples were drawn from the *survey clusters* network and a sub-set of 10% of the *survey clusters* was removed before re-estimating the indicator values as described above. The indicator values estimated for each settlement in each of the 10 samples were compared to the value of the indicator obtained using the original (complete) data. These comparisons are summarized using the Bias, Root Mean Square Error (RMSE) and Mean Absolute Difference.

## Data Records

The SEEDNet datasets^1^ are available at Borealis Canada in the SEEDNet Dataverse (https://doi.org/10.5683/SP3/7XSBPS). The datasets^61^ corresponding to the 10 countries included in the technical validation are organized as a set of 10 shapefiles, including the settlement polygons of each country. The shapefiles are projected using EPSG:4326 - WGS84 and contain, as features, the predicted prevalence of each of the 15 indicators assessed in this paper, the proportion of the country population within each settlement, and the total settlement population count.

A list of additional shapefiles representing other countries and survey years is provided in the SEEDNet library^1^, including additional countries, and is available on the SEEDNet Dataverse in Borealis Canada. Other countries and indicators will be added as new surveys become available. Currently, the 98 surveys covering 52 countries in the library^1^ are: Albania-2017, Angola-2015, Armenia-2010, Armenia-2015, Bangladesh-2011, Bangladesh-2014, Bangladesh-2017, Bangladesh-2022, Benin-2011, Benin-2017, Burkina Faso-2010, Burkina Faso-2021, Burundi-2010, Burundi-2016, Cambodia-2010, Cambodia-2014, Cambodia-2021, Cameroon-2011, Cameroon-2018, Chad-2014, Colombia-2009, Comoros-2012, Congo, Democratic Republic-2013, Coted’Ivoire-2011, Cote d’Ivoire-2021, Dominican Republic-2013, Egypt-2014, Ethiopia-2011, Ethiopia-2016, Ethiopia-2019, Gabon-2012, Gabon-2019, Gambia-2019, Ghana-2014, Ghana-2022, Guatemala-2014, Guinea-2012, Guinea-2018, Haiti-2012, Haiti-2016, Honduras-2011, Jordan-2012, Jordan-2017, Jordan-2023, Kenya-2014, Kenya-2022, Kyrgyz Republic-2012, Lesotho-2014, Lesotho-2023, Liberia-2013, Liberia-2019, Madagascar-2021, Malawi-2010, Malawi-2015, Mali-2012, Mali-2018, Mauritania-2019, Mozambique-2011, Mozambique-2022, Myanmar-2015, Namibia-2013, Nepal-2011, Nepal-2016, Nepal-2022, Niger-2012, Nigeria-2013, Nigeria-2018, Pakistan-2017, Philippines-2017, Philippines-2022, Rwanda-2010, Rwanda-2014, Rwanda-2019, Senegal-2010, Senegal-2012, Senegal-2014, Senegal-2015, Senegal-2016, Senegal-2017, Senegal-2018, Senegal-2019, Senegal-2023, Sierra Leone-2013, Sierra Leone-2019, South Africa-2016, Tajikistan-2012, Tajikistan-2017, Tanzania-2009, Tanzania-2015, Tanzania-2022, Timor-Leste-2016, Togo-2013, Uganda-2011, Uganda-2016, Zambia-2013, Zambia-2018, Zimbabwe-2010, Zimbabwe-2015.

Each dataset (shapefile) contains the following features: File name, composed of the country alpha-3 code, survey year, and the _sett_indic stub (e.g. ben_2017_sett_indic.shp corresponds to Benin, 2017). Within the file, each settlement has a unique settlement code (SETT_CODE) that includes the country alpha-3 code and a unique number (e.g. BEN003269), the settlement population (SETT_POP), the fraction of the national population within the settlement(SETT_FRAC), the proportion of the settlement population with access to electricity (ELEC_H_ELC), improved water source (SRCE_H_IMP), iodized salt intake (IODZ_HLIOD), prevalence of anemia among women (ANEM_W_ANY), proportion of women who attended four or more antenatal care visits for their last most recent live birth or stillbirth (ANCN_W_N4P), prevalence of low birth weight (newborns weighing less than 2.5 kg) among the live births in the two years preceding the survey who were weighed at birth (SZWT_C_L25), proportion of children aged 0-5 months living with their mother who were exclusively breastfeeding (breastfeeding and given nothing else) on the day preceding the interview (IYCB_C_EXB), proportion of living children age 12–23 months receiving BCG vaccine according to vaccination card or mother’s report (VACC_C_BCG), proportion of living children age 12–23 months receiving the third dose of the DPT vaccine according to vaccination card or mother’s report (VACC_C_DP3), proportion of living children age 12–23 months receiving the third dose of the Measles vaccine according to vaccination card or mother’s report (VACC_C_MSL), proportion of children age 0-59 months who slept under an insecticide-treated net (NETC_C_ITN), proportion of living children age 0-59 months with diarrhea during the two weeks preceding interview who were given fluid from oral rehydration salts packet or pre-packaged ORS fluid (DIAT_C_ORS), stunting prevalence, or proportion of children age 0-59 months whose height-for-age z-score is 2.0 standard deviations (SD) below the median of the WHO Child Growth Standards (NUTS_C_HA2), wasting prevalence, or the proportion of children < age 0-59 months whose weight-for-height z-score is 2.0 standard deviations (SD) below the median on the WHO Child Growth Standards (NUTS_C_WH2), and proportion of children whose hemoglobin count is less than 11 grams per deciliter (ANMC_C_ANY). The complete codebook for the library in pdf format can be accessed through this URL^62^.

### Technical Validation

We used LIDW to generate high-resolution health indicator base maps for 15 indicators in 10 countries drawing on estimates from each country’s most recent household survey. These maps were used to produce aggregated estimates of indicators for all identified settlements by averaging the value of the pixels that fall within the defined settlement boundaries. Three validation strategies were used: (1) For the initial assessment and benchmarking during method development, the estimates were compared to previously published high-resolution maps produced using a different method. Subsequently, (2) we conducted leave-one-out cross-validation using the network of *survey clusters* (source data) as the units of analysis, and (3) using the output maps produced using our methodology, we simulated out-of-sample *predictions* through another leave-one-out cross-validation using the population settlements as the units of analysis.

### Benchmarking during method development

#### Comparison of LIDW Estimates to Spatial GLM-based Estimates and to DHS survey estimates

During method development, we benchmarked the regional-level estimates of measles vaccination coverage obtained using our high-resolution base maps to those obtained using the model described by Utazi et al.^20^ as well as to regional estimates published in the DHS survey reports. The Spatial Bayesian Geostatistical Model (BGM) method fits equations using non-DHS covariates to predict measles vaccination coverage in these three countries (Cambodia-2014, Mozambique-2011, and Nigeria-2013). The high-resolution *predictions* of vaccination coverage are estimated using georeferenced covariates selected from a non-spatial frequentist binomial generalized linear model. Covariates are then used in a Spatial Bayesian Geostatistical Model to predict indicator coverage in small areas. The reference maps for the three countries are available from WorldPop^63^. The settlement level vaccination coverage estimates obtained using our method were aggregated into regions for comparison with the published BGM-based estimates and with the regional estimates available in the DHS reports. Our estimates achieved high agreement with the DHS *direct* estimates (Figure 6) as well as with the BGM-based values. Quantification of this agreement using Root Mean Square Distance (RMSD) by region in Cambodia, Mozambique, and Nigeria shows acceptable values of 1.7%, 1.5%, and 1.7%, respectively Table (2). The difference between our estimates and the DHS-reported estimates at the regional level was, on average, 2.3 percentage points from the DHS estimate in Nigeria, 1.3 percentage points in Cambodia, and 3.3 percentage points in Mozambique. The differences were larger when compared to the BGM-based model, as expected, given those are based on model predictions of small-area coverage instead of direct use of the survey values (Supplementary Table 2).

**Figure 6.**
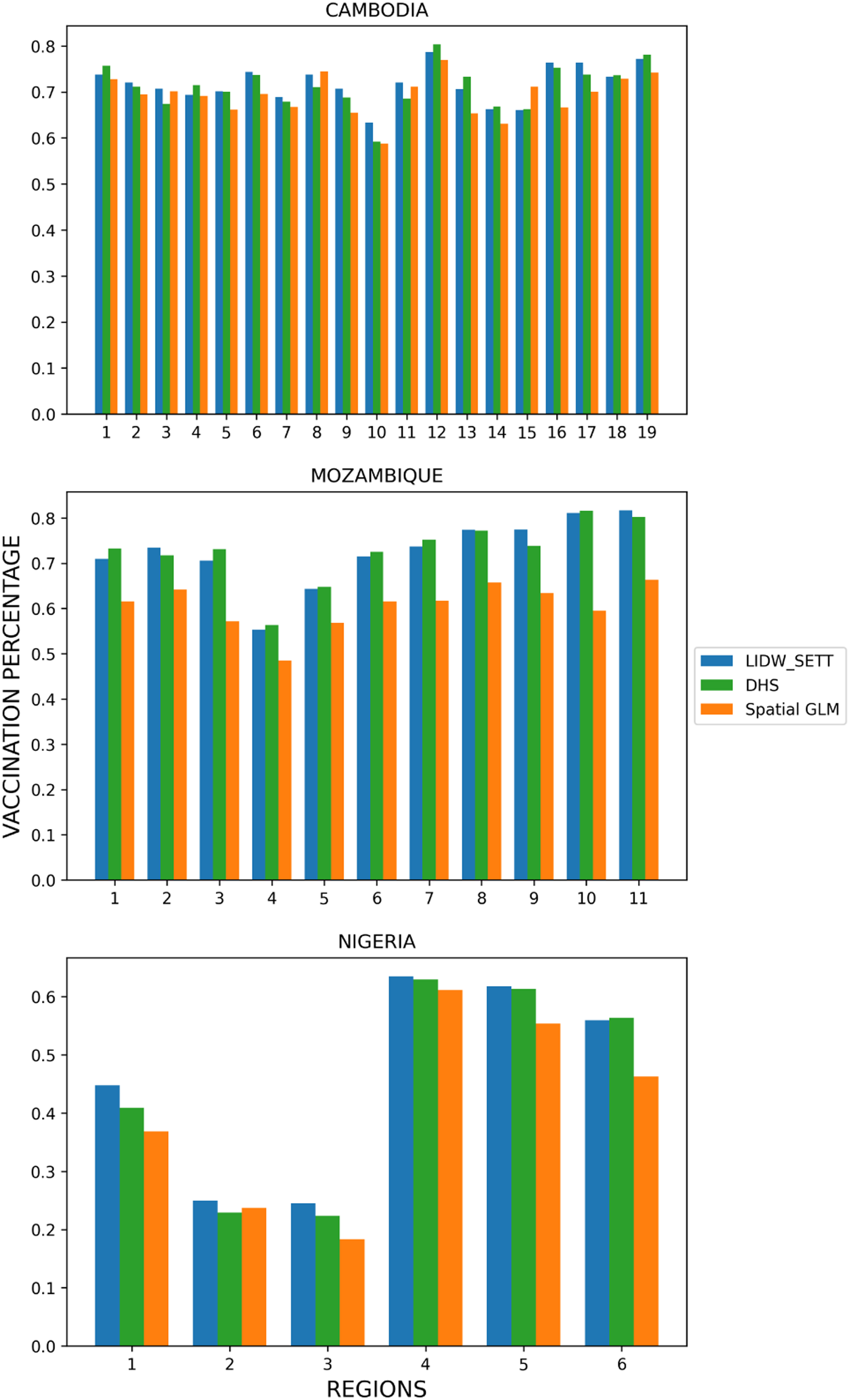
Comparison of the LIDW estimates to the Spatial GLM-based method and DHS regional level estimates. Estimation of the measles vaccination coverage among children under five years of age at the regional level for Cambodia-2014, Mozambique-2011, and Nigeria-2013.

**Table 2.**
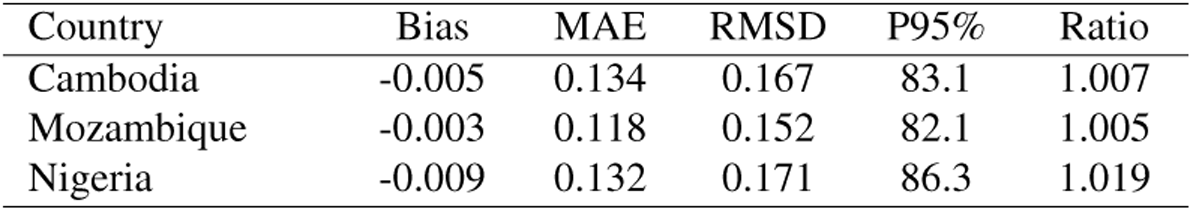
Validation metrics for initial Leave-One-Out Cross-Validation (LOOCV) for measles immunization coverage among children younger than five years in Cambodia, Mozambique and Nigeria.

### Leave-One-Out Cross Validation using the *survey clusters*

Comparisons of the DHS-based estimate for each georeferenced DHS cluster to the LIDW-based estimate for that same DHS cluster through leave-one-out cross-validation are presented in Figure 7. This preliminary assessment is based on randomly selected units sampled from a census grid as described in the DHS survey methodology. These are used as the unit of analysis for internal model validation only as they do not align with the intended use of the survey data or the settlement-level data products presented here. At this model validation stage, the Bias, or systematic discrepancy between *predicted* and *direct* estimates, was low (within ±0.02) for most indicators and did not vary across levels of indicator value (prevalence), of its standard error or its relative standard error (Ratio of the standard error to the value of the indicator). The Bias was comparatively higher (between 0.02 and 0.06) for the infrastructure indicators measured in the DHS surveys, such as households with Access to Electricity and Improved Water Sources. These indicators, in addition to the indicator Iodized Salt Intake in the Household, were included in the validation as negative controls, as they were not expected to be predicted well by our model since their availability is highly influenced by factors such as national policies, in the case of Iodized Salt Intake in the Household, and infrastructure investments, in the case of Access to Electricity and to some extent Access to Improved Water Sources.

**Figure 7.**
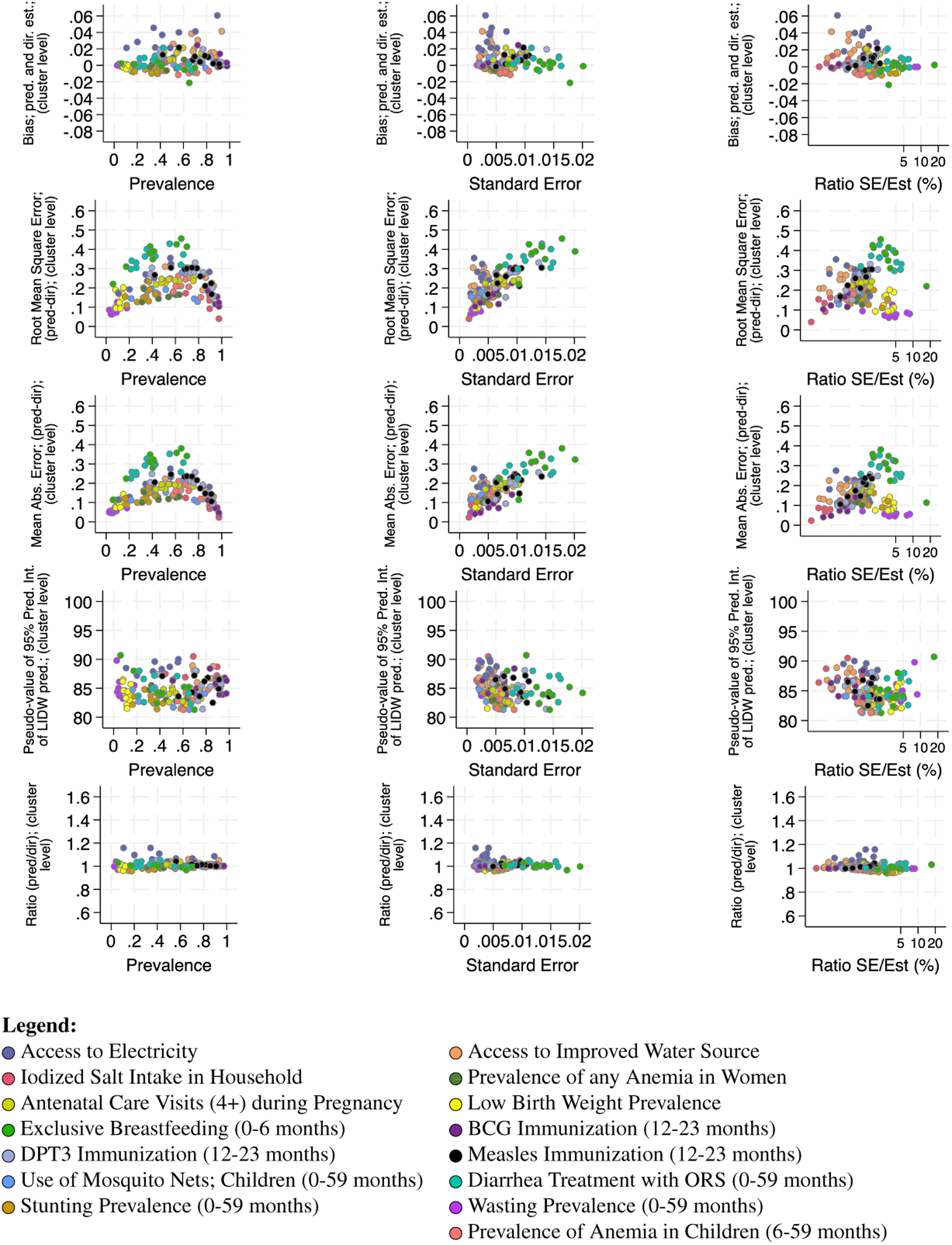
Leave-one-out cross-validation metrics for the network of *survey clusters* (Voronoi cells) used for the LIDW predictions.

Indicators measured with lower precision within the DHS surveys, such as those measured within narrow age groups, narrow time windows, or their combination (e.g., Diarrhea Treatment with Oral Rehydration Salts (ORS), which is measured in a subpopulation of children experiencing diarrhea disease within a narrow time-window of two weeks before the survey) had the highest Root Mean Square Error (RMSE), a performance metric that is more sensitive to outliers (compared to MAE), when the indicator neared 50% and increased with increasing standard error, in line with what is observed in probabilistic sampling (second row in Figure 7). Similarly, the highest Mean Absolute Error (MAE) was seen for these indicators, as seen in the third row in Figure 7. As observed with Bias, infrastructure indicators had higher Mean Absolute Error at lower standard error values. RMSE and MAE were lower for indicators with larger denominators, such as vaccine coverage (Measles, DPT3, BCG).

The preliminary LIDW model *predictions* for this sparse and randomly (by nature of the DHS sampling strategy) defined sample of *survey clusters* are accurate and are not biased estimates of the original survey estimates for the same areas, regardless of indicator value (prevalence) or quality (captured by the relative standard error). The Pseudo-value of the 95% Prediction Interval (P95%; shown in the fourth row in Figure 7) was between 80 and 90% across all indicators. The Ratio between the average of the *direct* estimates (DHS cluster estimate) and the average *predicted* estimates in the cross-validation was near 1.0 (and between.95 and 1.05) across all health indicators regardless of the indicator value, standard error or relative standard error (bottom row in Figure 7). The two infrastructure indicators performed slightly worse (Ratio between 1.05 and 1.16). It is important to note that the leave-One-Out Cross-Validation (LOOCV) estimates for the DHS Voronoi cell centroids are conservative because errors from all iterations (one per data point) contribute to the estimated performance metrics^64^.

The validation metrics for the model predictions compared to the DHS cluster estimates when aggregated into the administrative level 1 (regional), level 2 (state/province), and settlement level are shown in Figures 8 and 9. The model performance across the range of relative standard error was excellent, with low dissimilarity across the spectrum except for the infrastructure indicators and those estimated with lower precision within the original survey, as discussed above. Even for these, the predictions only performed poorly in very specific scenarios, such as low prevalence and high level of aggregation (as is the case of Exclusive Breastfeeding in Benin, with 6% prevalence, seen as the extreme outlier in the top panels in figure 8). The model prediction accuracy captured by the Ratio of *predicted* to *directly* estimated values of the indicators improved with aggregation of estimates into lower-level administrative areas (level 2, state/province; shown in the middle row in figure 8) and improved further at the settlement level. This can be seen by the tightening of the ratios around 1.0 as we move from administrative level 1 (region, shown in the top row) to administrative level 2 (state/province, shown in the middle row) and to settlement (bottom row) in Figure 8. The only indicators deviating from this trend are infrastructure-related, most notably access to electricity. At this natural level of aggregation, the *predictions* are robust to changes in data quality (standard error, Ratio of the standard error to the indicator value/prevalence) and value of the indicator (prevalence). Both MAD and RMSD tended to increase slightly as the relative standard error increased and tended to stay near 0.1 and below 0.2 for most of the indicators. The relative (to the maximum estimates and maximum predictions) Mean Absolute Differences between *predicted* and *directly* estimated values and the Mean Absolute Difference by aggregation level are shown in Figure 9. In summary, the Ratio (Figure 8) and the various measures of mean absolute deviance (Figure 9) confirm the good performance of the model *predictions*, especially at the level of aggregation for which this method is intended to produce estimates (settlement level). The exceptions are infrastructure indicators, most notably access to electricity. Files with the summary validation metrics for all indicators and countries shown in Figures 7, 8 and 9 as well as the raw files used to produce the summary metrics are available on GitHub and on the SEEDNet Dataverse^1^ in Borealis Canada.

**Figure 8.**
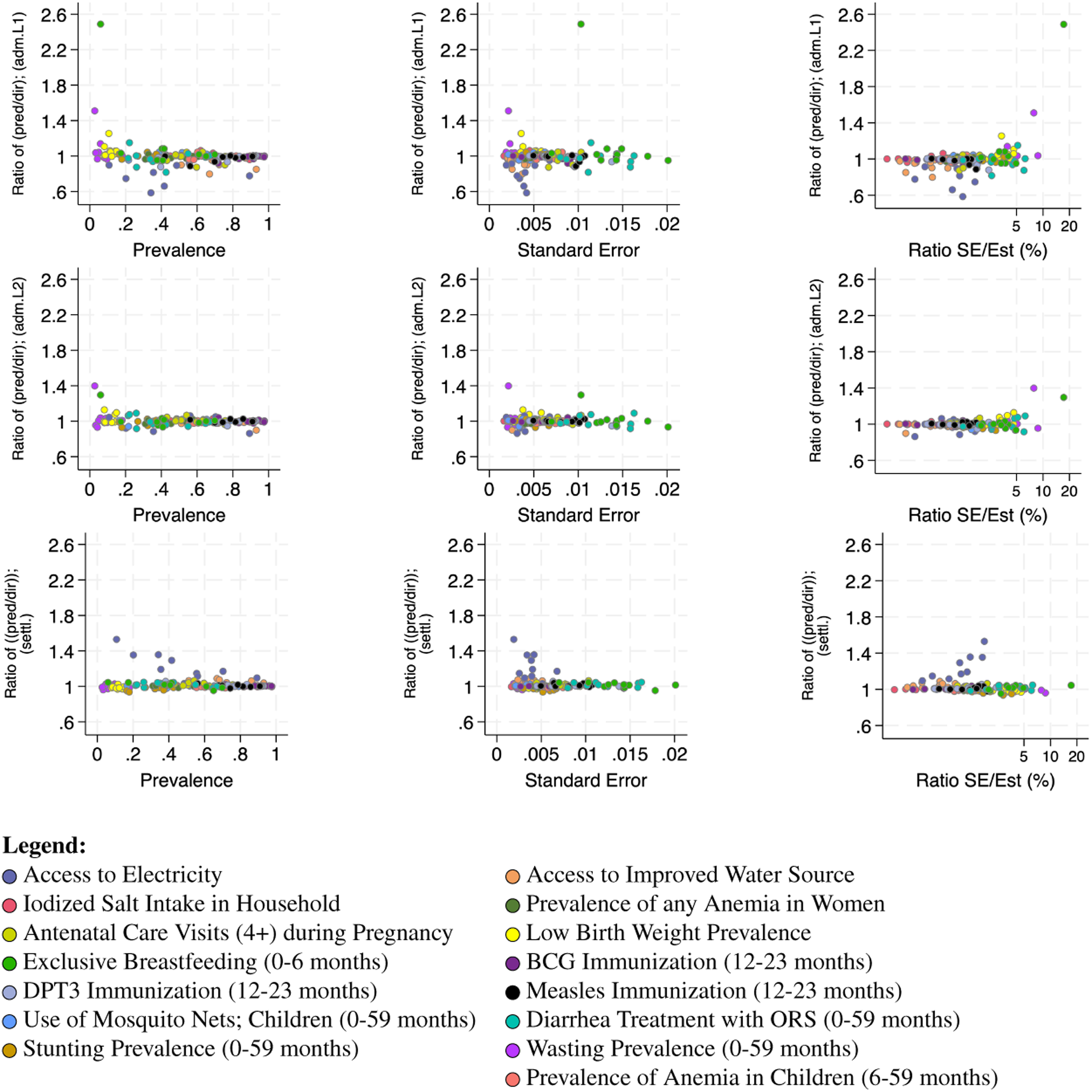
Validation metrics (Ratio of LIDW *predictions* to DHS *direct* estimates) for the LIDW *predictions* compared to DHS *direct* estimates by aggregation level for the 15 indicators and ten countries.

**Figure 9.**
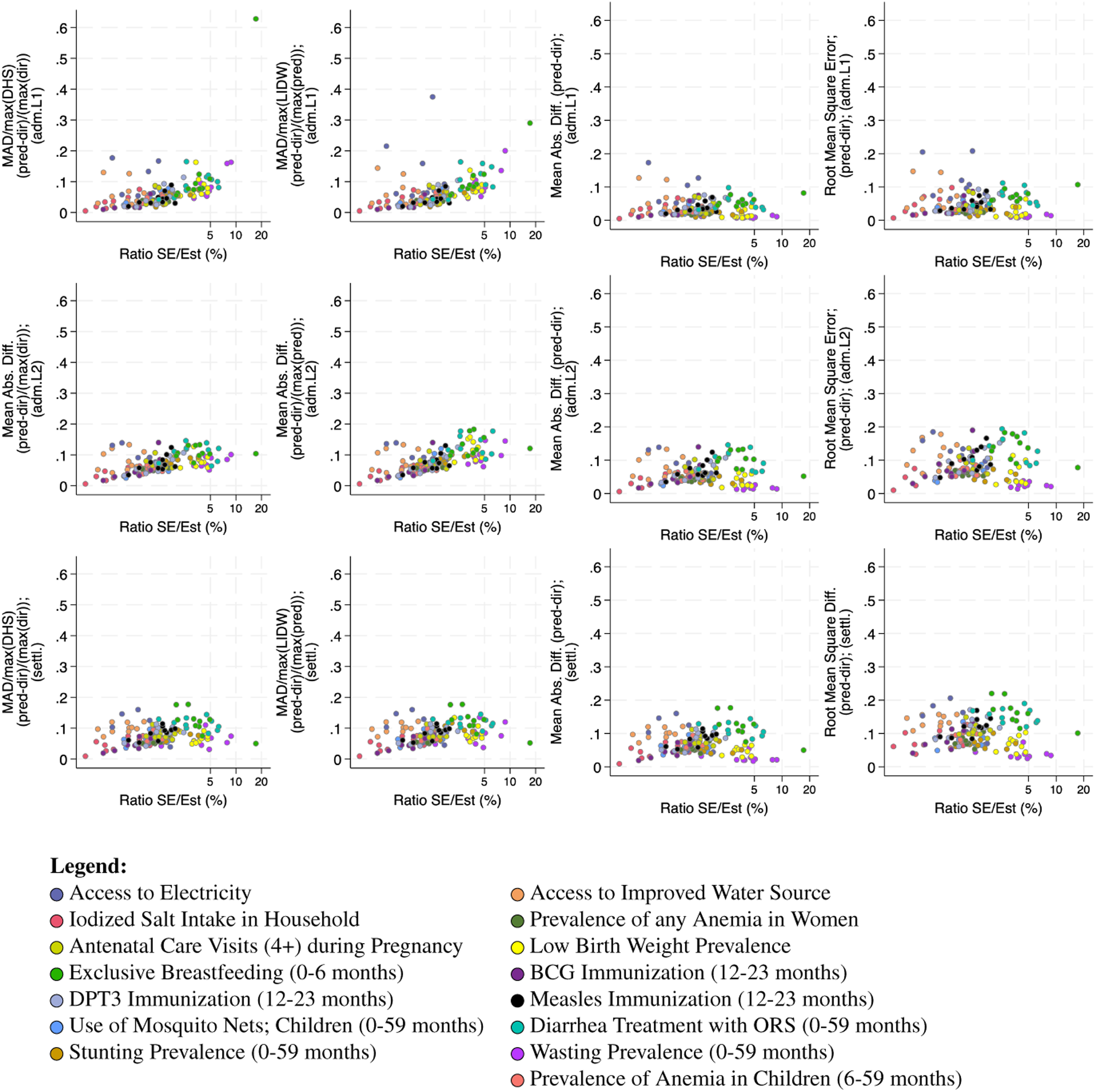
Additional validation metrics for the LIDW *predictions* compared to DHS *direct* estimates by aggregation level for the 15 indicators and ten countries.

### Cross-validation of out-of-sample *predictions* - Comparison of LIDW to DHS Estimation at the Settlement Level

The leave-one-out cross-validation included 10,917 settlements within the ten validation countries. Summary results comparing the simulated out-of-sample *predicted* and *direct* estimates for each of the 15 indicators within each of the 10,917 settlements in the ten validation countries are shown in Figure 10. Overall performance of the model predictions at the settlement level was very good; indicators with coverage near the 50% value tended to have slightly higher RMSD and MAD, but overall values were low, indicating high similarity (low RMSD and MAD) at the settlement level validation across most indicators. The similarity was highest for indicators measured with higher precision in the original survey (lowest standard error). Indicators of access to infrastructure (Access to Improved Water Source and Electricity) had relatively higher dissimilarity (highest RMSD and MAD) despite being estimated with high precision (low standard error) and high relative precision (Ratio of standard error to indicator value). The highest Bias was also observed for the infrastructure indicators, with the remaining being estimated with relatively low Bias (within 0.02 of the *direct* estimate). Supplementary Figure 3 shows these results with the color of the markers indicating the survey instead of the indicator type and suggests no trends or clustering of performance indicators by country.

**Figure 10.**
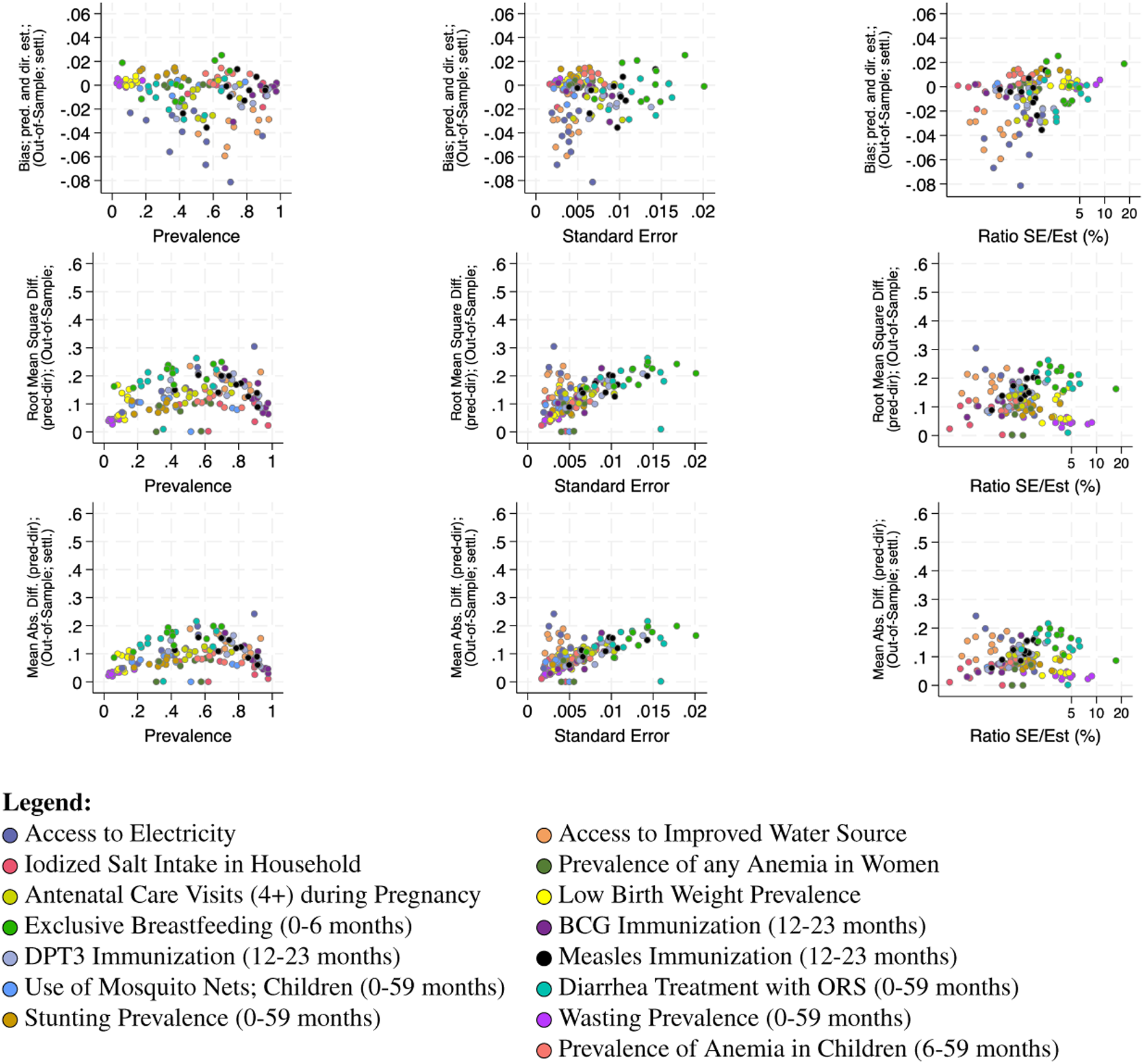
Validation metrics (Bias, Root Mean Square Difference and Mean Absolute Difference of LIDW *predictions* and DHS *direct* estimates) by indicator prevalence, standard error and relative standard error (s.e./indicator value) for the 15 indicators and ten countries.

For the sub-sample of settlements used in the out-of-sample validation, the *predicted* and *direct* estimates were similar across all countries as shown in Figure 11. Overall, the difference between *predicted* and *direct* estimates across all the 15 indicators was small but increased for the largest settlements. With the exception of Access to Electricity and Improved Water Source, the distribution of the difference between *predicted* and *direct* estimates had a median of zero for settlements up to an area of at least 10^5^ pixels and within settlements of these sizes it was slightly more variable within the smallest settlements. Similarly, differences between the *predicted* and *direct* estimates increased when a larger number of *survey clusters* were excluded from the estimation sample but were very robust when estimates for a given settlement were based on between 1 and 10 clusters as is shown in Figure 12. Within the countries the wider range of differences between the *predicted* and *direct* estimates were observed for indicators measured with less precision in the original survey, for the reasons noted above. The settlements with the smallest area (1 pixel) and the largest areas had more systematic differences (medians above or below one) between the *predicted* and *direct* estimates. Settlement area was independently associated with small but increasing differences between *predicted* and *direct* estimates, as was the number of clusters excluded from the estimation sample, but the direction of the effect varied by indicator, and their magnitude was small (Supplementary Table 3). The changes in the differences between the *predicted* and *direct* estimates for all indicators in each of the 10 countries by settlement area are shown in the Supplementary Figures 4 to 18. There were larger differences in the simulated out-of-sample predictions for settlements whose original values were based on multiple *survey clusters* across most, but not all, indicators. These findings are limited to this specific validation step, where we removed all estimation units (*survey clusters*) that overlap with a settlement to assess the predictions in a simulated scenario where observed settlements had their values re-estimated as if no *survey clusters* overlapped with their boundaries. For very large settlements, this resulted in larger than usual distances between the remaining estimation units, and for small settlements, it may have resulted in using information from very distant remaining clusters. The differences between the *predicted* and *direct* estimates for all indicators in each country by number of clusters excluded from the estimation sample are shown in Supplementary Figures 19 to 33. The DHS cluster networks for all ten countries are shown in Supplementary Figure 34.

**Figure 11.**
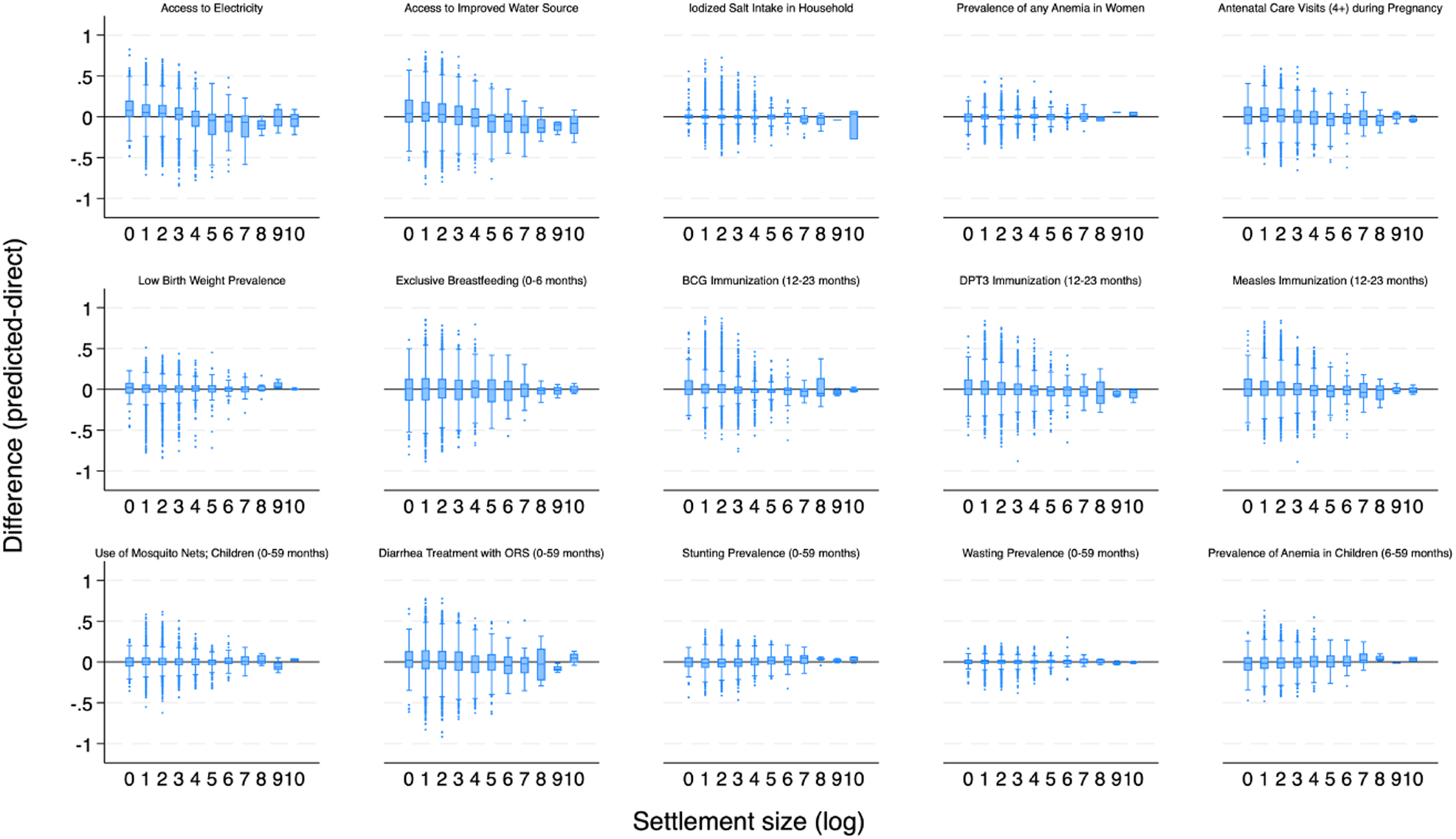
Difference between LIDW *predictions* and DHS *direct* estimates) for each of the 15 indicators across 10,917 settlements in the 10 countries by settlement area in pixels (log).

**Figure 12.**
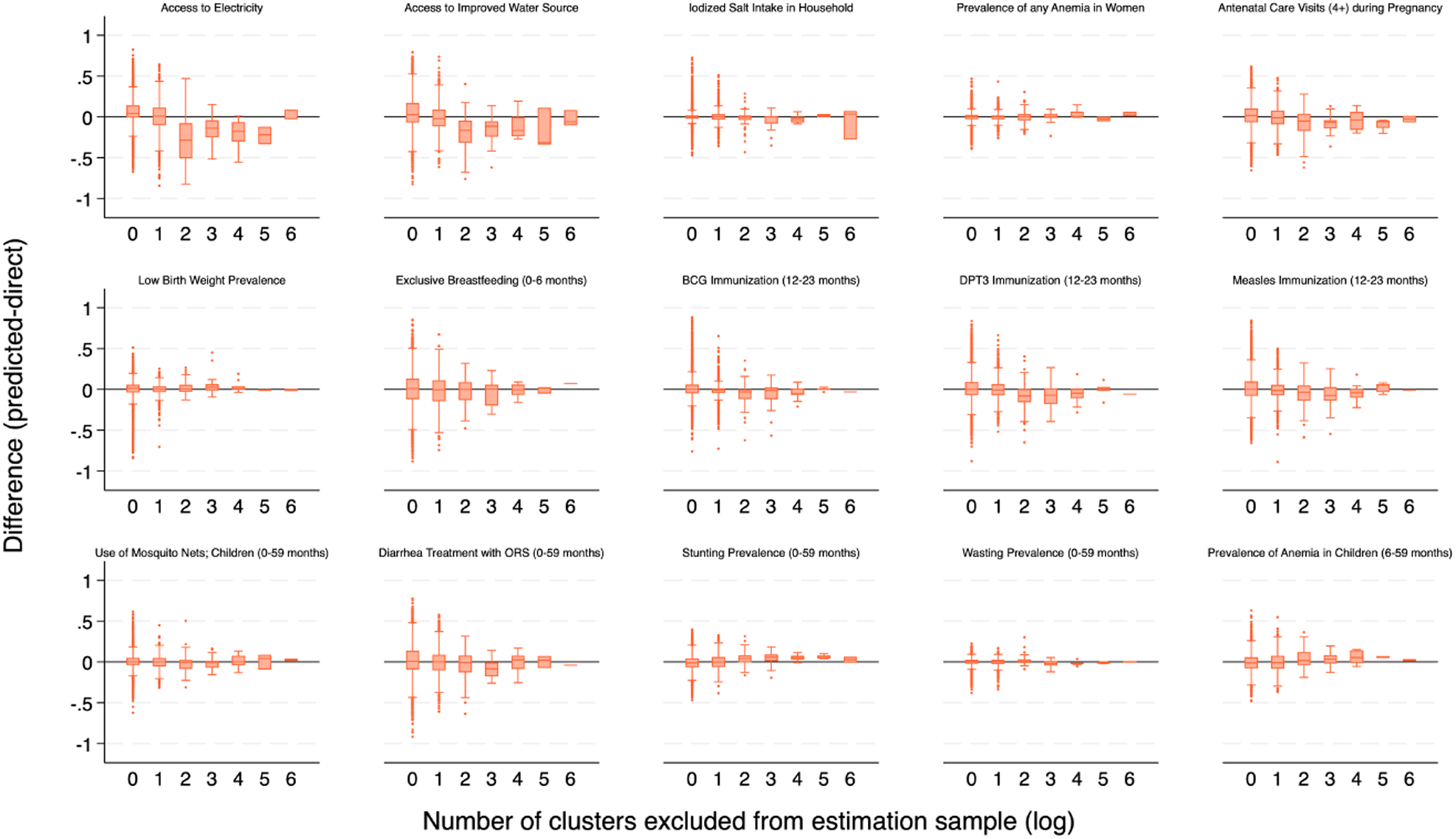
Difference between LIDW *predictions* and DHS *direct* estimates) for each of the 15 indicators across 10,917 settlements in the 10 countries by number of *survey clusters* excluded from the estimation sample (log).

The files containing the summary validation metrics for all indicators and countries that are shown in Figures 11 and 12, Supplementary Figures 4 to 18 and Supplementary Figures 19 to 33 as well as the complete data files used to produce the summary metrics, including the *predicted* and *direct* estimates for all 147,548 settlements included in the Leave-One-Out Cross-Validation can be downloaded from GitHub or from the SEEDNet Dataverse^1^ in Borealis Canada.

### K-fold validation

Figure 13 shows the validation metrics for the K-fold validation using 10 folds of 10% of the sample. Supplementary Figures 35 to 49 show the K-fold validation results as scatterplots comparing the *predicted* and *direct* estimates within all settlements (n=101,435) for all 10 countries and 15 indicators. The distribution of the settlements by settlement area (in log pixels; log refers to the natural logarithm throughout the manuscript) across the ten countries used in the technical validation is shown in Figure 14 and the stratified distributions by country are shown in the Supplementary Figure 2. The RMSE estimates were below 0.03, and the R^2^ was above 0.97 for all indicators across all countries. Table 3, and Supplementary Tables 4 and 5 show the summary results of this validation for all countries and indicators. The performance of the predictions was very good, and as noted in previous assessments, indicators measured with less precision in the original survey performed worse, relative to the overall very good validation results, as did infrastructure indicators. Bias was overall highest for Access to Electricity and Improved Water Sources, but still no higher than 0.8 percentage points. The Bias for standard indicators of child growth and nutrition (Stunting and Wasting) was lower than 0.5 percentage points across all 10 countries and their average was below 0.1 percentage points. Similarly, good performance was observed for immunization indicators and pregnancy and delivery indicators across all countries (Supplementary Table 4). The Root Mean Square Error and Mean Absolute Deviance were no higher than 3 and 2 percentage points, respectively, for any indicator (Table 3 and Supplementary Table 5).

**Figure 13.**
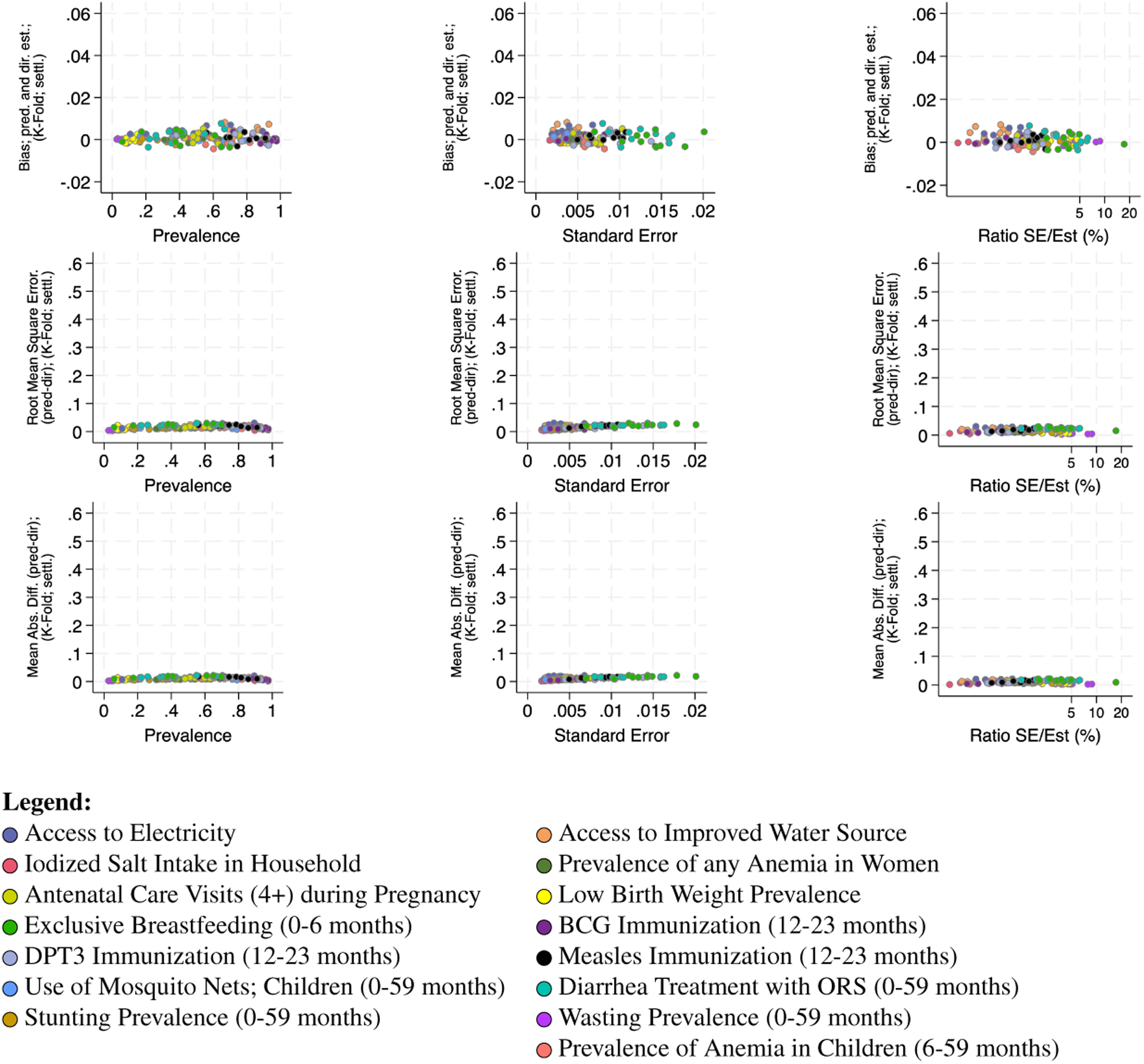
K-Fold validation metrics for the LIDW *predictions* compared to DHS *direct* estimates at the settlement level for the 15 indicators and ten countries.

**Figure 14.**
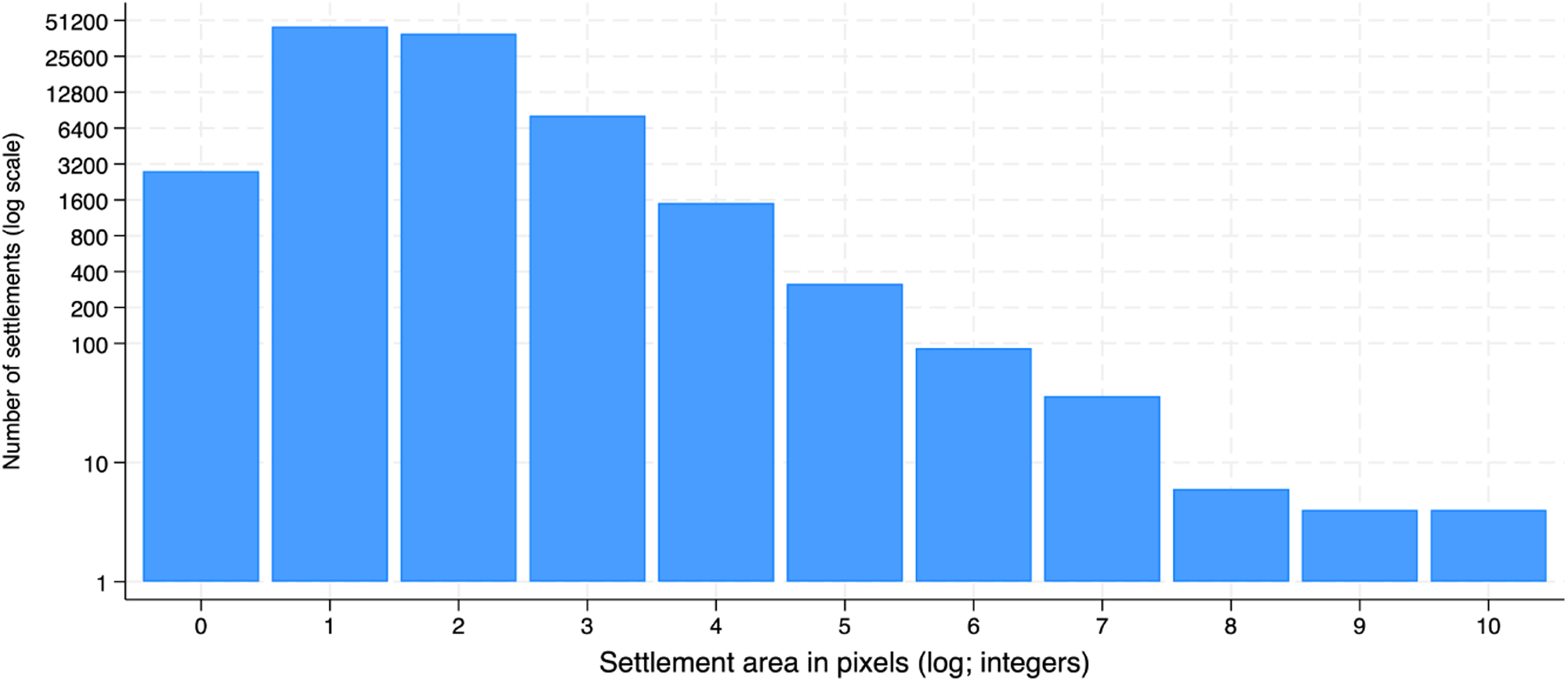
Distribution of the settlements (n=101,435) across all ten surveys by area size. Logarithmic scale.

**Table 3.** K-Fold validation results. Root Mean Square Error (RMSE) of the model predictions for each settlement across the 10 validation countries for each of the 15 indicators assessed.

| Indicator | Angola-2015 | Benin-2017 | Cambodia-2014 | Gabon-2012 | Malawi-2015 | Mali-2018 | Mozambique-2011 | Nigeria-2013 | Senegal-2019 | Zambia-2018 |
| --- | --- | --- | --- | --- | --- | --- | --- | --- | --- | --- |
| **RMSE** |  |  |  |  |  |  |  |  |  |  |
| Access to Electricity | 0.016 | 0.016 | 0.028 | 0.031 | 0.011 | 0.017 | 0.013 | 0.025 | 0.029 | 0.020 |
| Improved Water Source | 0.026 | 0.018 | 0.020 | 0.018 | 0.021 | 0.023 | 0.025 | 0.023 | 0.023 | 0.021 |
| Iodized Salt Intake | 0.013 | 0.004 | 0.014 | 0.006 | 0.014 | 0.011 | 0.012 | . | . | 0.016 |
| Prev. of Anemia; Women | . | 0.015 | 0.010 | 0.014 | 0.016 | 0.011 | 0.013 | . | . | 0.010 |
| Antenatal Care (4+) | 0.017 | 0.017 | 0.018 | 0.020 | 0.014 | 0.016 | 0.019 | 0.018 | 0.017 | 0.016 |
| Low Birth Weight Prevalence | 0.012 | 0.008 | 0.007 | 0.013 | 0.007 | 0.017 | 0.013 | 0.023 | 0.007 | 0.006 |
| Exclusive Breastfeeding (0-5m) | 0.022 | 0.020 | 0.029 | 0.015 | 0.030 | 0.024 | 0.027 | 0.021 | 0.025 | 0.024 |
| BCG Immun. (12-23m) | 0.025 | 0.017 | 0.014 | 0.019 | 0.010 | 0.022 | 0.016 | 0.017 | 0.008 | 0.009 |
| DPT3 Immun. (12-23m) | 0.020 | 0.017 | 0.018 | 0.021<sup>a</sup> | 0.013 | 0.019 | 0.020 | 0.019 | 0.010 | 0.012 |
| Measles Immun. (12-23m) | 0.023 | 0.021 | 0.025 | 0.023 | 0.013 | 0.020 | 0.019 | 0.018 | 0.014 | 0.015 |
| Bed Net Use; Children (0-59m) | 0.013 | 0.010 | . | 0.013 | 0.013 | 0.008 | 0.013 | 0.010 | 0.017 | 0.012 |
| Diarrhea Treat.; ORS (0-59m) | 0.022 | 0.022 | 0.024 | 0.022 | 0.023 | 0.020 | 0.030 | 0.021 | 0.024 | 0.026 |
| Stunting Prevalence (0-59m) | 0.012 | 0.010 | 0.013 | 0.012 | 0.017 | 0.008 | 0.011 | 0.010 | 0.009 | 0.012 |
| Wasting Prevalence (0-59m) | 0.006 | 0.003 | 0.009 | 0.004 | 0.004 | 0.005 | 0.005 | 0.008 | 0.005 | 0.005 |
| Anemia; Children (6-59m) | 0.018 | 0.013 | 0.016 | 0.019 | 0.017 | 0.010 | 0.016 | . | . | 0.012 |

Note: <sup>a</sup> For our estimates of the Gabon 2012 DHS, children 12-23m were deemed to have received three doses of DPT if they received three doses combined of any of DPT containing vaccines prevalent at the time (i.e: DTPCoq, pentavalent, Pentacoq, Pentaxim and Tetracoq)

The files containing the summary validation metrics for all indicators and countries that are shown in Figures 13 and 14, Supplementary Figure 2 and Supplementary Tables 4 to 5 as well as the complete data files used to produce the summary metrics, including the *predicted* and *direct* estimates for all 1,048,574 settlements included in the K-fold validation can be downloaded from GitHub or from the SEEDNet Dataverse^1^ in Borealis Canada.

In summary, here we describe a novel method and establish a public-use pipeline that produces harmonized small-area estimates of health indicators. The method uses existing standard national surveys and global map products, reducing data input requirements to a minimum without the need for country-survey-specific model fitting. This greatly reduces sources of variability to the single source of input data. The method can be implemented automatically using open sources of data and can be extended to cover additional indicators while continuing to require no parameter selection or context-and indicator-specific modeling decisions. The method is based on Local IDW interpolation and incorporates weighting exclusively for a subset of the nearest known points. It accomplishes this by dividing the country map into 1 km^2^ pixels and estimating indicator values using georeferenced *survey clusters*, associating each pixel with a Voronoi cell, and estimating indicator values for the unknown pixels within the polygon using the representative point within each polygon and neighboring polygons.

Validation results showed that this method performs very well across indicators, with lower RMSE values for indicators measured with higher precision, such as those measured among all respondents within a sampling unit. Indicators calculated using narrow age bands or specific sub-samples result in only relatively lower performance. The validation procedures shown here compare LIDW estimates for each settlement to DHS-based settlement estimates. Although these are the best available estimates for validation, they must be interpreted with caution as the DHS surveys are not designed to produce reliable estimates at the settlement level.

### Usage Notes

Settlement boundaries were extracted automatically using GDAL^60^ from the GHSL map products (GHS-SMOD R2023A; for a detailed description of the procedures, please refer to the Settlement identification subsection). As discussed by the GHSL authors, the built-up area classification may miss some settlements not captured by satellite imagery, such as those constructed under dense vegetation or tree canopies, those built under rock formations, on cliff-sides or underground^65^.

Our datasets and maps are intended for use at the validated (settlement level) or higher level of aggregation. Higher-resolution estimates of health indicators produced during the intermediate steps of our estimation procedures, specifically the grid-level estimates within the settlements are not intended for research use as these cannot be validated due to the lack of a standard or reference for multi-country high-resolution estimates. For this reason, the output files do not include those intermediate estimates.

Settlement level estimates for the largest urban settlements as well as other extensive continuously populated areas have been validated but they should be interpreted with caution in studies focusing exclusively on these areas. This is due to the small number of such large settlements (settlements with a pixel area larger than 10^6^) available for validation within each country. All settlements were defined using a standard criteria in our datasets, but we believe these largest polygons may not fit the functional definition of a community given their territorial extension, being likely to encompass multiple functional communities within its boundaries. Dividing these settlements into smaller units may be justified depending on the research objectives. Similarly, the appropriateness of their direct use (as is) as nodes in networks should be assessed by researchers on a case-by-case basis, accounting for research objectives, contextual factors as well as the desired level of inference. Upcoming releases of the datasets incorporate the partition of large settlements into smaller areas centered around population peaks. The rationale for this process and its method is described in the supplementary appendix.

The technical validation was executed in a variety of system configurations. The computation time for 10 countries and 15 indicators is just over 24 hours on an Apple M1 Ultra chip, featuring 128 GB of memory and 20 logical CPUs. This computation time is 3-4 times greater on a system with lower processing capacity, specifically a 2.4 GHz 8-Core Intel Core i9 with 16 logical CPUs. The preparation steps, including settlement identification for the 10 countries, are completed in approximately 10 minutes, depending on the download speeds of the input files. The preparation, estimation, and validation of the settlement-level estimates for 10 countries and 15 indicators require approximately 24 hours on the faster of the two systems tested. The provided code is configured for 40 parallel processes, a setting that can be adjusted as needed. The preparation and estimation phases, excluding validation, can be completed in approximately 2 hours for all 15 indicators across the 10 surveys evaluated. There is considerable variability in computation time, which is influenced by the size of the input survey and the number of settlements within the country.

## Code availability

Detailed instructions and the code to replicate our findings and to produce maps for additional indicators and countries is available on GitHub and on the SEEDNet Dataverse^1^ in Borealis Canada. The code and documentation include details on replicating our results and reproducing our data outputs.

The code used to replicate the datasets and their validation is divided into four parts: (1) The code to calculate the indicators for each survey cluster (R) (2) The code to estimate the indicators by settlement (Python) (3) The validation of the estimates (Python) (4) The code to reproduce all Tables and Figures (Python and Stata) Part 1 requires access to the relevant DHS datasets from the DHS program. The output from part 1 is provided as a.csv file. Details of the content of each folder can be found in the README.txt file.

## Data Availability

All data produced are available at Borealis Canada in the \href{https://borealisdata.ca/dataverse/SEEDNet}{SEEDNet Dataverse} (https://doi.org/10.5683/SP3/7XSBPS).

https://borealisdata.ca/dataverse/SEEDNet

https://github.com/DiegoGBassani/SEEDNet

## Acknowledgements

This research was funded by the New Frontiers Research Foundation (NFRF) under grant number NFRFE-2020-01630. The early stages of this project were carried out with the aid of a grant from Nutrition International, Ottawa, Canada, through the financial assistance of the Government of Canada through Global Affairs Canada (CA-3-P005253002).

## Author contributions statement

A.H.D., J.L.K., K.M., S.C. and D.G.B. conceived the study, A.H.D., J.L.K. and C.G. acquired and assembled the raw data, A.H.D. and J.L.K. prepared the datasets and the analytical code, A.H.D., J.L.K., and D.G.B. performed the technical validation of the raw data and final datasets, A.H.D., J.L.K. and D.G.B. analysed the results, A.H.D and J.L.K. produced the final datasets, A.H.D., J.L.K., and D.G.B. prepared the tables and figures, A.H.D., J.L.K., K.M., S.C., and D.G.B. drafted the manuscript. All authors read and approved the final version of the manuscript.

