## Supplementary Material for "SEEDNet: Covariate-free multi-country settlement-level epidemiological estimates datasets for network analysis"

#### ABSTRACT

Detailed instructions and the code to replicate our findings and to produce maps for additional indicators and countries is available on GitHub and on Borealis Canada. The DHS survey datasets and their corresponding geo-referenced files necessary to reproduce our study from the raw original data are openly available but require user registration for access. Requests to access the data can be made through the DHS program website. At the time of writing this manuscript, access to the remaining input datasets was available without requiring registration. The code and documentation include details on replicating our results and reproducing our data outputs.

### Statistical Measures for Error

Typically, after conducting an assessment, we obtain two lists: one containing the *predicted* values and another containing the corresponding actual values. These lists serve as the basis for evaluating the performance and accuracy of the prediction model or method employed during the assessment.

When comparing two lists of predicted and actual values, several measures are available to quantify their similarity or distance. The root mean square error (RMSE) is a commonly used measure that indicates the extent to which the two lists differ. It provides a numerical value representing the average magnitude of the differences between corresponding elements in the lists, thereby indicating their overall dissimilarity.

$$RMSE = \sqrt{\frac{1}{N}\sum_{\circ\in \mathrm{DHS}} |v_\circ - \hat{v}_\circ|^2} \tag{1}$$

where $v_\circ$ ($\hat{v}_\circ$) is actual (predicted) values for DHS points at $\mathbf{x}_\circ$. The summation goes over all clusters in the survey dataset, $N$ stands for the total number of clusters. Likewise, the Mean Absolute Error (MAE) can be defined for the same purpose,

$$MAE = \frac{1}{N}\sum_{\circ\in \mathrm{DHS}} |v_\circ - \hat{v}_\circ| \tag{2}$$

Bias shows the systematic discrepancy between actual and predicted values. In other words, it indicates the inadequacy of the prediction method in producing accurate results. The following equation is used for the calculation of bias.

$$Bias = \sum_{\circ\in \mathrm{DHS}} (v_\circ - \hat{v}_\circ) \tag{3}$$

In addition to bias, assessing the ratio between the average *direct*estimates and the average predicted estimates is important. A ratio close to one indicates that no significant overestimation or underestimation is occurring.

$$Ratio = \frac{< v_\circ >}{< \hat{v}_\circ >} \tag{4}$$

In fact, the weights are probabilities by definition, Eq. 2 and we can calculate the standard deviation, $\Delta_\circ$ for a given survey cluster,

$$\Delta_\circ^2 = \sum_{\circ'\in\mathscr{N}(\mathbf{x}_\circ)} w_{\circ'}(v_{\circ'} - \hat{v}_\circ)^2 \tag{5}$$

Now it is useful to know whether an actual value of health indicator in a cluster, $v_\circ$ falls within the interval $[\hat{v}_\circ - 1.96\Delta_\circ, \hat{v}_\circ + 1.96\Delta_\circ]$. The fraction of *survey clusters* that meet this condition shows how the actual data are scattered around the predicted ones. Compared to a similar statistical concept, we call this fraction the prediction interval ($PI95\%$). For a normal distribution of errors, the fraction is equal to 0.95.

### Partition of large settlements

For the settlements spanning vast areas, the geographical extent does not support the assumption of frequent interaction between most of its inhabitants used to define settlements in this study. Our definition of settlement is intended to capture the space where the people interact with one another most frequently on a near-daily basis. It relies on the spatial structure of a population that typically defines the areas where people live and commute on a daily basis. These areas are generally centered around population peaks, defined as the location (grid cell) where the population size is greater than that of its surrounding areas. In our datasets, population peaks were identified using a population density raster file by sliding a window of a predetermined size across the raster pixels and applying a condition to detect areas of higher population density. Since population peaks are relative to the population of neighboring areas of the grid, a threshold is used to identify them. Here we used a window size of 11 km and threshold of 0.01 of the maximum peak size to identify significant population peaks within a settlement polygon. Settlements that contained more than one population peak need were split using a Voronoi algorithm to delineate the dominance area for each peak so that within each peak's dominance area all points were closer to that specific peak than to any other peak in the settlement. When this process resulted in disconnected areas we assigned the non-peak areas to the peak area with which they share the largest common boundary.

### Additional Tables

**Supplementary Table 1.** Definition of the survey indicators used in the Technical Validation.

| Indicator | Numerator | Denominator |
|---|---|---|
| **Initial Assessment** | | |
| Measles Immunization (0-59 months)[a] | Living children age 0-59 months receiving the vaccine according to information on vaccination card or mother's report | Living children age 0-59 months |
| **Technical validation**[b] | | |
| Access to Electricity | Households with electricity | All households |
| Access to Improved Water Source | Households whose main source of drinking water is an improved source | All households |
| Iodized Salt Intake in Household | Number of households with salt tested with iodized salt | Number of households where salt was tested for iodine content |
| Percentage of women with anemia | Non-pregnant women whose hemoglobin count is less than 12.0 grams per deciliter (g/dl) plus the number of pregnant women whose count is less than 11.0 g/dl | Women of reproductive age measured for anemia in households selected for anemia testing |
| Antenatal Care Visits (4+) during Pregnancy | Women who attended four or more antenatal care visits for their last most recent live birth or stillbirth | Women who have had a live birth in the two years preceding the survey |
| Low Birth Weight Prevalence | Live births in the two years preceding the survey who were weighed at birth and were reported as weighing less than 2.5 kg | Live births in the last two years who were weighed at birth |
| Exclusive Breastfeeding (0-5 months) | Children aged 0-5 months living with their mother who were breastfeeding and given nothing else the day preceding the interview | Children born 0-5 months preceding the survey who are living with their mother |
| BCG Immunization (12-23 months) | Living children age 12–23 months receiving specified vaccine according to vaccination card or mother's report | Living children age 12–23 months |
| DPT3 Immunization (12-23 months) | Living children age 12–23 months receiving specified vaccine according to vaccination card or mother's report | Living children age 12–23 months |
| Measles Immunization (12-23 months) | Living children age 12–23 months receiving specified vaccine according to vaccination card or mother's report | Living children age 12–23 months |
| Children Slept u/ Bed Net (0-59 months) | Children age 0-59 months who slept under an insecticide-treated net (ITN) | Children age 0-59 months |
| Diarrhea Treatment with ORS (0-59 months) | Living children <59 months with diarrhea during the two weeks preceding interview who were given fluid from oral rehydration salts packet or pre-packaged ORS fluid | Children 0-59 months with diarrhea in the two weeks preceding the interview |
| Stunting Prevalence (0-59 months) | Children <59 months whose height-for-age z-score is below minus 2 (-2.0) standard deviations (SD) below the median on the WHO Child Growth Standards | Living children 0-59 months before the survey who have a valid non-flagged height for age z-scores |
| Wasting Prevalence (0-59 months) | Children <59 months whose weight-for-height z-score is below minus 2 (-2.0) standard deviations (SD) below the median on the WHO Child Growth Standards | Living children 0-59 months before the survey who have a valid non-flagged weight for height z-scores |
| Prevalence of Anemia in Children (6-59 months) | Children whose hemoglobin count is less than 11 grams per deciliter (g/dl) | Children 6-59 months who were measured in households selected for anemia testing |

Note: [a] Definition used only for initial comparison with Utazi *et al.* (see ref. 19 in main paper) in Section Comparison of LIDW Estimates to Spatial GLM-based Estimates. [b] Standard definitions are used in all other stages of the Technical Validation and in the final Settlement-level datasets. For the most recent detailed definitions, we refer to the Guide to DHS Statistics DHS-8: Anemia; Women, Anemia; Children, Wasting, Stunting, Diarrhoea Treatment; ORS, Water Source, Low Birth Weight, Exclusive Breastfeeding, BCG, DPT3 and Measles Immunization, Iodized Salt, Electricity, Antenatal Care.

**Supplementary Table 2.** Comparison of estimates of measles immunization coverage among children younger than five years by region in Cambodia, Mozambique and Nigeria.

| **Country** Region | LIDW-based estimate (settle-ments) | LIDW-based estimate based on grid-estimate (all pixels) within region | Survey-based estimate (cluster buffer zones overlap with region) | Diff. between LIDW-based estimate (settlement) and DHS estimate; (pp) | BGM-based estimate (high-resolution raster); Utazi, 2018 | DHS-estimate (as reported) | Diff. between LIDW (settlement) and BGM-based esti-mate;(pp) |
|---|---|---|---|---|---|---|---|
| **Nigeria** | | | | | | | |
| North Central | 44.8% | 42.3% | 40.9% | 0.9 | 43.0% | 45.7% | 1.7 |
| North East | 24.9% | 23.9% | 22.9% | 0.6 | 24.4% | 25.5% | 0.5 |
| North West | 24.5% | 22.6% | 22.3% | 4.3 | 19.4% | 20.2% | 5.1 |
| South East | 63.5% | 62.8% | 62.9% | 1.8 | 67.1% | 61.7% | 3.6 |
| South South | 61.8% | 61.0% | 61.3% | 2.3 | 63.1% | 59.6% | 1.3 |
| South West | 55.9% | 52.9% | 56.3% | 3.7 | 48.3% | 59.7% | 7.7 |
| | | | **Mean Diff.** | **2.3** | | **Mean Diff.** | **3.3** |
| **Cambodia** | | | | | | | |
| Banteay Meanchey | 73.8% | 73.1% | 75.7% | 2.3 | 77.5% | 76.1% | 3.7 |
| Battambang/Pailin | 72.0% | 71.6% | 71.1% | 0.6 | 76.0% | 71.4% | 4.0 |
| Kampong Cham | 70.7% | 70.7% | 67.4% | 1.1 | 78.6% | 71.9% | 7.8 |
| Kampong Chhnang | 69.4% | 68.4% | 71.4% | 2.5 | 73.5% | 71.9% | 4.1 |
| Kampong Speu | 70.1% | 68.6% | 70.0% | 1.8 | 73.5% | 71.9% | 3.4 |
| Kampong Thom | 74.4% | 73.9% | 73.7% | 2.5 | 69.4% | 71.9% | 4.9 |
| Kampot/Kep | 68.8% | 68.5% | 67.9% | 0.9 | 75.7% | 67.9% | 6.9 |
| Kandal | 73.7% | 73.9% | 71.0% | 0.5 | 83.6% | 73.2% | 9.9 |
| Kratie | 70.7% | 70.4% | 68.8% | 0.4 | 68.3% | 71.1% | 2.4 |
| Mondul/R. Kiri | 63.3% | 64.4% | 59.2% | 0.2 | 60.0% | 63.2% | 3.4 |
| Otdar Meanchey | 72.0% | 72.5% | 68.6% | 0.3 | 71.8% | 71.7% | 0.2 |
| Phnom Penh | 78.7% | 78.7% | 80.3% | 1.3 | 88.0% | 80.0% | 9.4 |
| P. Sihanouk/K. Kong | 70.7% | 70.6% | 73.2% | 3.6 | 64.5% | 74.3% | 6.1 |
| P. Vihear/S. Treng | 66.2% | 65.6% | 66.8% | 0.6 | 64.8% | 66.9% | 1.4 |
| Prey Veng | 66.0% | 66.2% | 66.2% | 0.5 | 81.1% | 66.6% | 15.1 |
| Pursat | 76.4% | 74.3% | 75.2% | 0.9 | 65.9% | 75.5% | 10.5 |
| Siem Reap | 76.4% | 76.7% | 73.8% | 2.4 | 73.2% | 74.0% | 3.2 |
| Svay Rieng | 73.3% | 73.3% | 73.6% | 1.0 | 81.2% | 74.3% | 7.9 |
| Takeo | 77.2% | 77.5% | 78.1% | 0.7 | 82.2% | 76.5% | 5.0 |
| | | | **Mean Diff.** | **1.3** | | **Mean Diff.** | **7.2** |
| **Mozambique** | | | | | | | |
| Niassa | 71.0% | 73.3% | 73.2% | 5.1 | 65.5% | 65.9% | 5.5 |
| Cabo Delgado | 73.4% | 73.0% | 71.7% | 1.2 | 64.1% | 72.2% | 9.3 |
| Nampula | 70.6% | 70.1% | 73.1% | 0.7 | 57.7% | 69.9% | 12.9 |
| Zambezia | 55.3% | 56.1% | 56.3% | 1.4 | 57.5% | 53.9% | 2.2 |
| Tete | 64.3% | 65.7% | 64.8% | 1.5 | 57.7% | 65.8% | 6.6 |
| Manica | 71.5% | 71.6% | 72.5% | 1.3 | 59.6% | 70.2% | 11.9 |
| Sofala | 73.6% | 74.1% | 75.2% | 1.3 | 63.2% | 72.3% | 10.4 |
| Inhambane | 77.4% | 75.9% | 77.2% | 2.1 | 64.7% | 75.3% | 12.7 |
| Gaza | 77.4% | 75.1% | 73.9% | 3.1 | 62.8% | 74.4% | 14.6 |
| Maputo | 81.0% | 81.0% | 81.6% | 9.8 | 65.6% | 71.2% | 15.5 |
| Maputo City | 81.7% | 81.9% | 80.3% | 8.9 | 73.7% | 72.8% | 8.0 |
| | | | **Mean Diff.** | **3.3** | | **Mean Diff.** | **12.2** |

**Supplementary Table 3.** Association of the difference between *predicted* and *direct* estimates with settlement area and number of clusters removed from estimation sample. Estimates from a generalized linear model, adjusted for survey and interaction between settlement area and number of cluster units removed from estimation sample.

| Indicator | exp(b) | std. err. | z | p-value | 95% CI | |
|---|---|---|---|---|---|---|
| | | | | | LB | UB |
| Access to Electricity | | | | | | |
| Clust. excl.(log) | 1.008 | 0.011 | 0.750 | 0.456 | 0.987 | 1.030 |
| Set. size(log) | 0.998 | 0.000 | -4.380 | 0.000 | 0.998 | 0.999 |
| Access to Improved Water Source | | | | | | |
| Clust. excl.(log) | 0.990 | 0.012 | -0.890 | 0.375 | 0.967 | 1.013 |
| Set. size(log) | 0.997 | 0.000 | -6.610 | 0.000 | 0.996 | 0.998 |
| Iodized Salt Intake in Household | | | | | | |
| Clust. excl.(log) | 1.034 | 0.007 | 4.780 | 0.000 | 1.020 | 1.049 |
| Set. size(log) | 1.000 | 0.000 | -1.350 | 0.176 | 0.999 | 1.000 |
| Prevalence of any Anemia in Women | | | | | | |
| Clust. excl.(log) | 0.999 | 0.005 | -0.230 | 0.820 | 0.988 | 1.010 |
| Set. size(log) | 0.999 | 0.000 | -3.550 | 0.000 | 0.999 | 1.000 |
| Antenatal Care Visits (4+) | | | | | | |
| Clust. excl.(log) | 1.006 | 0.009 | 0.660 | 0.511 | 0.989 | 1.023 |
| Set. size(log) | 0.999 | 0.000 | -4.860 | 0.000 | 0.998 | 0.999 |
| Low Birth Weight Prevalence | | | | | | |
| Clust. excl.(log) | 0.986 | 0.008 | -1.710 | 0.088 | 0.970 | 1.002 |
| Set. size(log) | 0.999 | 0.000 | -4.240 | 0.000 | 0.998 | 0.999 |
| Exclusive Breastfeeding (0-6 months) | | | | | | |
| Clust. excl.(log) | 1.008 | 0.015 | 0.560 | 0.575 | 0.980 | 1.037 |
| Set. size(log) | 0.998 | 0.000 | -4.970 | 0.000 | 0.997 | 0.999 |
| BCG Immunization (12-23 months) | | | | | | |
| Clust. excl.(log) | 0.974 | 0.010 | -2.650 | 0.008 | 0.955 | 0.993 |
| Set. size(log) | 0.997 | 0.000 | -7.480 | 0.000 | 0.997 | 0.998 |
| DPT3 Immunization (12-23 months) | | | | | | |
| Clust. excl.(log) | 0.992 | 0.011 | -0.740 | 0.461 | 0.972 | 1.013 |
| Set. size(log) | 0.997 | 0.000 | -7.000 | 0.000 | 0.997 | 0.998 |
| Measles Immunization (12-23 months) | | | | | | |
| Clust. excl.(log) | 0.978 | 0.011 | -2.090 | 0.036 | 0.957 | 0.999 |
| Set. size(log) | 0.998 | 0.000 | -6.420 | 0.000 | 0.997 | 0.998 |
| Use of Bed Nets; Children (0-59 months) | | | | | | |
| Clust. excl.(log) | 0.991 | 0.007 | -1.230 | 0.217 | 0.978 | 1.005 |
| Set. size(log) | 0.999 | 0.000 | -4.970 | 0.000 | 0.998 | 0.999 |
| Diarrhea Treatment w/ ORS (0-59 months) | | | | | | |
| Clust. excl.(log) | 0.975 | 0.013 | -1.880 | 0.060 | 0.950 | 1.001 |
| Set. size(log) | 0.998 | 0.000 | -4.180 | 0.000 | 0.997 | 0.999 |
| Stunting Prevalence (0-59 months) | | | | | | |
| Clust. excl.(log) | 1.012 | 0.005 | 2.210 | 0.027 | 1.001 | 1.023 |
| Set. size(log) | 0.999 | 0.000 | -3.830 | 0.000 | 0.999 | 1.000 |
| Wasting Prevalence (0-59 months) | | | | | | |
| Clust. excl.(log) | 1.009 | 0.003 | 2.660 | 0.008 | 1.002 | 1.016 |
| Set. size(log) | 0.999 | 0.000 | -4.930 | 0.000 | 0.999 | 1.000 |
| Prev. of Anemia; Children (6-59 months) | | | | | | |
| Clust. excl.(log) | 1.031 | 0.008 | 4.010 | 0.000 | 1.016 | 1.047 |
| Set. size(log) | 0.999 | 0.000 | -3.140 | 0.002 | 0.999 | 1.000 |

**Supplementary Table 4.** K-Fold validation results. Bias of the model predictions for each settlement across the 10 validation countries for each of the 15 indicators assessed.

| Indicator | Angola-2015 | Benin-2017 | Cambodia-2014 | Gabon-2012 | Malawi-2015 | Mali-2018 | Mozambique-2011 | Nigeria-2013 | Senegal-2019 | Zambia-2018 |
|---|---|---|---|---|---|---|---|---|---|---|
| **Bias** | | | | | | | | | | |
| Access to Electricity | 0.004 | 0.004 | 0.007 | 0.004 | 0.003 | 0.003 | 0.003 | 0.003 | 0.007 | 0.004 |
| Improved Water Source | 0.008 | 0.001 | 0.002 | 0.007 | 0.005 | 0.005 | 0.001 | 0.001 | 0.006 | 0.004 |
| Iodized Salt Intake | -0.001 | -0.001 | -0.001 | 0.000 | 0.000 | 0.000 | 0.001 | . | . | 0.001 |
| Prev. of Anemia; Women | . | 0.002 | -0.002 | 0.000 | 0.000 | 0.000 | 0.001 | . | . | 0.000 |
| Antenatal Care (4+) | 0.001 | 0.001 | 0.001 | 0.002 | 0.002 | 0.001 | -0.002 | 0.003 | 0.005 | -0.002 |
| Low Birth Weight Prevalence | -0.001 | 0.000 | -0.002 | -0.001 | 0.001 | 0.003 | 0.001 | -0.002 | 0.001 | 0.001 |
| Exclusive Breastfeeding (0-5m) | -0.001 | 0.002 | -0.003 | -0.001 | 0.002 | -0.003 | 0.005 | 0.004 | 0.004 | -0.004 |
| BCG Immun.(12-23m) | 0.004 | -0.002 | 0.001 | 0.002 | -0.001 | 0.002 | 0.002 | 0.001 | -0.001 | 0.000 |
| DPT3 Immun. (12-23m) | 0.004 | 0.003 | 0.003 | -0.001 | -0.003 | -0.002 | 0.005 | 0.002 | 0.001 | -0.002 |
| Measles Immun. (12-23m) | 0.003 | 0.001 | 0.004 | -0.003 | 0.000 | 0.001 | 0.000 | 0.002 | 0.001 | 0.001 |
| Bed Net Use; Children (0-59m) | 0.001 | 0.000 | . | 0.002 | 0.003 | 0.000 | 0.001 | 0.001 | 0.000 | -0.001 |
| Diarrhea Treat.; ORS (0-59m) | 0.004 | 0.000 | -0.002 | 0.001 | 0.008 | -0.004 | 0.003 | 0.004 | 0.002 | 0.005 |
| Stunting Prevalence (0-59m) | -0.002 | -0.001 | -0.001 | -0.001 | -0.001 | 0.000 | 0.001 | -0.002 | 0.000 | 0.000 |
| Wasting Prevalence (0-59m) | 0.000 | 0.000 | 0.000 | 0.001 | 0.000 | -0.002 | 0.000 | 0.001 | 0.001 | 0.000 |
| Anemia; Children (6-59m) | -0.003 | -0.002 | -0.002 | -0.004 | -0.001 | -0.001 | -0.002 | . | . | 0.001 |

**Supplementary Table 5.** K-Fold validation results; Mean Absolute Deviance (MAD) of the model predictions for each settlement across the 10 validation countries for each of the 15 indicators assessed.

| Indicator | Angola-2015 | Benin-2017 | Cambodia-2014 | Gabon-2012 | Malawi-2015 | Mali-2018 | Mozambique-2011 | Nigeria-2013 | Senegal-2019 | Zambia-2018 |
|---|---|---|---|---|---|---|---|---|---|---|
| **MAD** | | | | | | | | | | |
| Access to Electricity | 0.010 | 0.012 | 0.019 | 0.020 | 0.007 | 0.012 | 0.008 | 0.019 | 0.021 | 0.012 |
| Improved Water Source | 0.019 | 0.014 | 0.013 | 0.012 | 0.013 | 0.016 | 0.017 | 0.017 | 0.015 | 0.015 |
| Iodized Salt Intake | 0.008 | 0.003 | 0.009 | 0.002 | 0.008 | 0.006 | 0.008 | . | . | 0.010 |
| Prev. of Anemia; Women | . | 0.010 | 0.007 | 0.009 | 0.011 | 0.008 | 0.008 | . | . | 0.006 |
| Antenatal Care (4+) | 0.012 | 0.012 | 0.013 | 0.013 | 0.010 | 0.011 | 0.013 | 0.014 | 0.013 | 0.011 |
| Low Birth Weight Prevalence | 0.008 | 0.006 | 0.005 | 0.007 | 0.004 | 0.011 | 0.010 | 0.014 | 0.005 | 0.004 |
| Exclusive Breastfeeding (0-5m) | 0.016 | 0.015 | 0.022 | 0.009 | 0.021 | 0.018 | 0.019 | 0.012 | 0.018 | 0.017 |
| BCG Immun. (12-23m) | 0.017 | 0.011 | 0.007 | 0.011 | 0.004 | 0.014 | 0.009 | 0.012 | 0.005 | 0.005 |
| DPT3 Immun. (12-23m) | 0.015 | 0.012 | 0.013 | 0.015 | 0.006 | 0.014 | 0.014 | 0.013 | 0.007 | 0.008 |
| Measles Immun. (12-23m) | 0.017 | 0.014 | 0.016 | 0.016 | 0.008 | 0.013 | 0.014 | 0.013 | 0.009 | 0.010 |
| Bed Net Use; Children (0-59m) | 0.009 | 0.008 | . | 0.009 | 0.009 | 0.006 | 0.009 | 0.007 | 0.012 | 0.009 |
| Diarrhea Treat.; ORS (0-59m) | 0.016 | 0.015 | 0.017 | 0.015 | 0.018 | 0.014 | 0.022 | 0.015 | 0.017 | 0.019 |
| Stunting Prevalence (0-59m) | 0.008 | 0.007 | 0.010 | 0.008 | 0.012 | 0.006 | 0.007 | 0.007 | 0.006 | 0.008 |
| Wasting Prevalence (0-59m) | 0.004 | 0.002 | 0.006 | 0.003 | 0.002 | 0.004 | 0.003 | 0.005 | 0.004 | 0.003 |
| Anemia; Children (6-59m) | 0.013 | 0.010 | 0.011 | 0.013 | 0.012 | 0.007 | 0.011 | . | . | 0.008 |

### Additional Figures

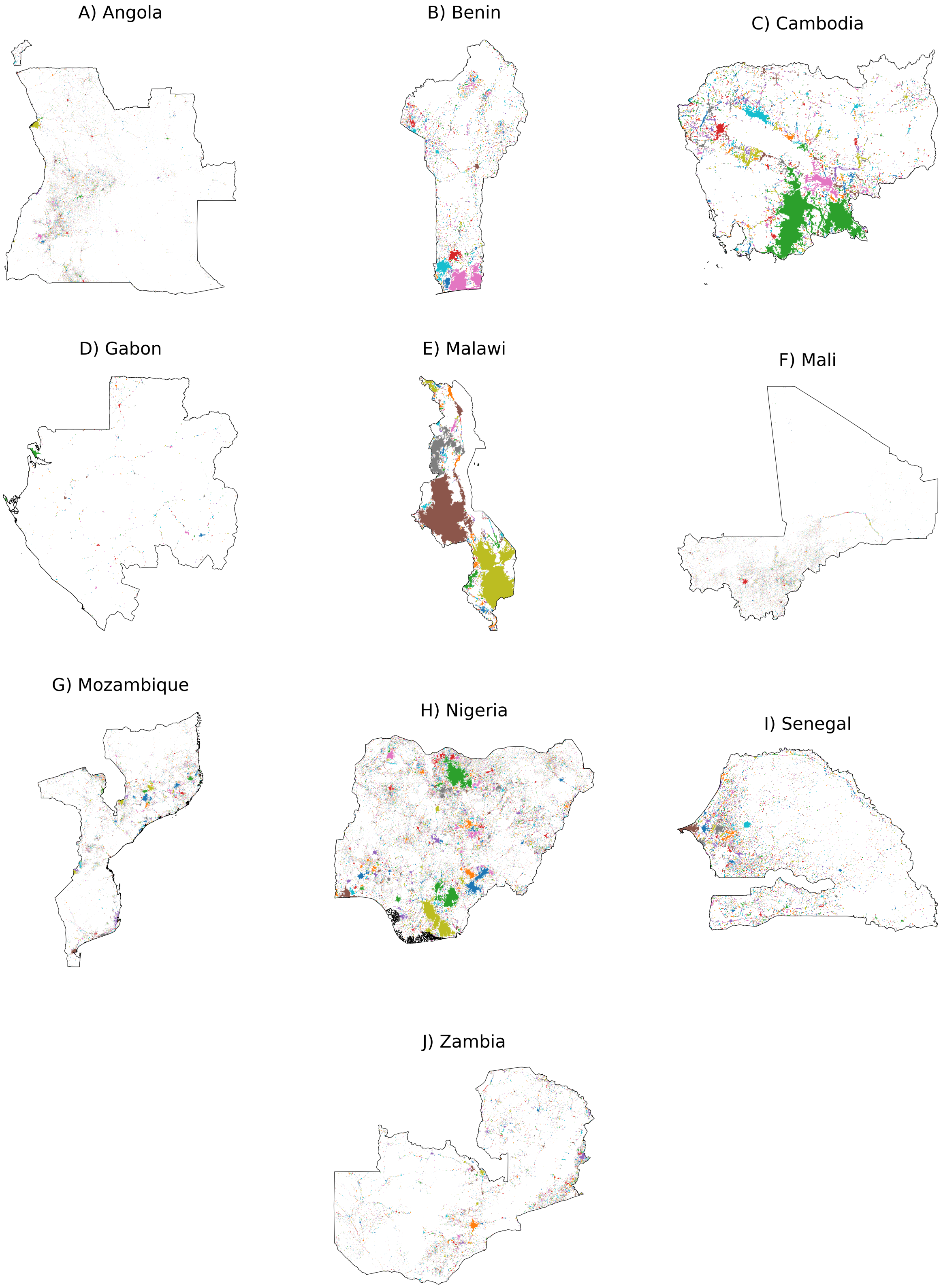


**Supplementary Figure 1.** Settlements mapped for each of the validation countries. Map scales are not uniform for illustrative purposes.

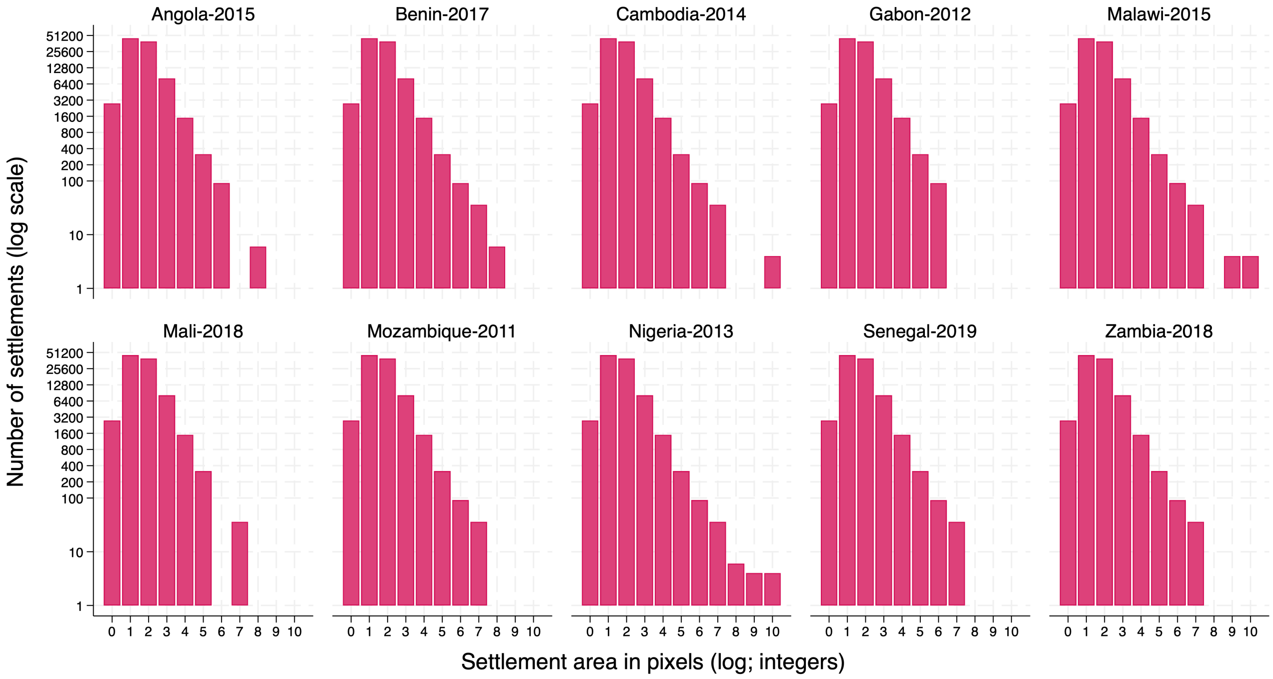


**Supplementary Figure 2.** Distribution of the settlements (n=101,435) across all 10 surveys included in the technical validation by area size. Logarithmic scale.

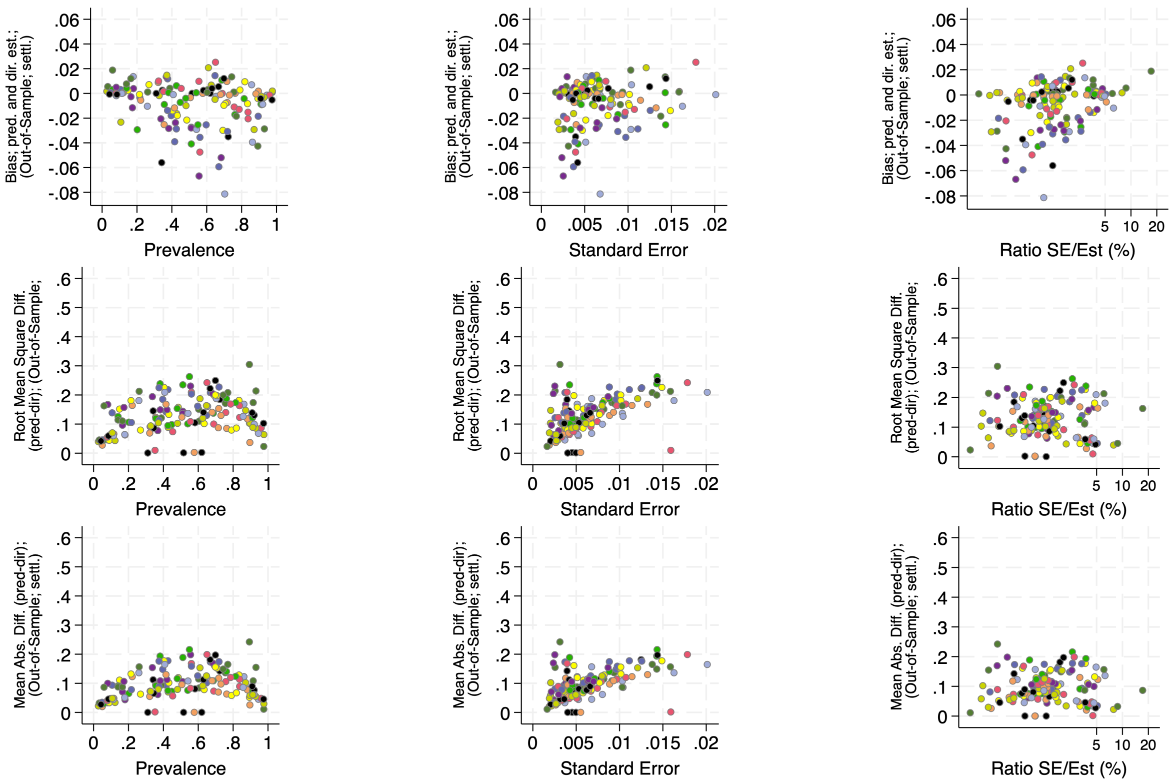


**Legend:**

- Angola - 2015
- Benin - 2017
- Cambodia - 2014
- Gabon - 2012
- Malawi - 2015
- Mali - 2018
- Mozambique - 2011
- Nigeria - 2013
- Senegal - 2019
- Zambia - 2018

**Supplementary Figure 3.** Validation metrics (Bias, Root Mean Square Difference and Mean Absolute Difference of LIDW *predictions* and DHS *direct* estimates) by indicator prevalence, standard error and relative standard error (s.e./indicator value) for the 15 indicators and 10 countries. The color of the markers indicates the DHS survey.

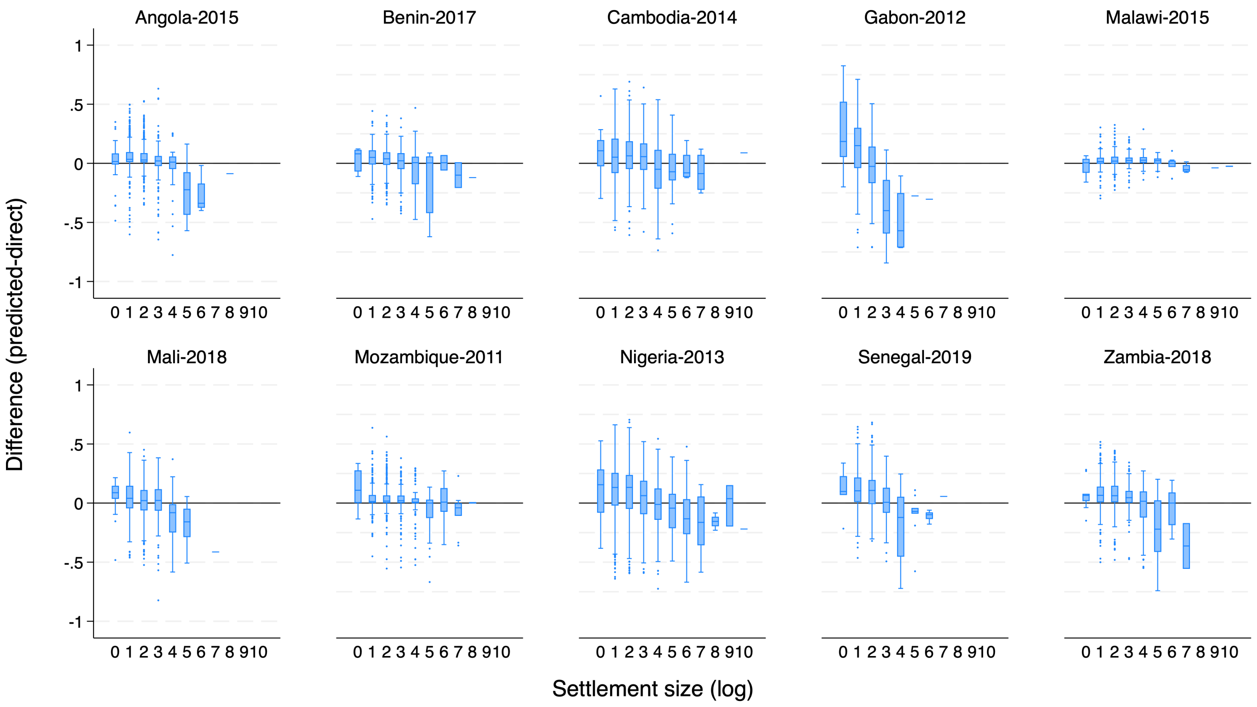


**Supplementary Figure 4.** Difference between LIDW-based Out-Of-Sample *predictions* and DHS *direct* estimates) for the indicator Access to Electricity across the 10 countries by settlement area in pixels (log).

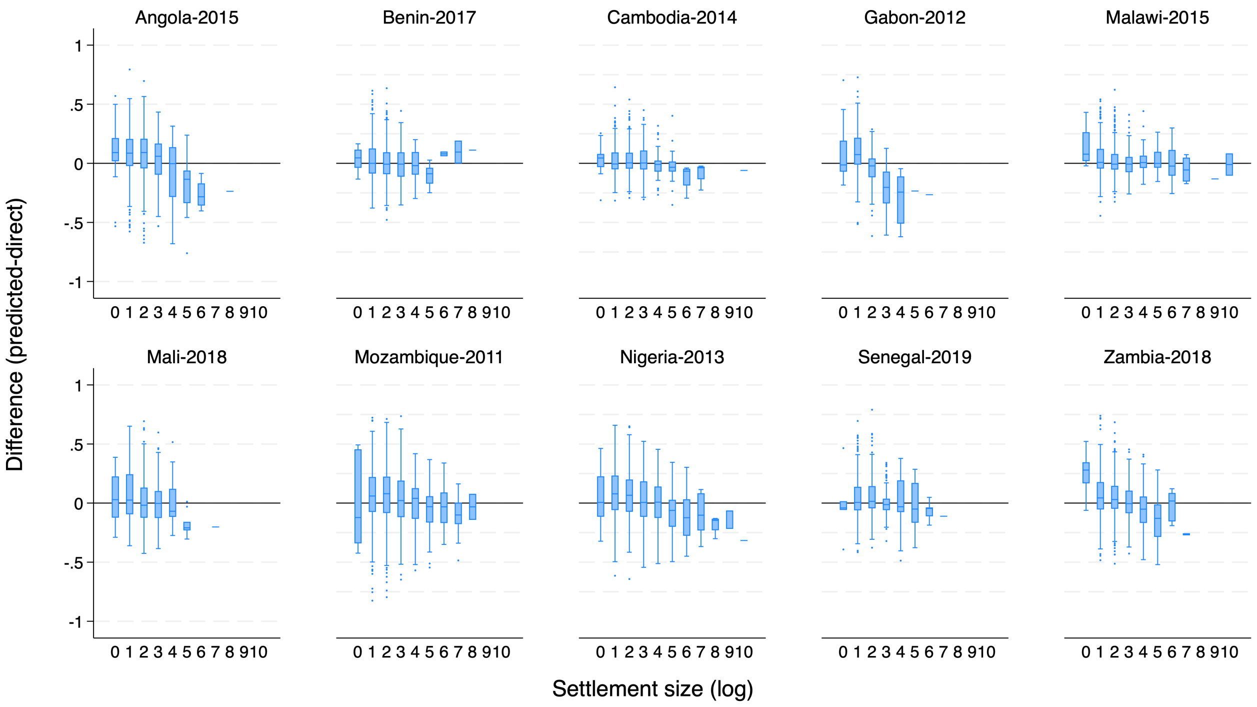


**Supplementary Figure 5.** Difference between LIDW-based Out-Of-Sample *predictions* and DHS *direct* estimates) for the indicator Access to Improved Water Source across the 10 countries by settlement area in pixels (log).

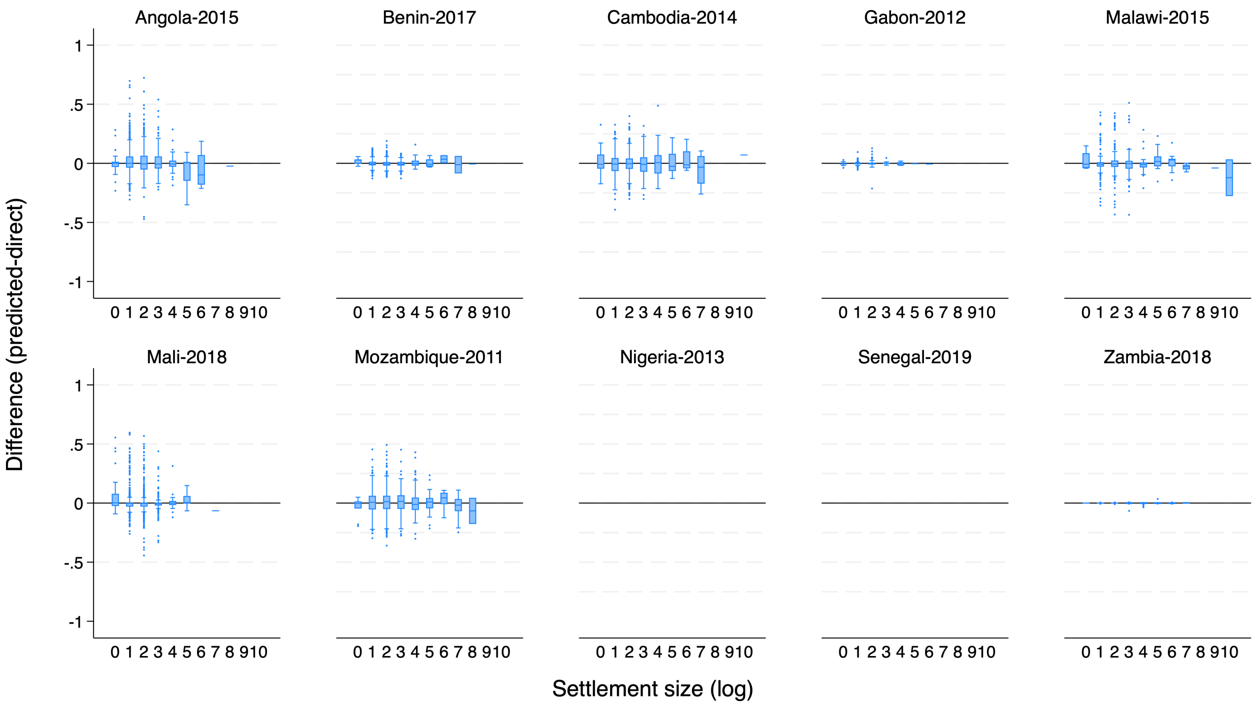


**Supplementary Figure 6.** Difference between LIDW-based Out-Of-Sample *predictions* and DHS *direct* estimates) for the indicator Iodized Salt Intake in the Household across the 10 countries by settlement area in pixels (log).

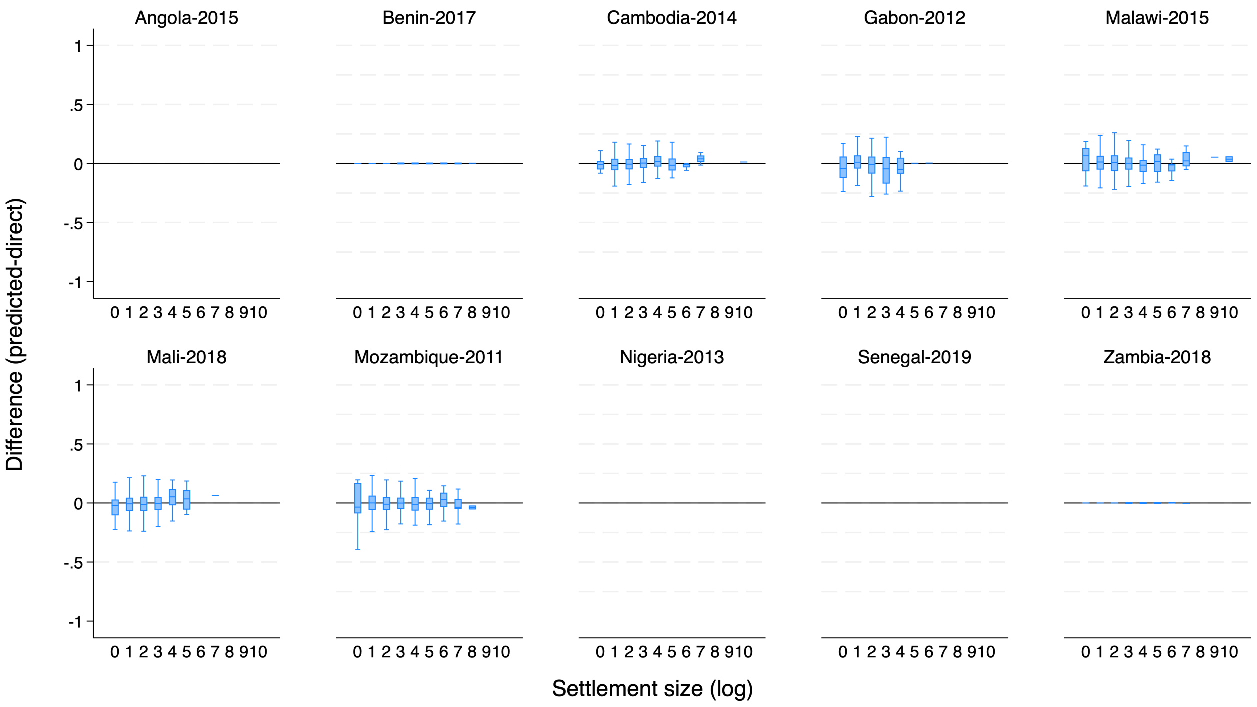


**Supplementary Figure 7.** Difference between LIDW-based Out-Of-Sample *predictions* and DHS *direct* estimates) for the indicator Prevalence of Anemia in Women across the 10 countries by settlement area in pixels (log).

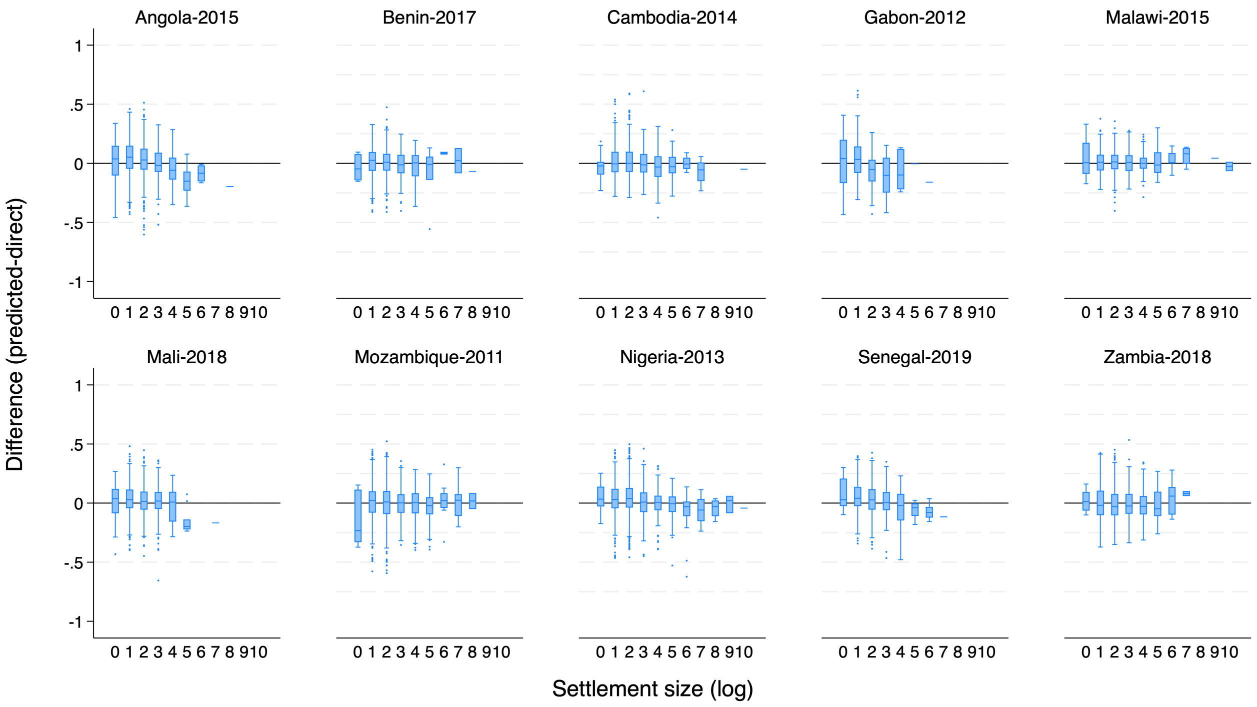


**Supplementary Figure 8.** Difference between LIDW-based Out-Of-Sample *predictions* and DHS *direct* estimates) for the indicator Antenatal Care Visits (4+) during Pregnancy across the 10 countries by settlement area in pixels (log).

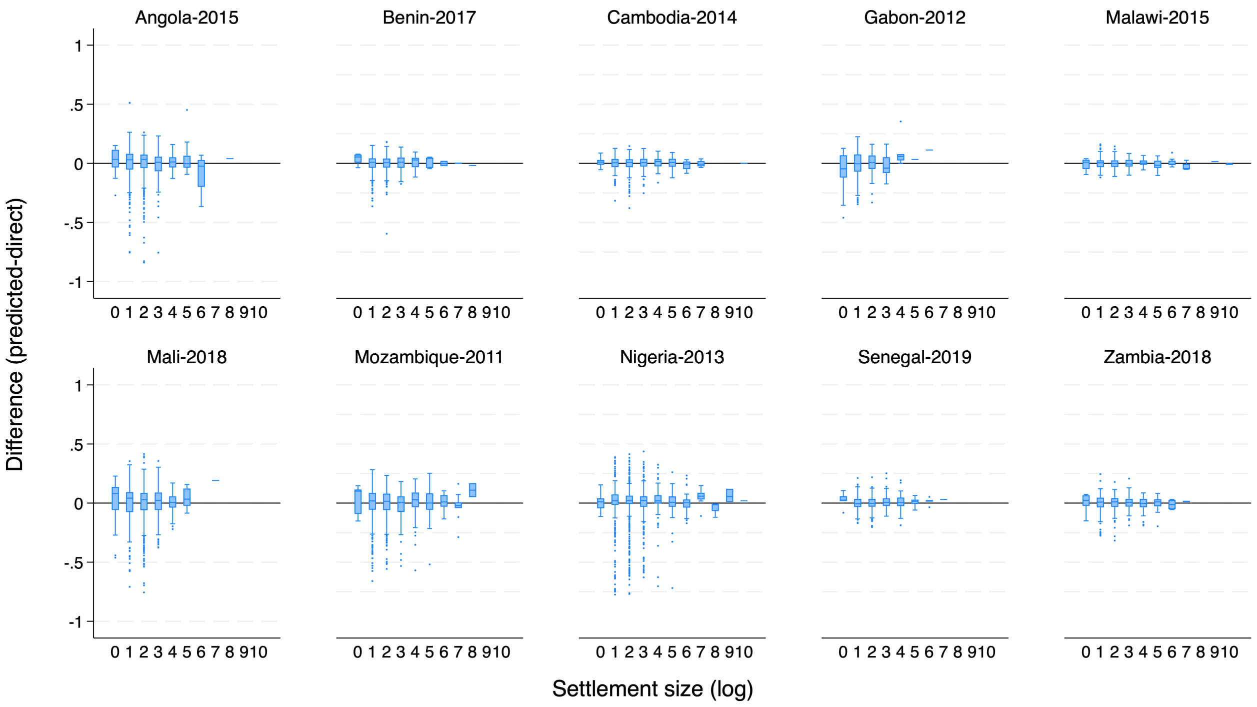


**Supplementary Figure 9.** Difference between LIDW-based Out-Of-Sample *predictions* and DHS *direct* estimates) for the indicator Low Birth Weight Prevalence across the 10 countries by settlement area in pixels (log).

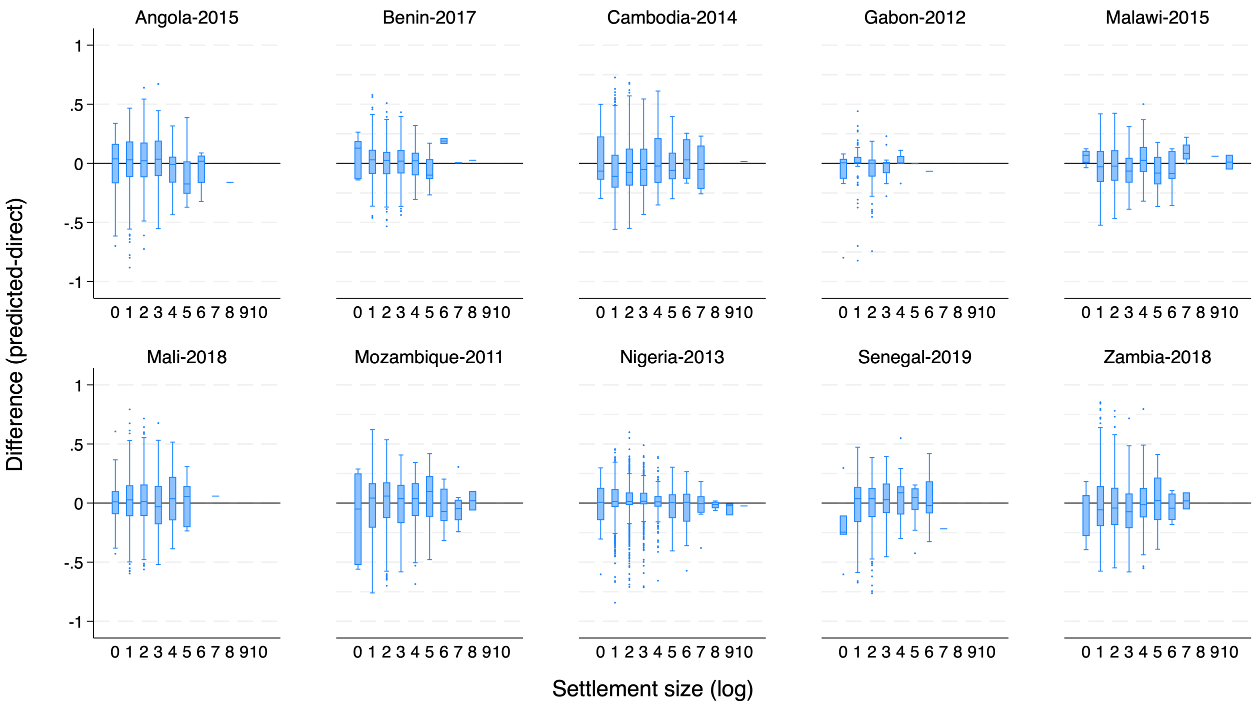


**Supplementary Figure 10.** Difference between LIDW-based Out-Of-Sample *predictions* and DHS *direct* estimates) for the indicator Exclusive Breastfeeding (0-6 months) across the 10 countries by settlement area in pixels (log).

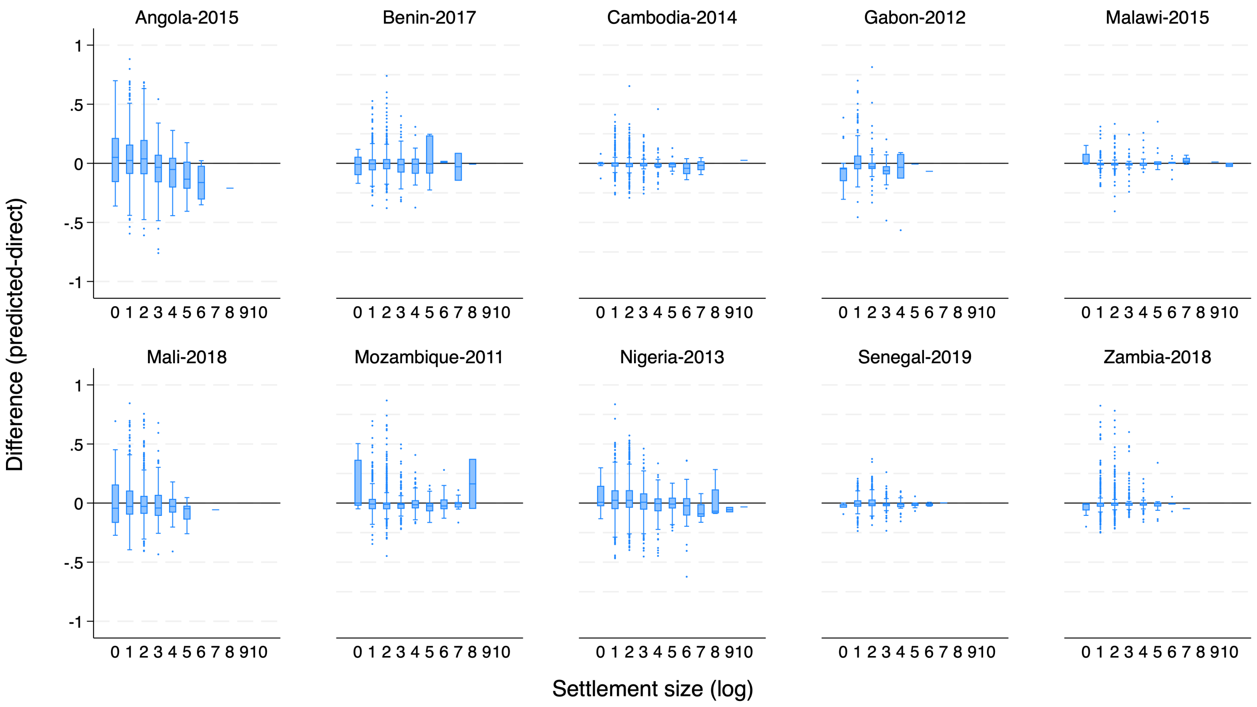


**Supplementary Figure 11.** Difference between LIDW-based Out-Of-Sample *predictions* and DHS *direct* estimates) for the indicator BCG Immunization (12-23 months) across the 10 countries by settlement area in pixels (log).

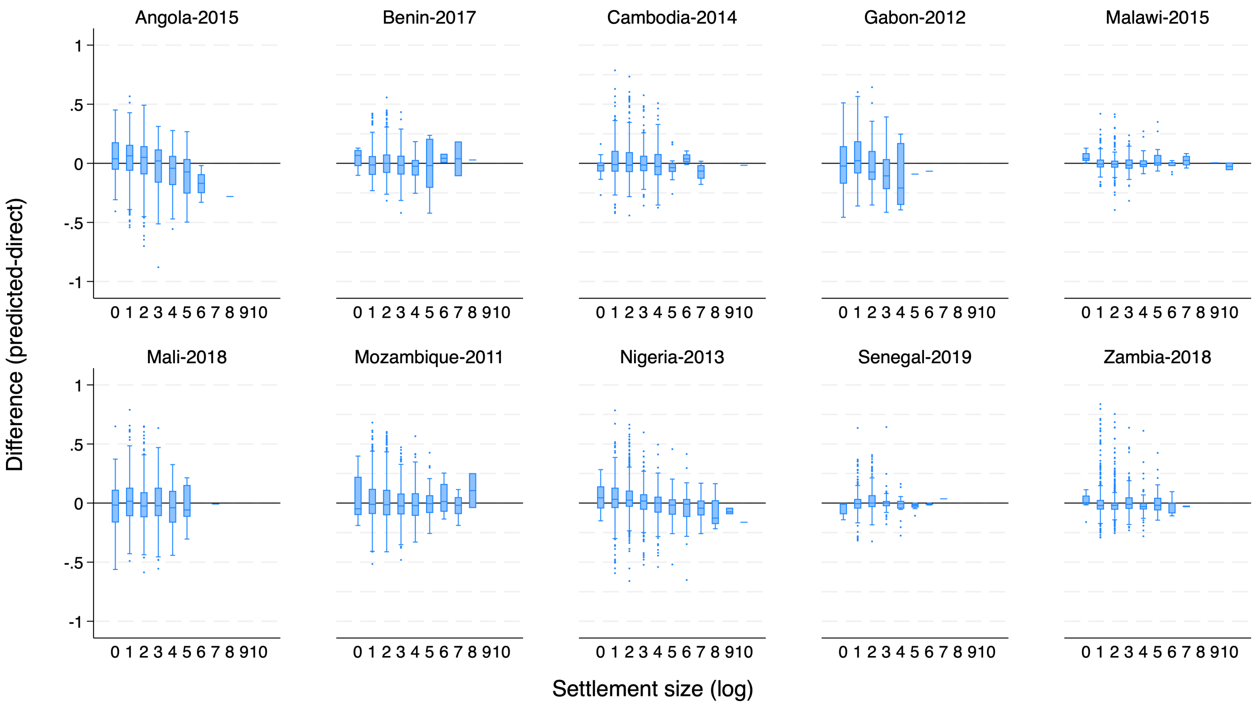


**Supplementary Figure 12.** Difference between LIDW-based Out-Of-Sample *predictions* and DHS *direct* estimates) for the indicator DPT3 Immunization (12-23 months) across the 10 countries by settlement area in pixels (log).

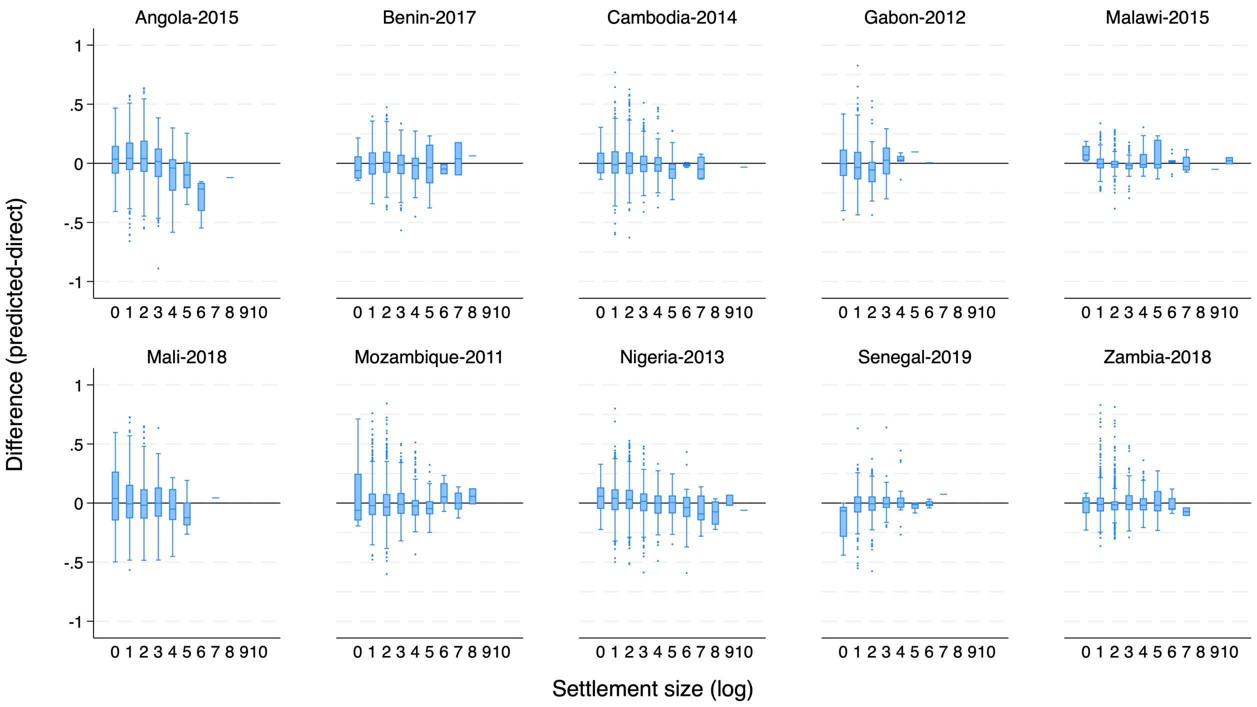


**Supplementary Figure 13.** Difference between LIDW-based Out-Of-Sample *predictions* and DHS *direct* estimates) for the indicator Measles Immunization (12-23 months) across the 10 countries by settlement area in pixels (log).

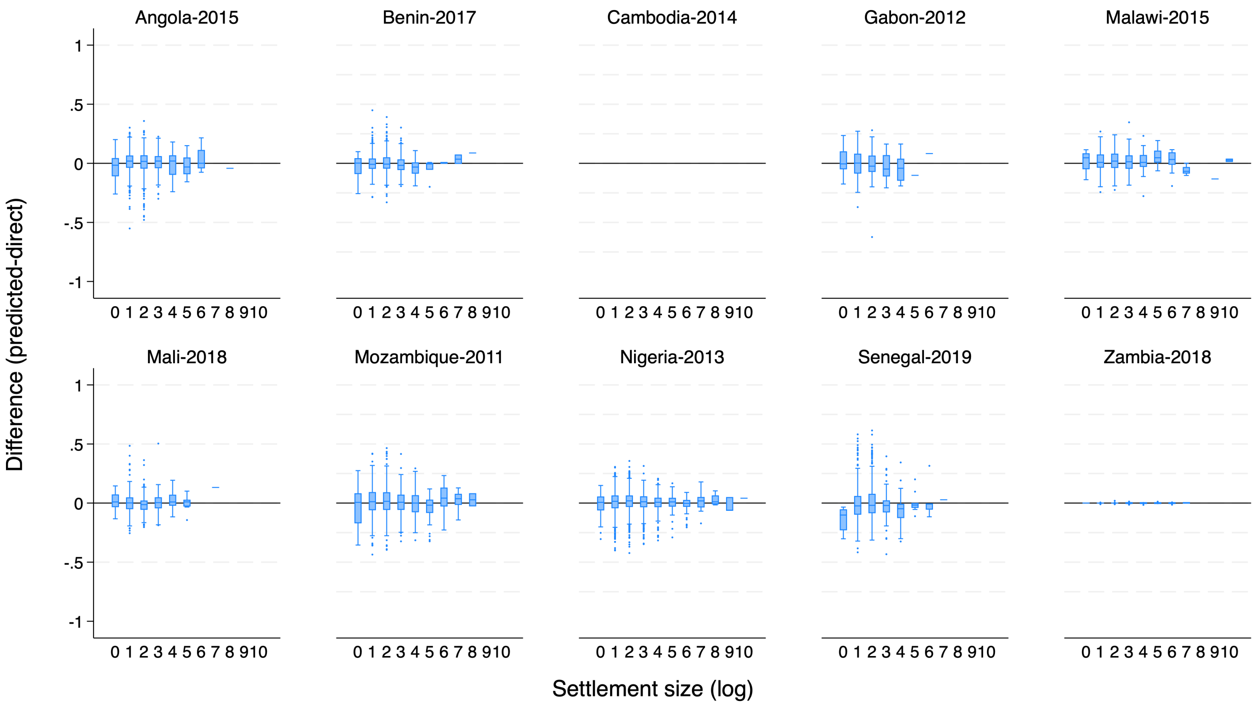


**Supplementary Figure 14.** Difference between LIDW-based Out-Of-Sample *predictions* and DHS *direct* estimates) for the indicator Use of Mosquito Nets among Children (0-59months) across the 10 countries by settlement area in pixels (log).

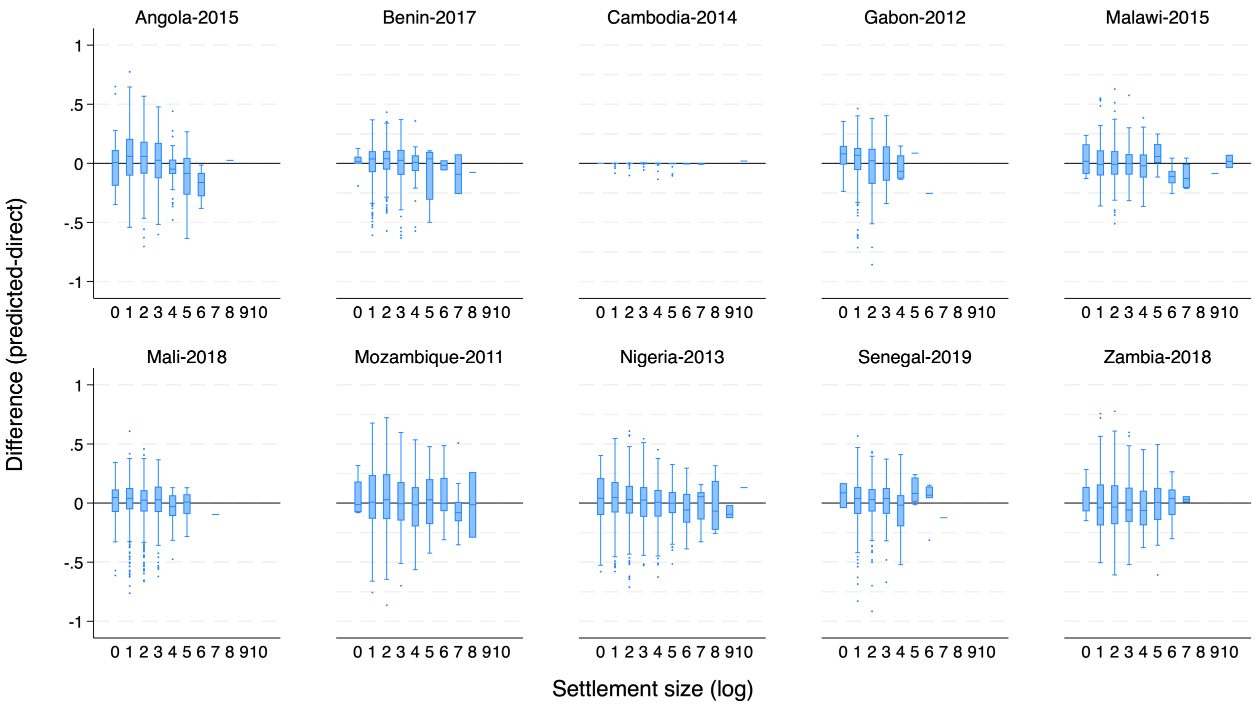


**Supplementary Figure 15.** Difference between LIDW-based Out-Of-Sample *predictions* and DHS *direct* estimates) for the indicator Diarrhea Treatment with ORS among children (0-59 months) across the 10 countries by settlement area in pixels (log).

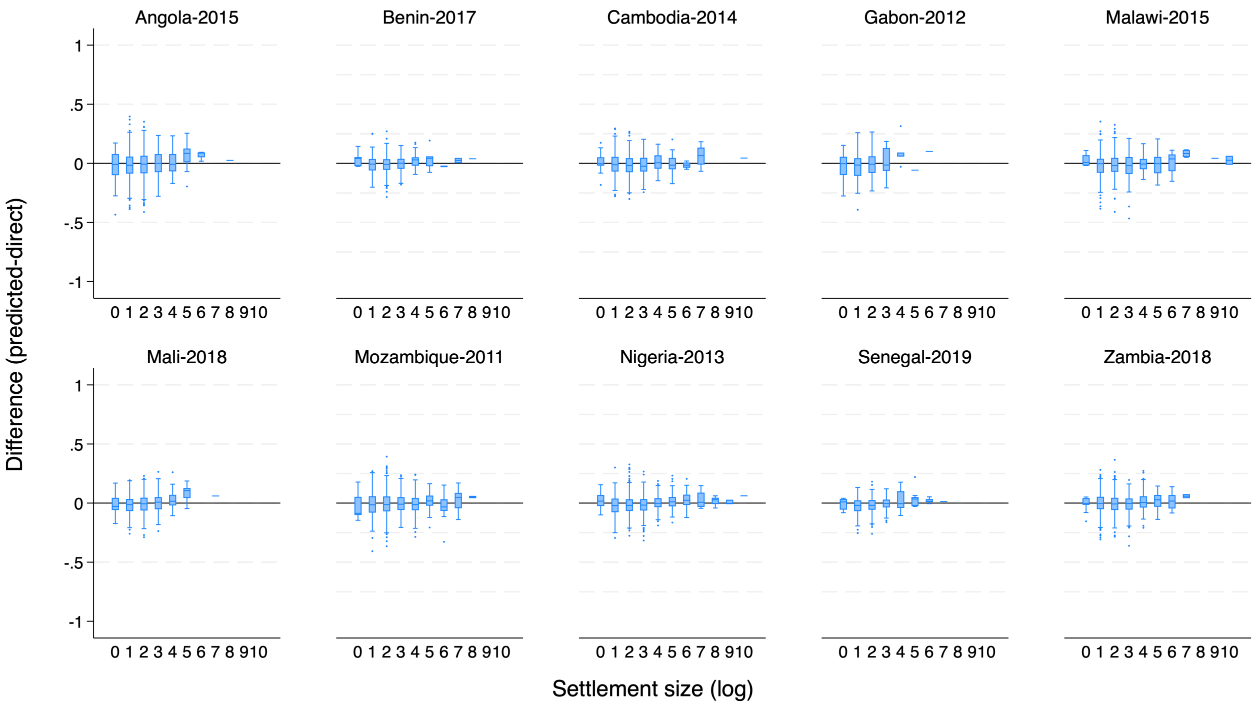


**Supplementary Figure 16.** Difference between LIDW-based Out-Of-Sample *predictions* and DHS *direct* estimates) for the indicator Stunting Prevalence (0-59 months) across the 10 countries by settlement area in pixels (log).

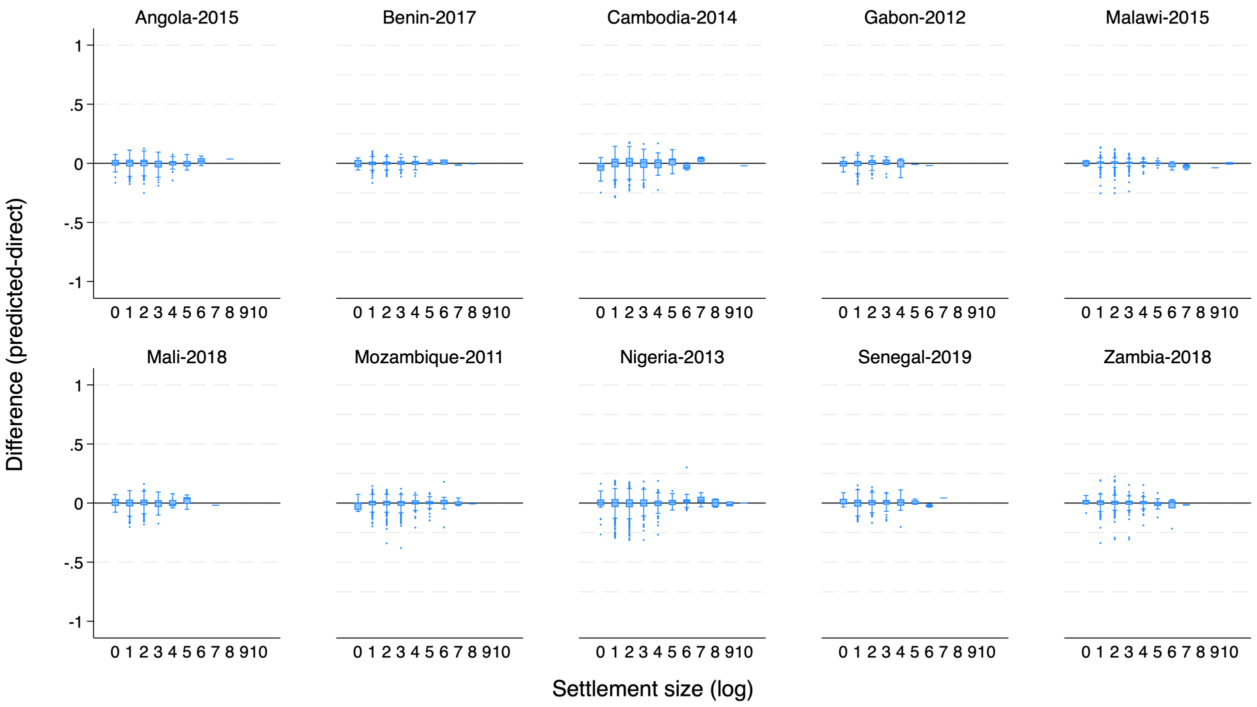


**Supplementary Figure 17.** Difference between LIDW-based Out-Of-Sample *predictions* and DHS *direct* estimates) for the indicator Wasting Prevalence (0-59 months) across the 10 countries by settlement area in pixels (log).

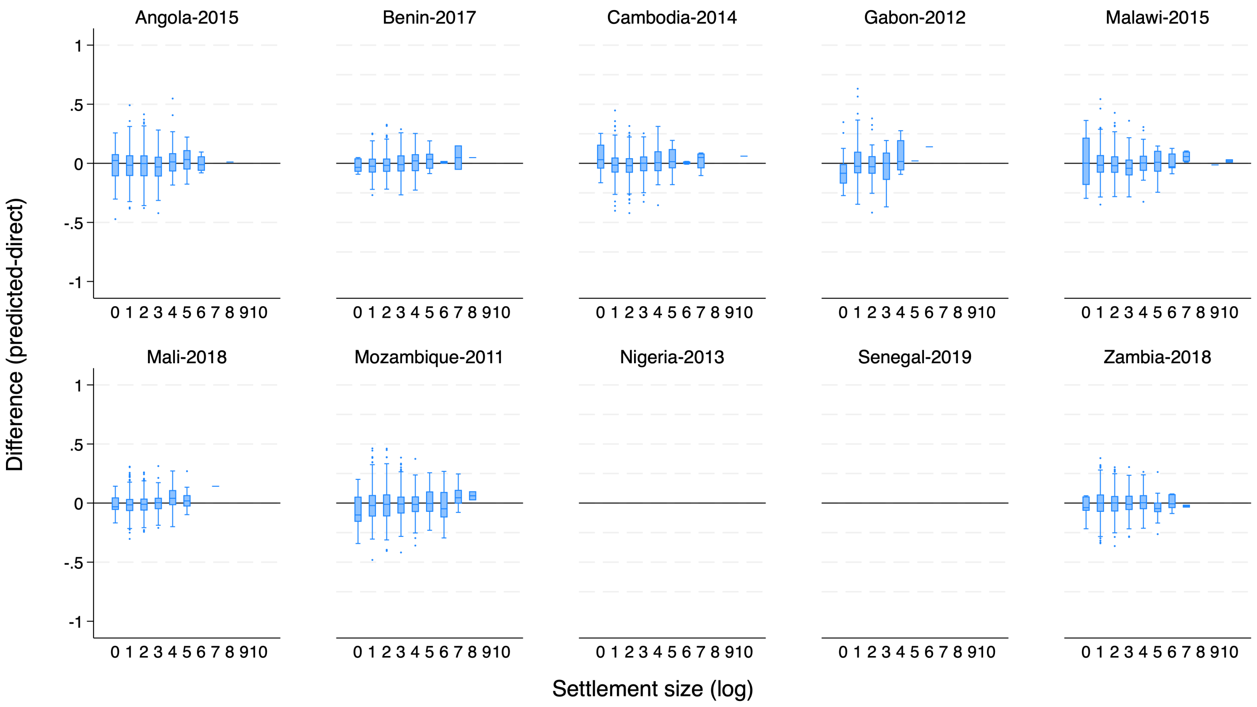


**Supplementary Figure 18.** Difference between LIDW-based Out-Of-Sample *predictions* and DHS *direct* estimates) for the indicator Prevalence of Anemia in Children (6-59 months) across the 10 countries by settlement area in pixels (log).

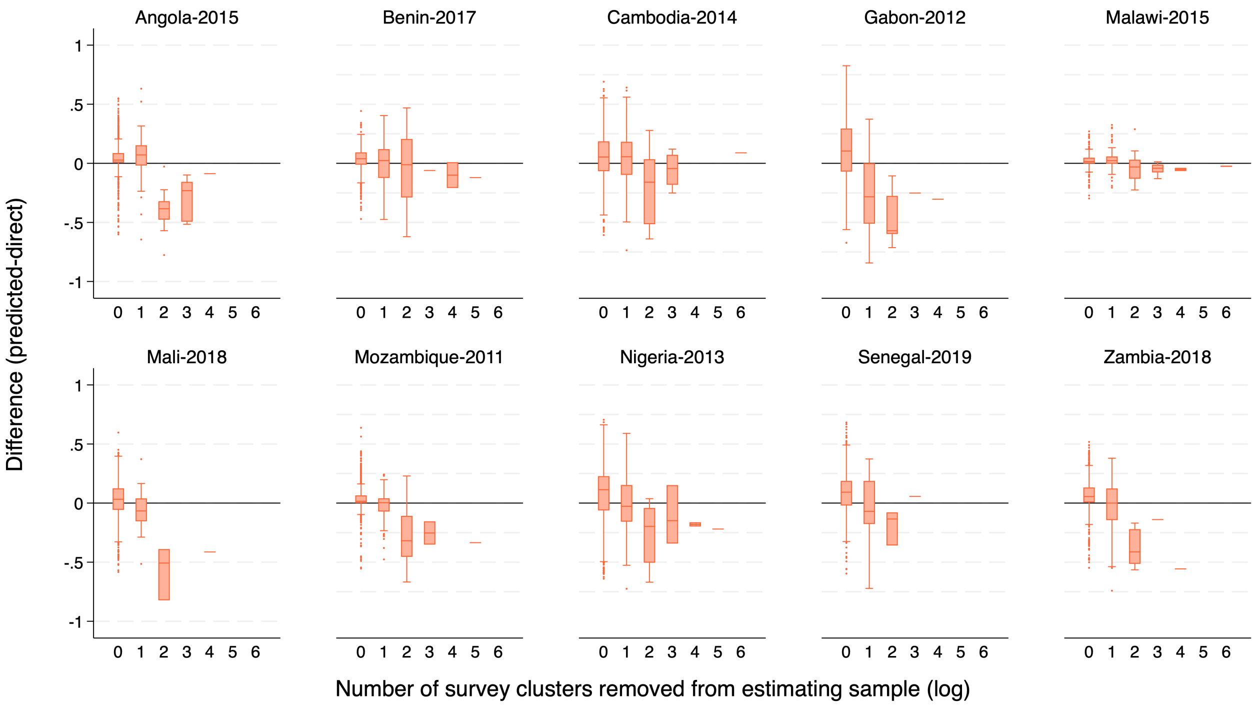


**Supplementary Figure 19.** Difference between LIDW-based Out-Of-Sample *predictions* and DHS *direct* estimates) for the indicator Access to Electricity across the 10 countries by number of *survey clusters* excluded from the estimation sample (log).

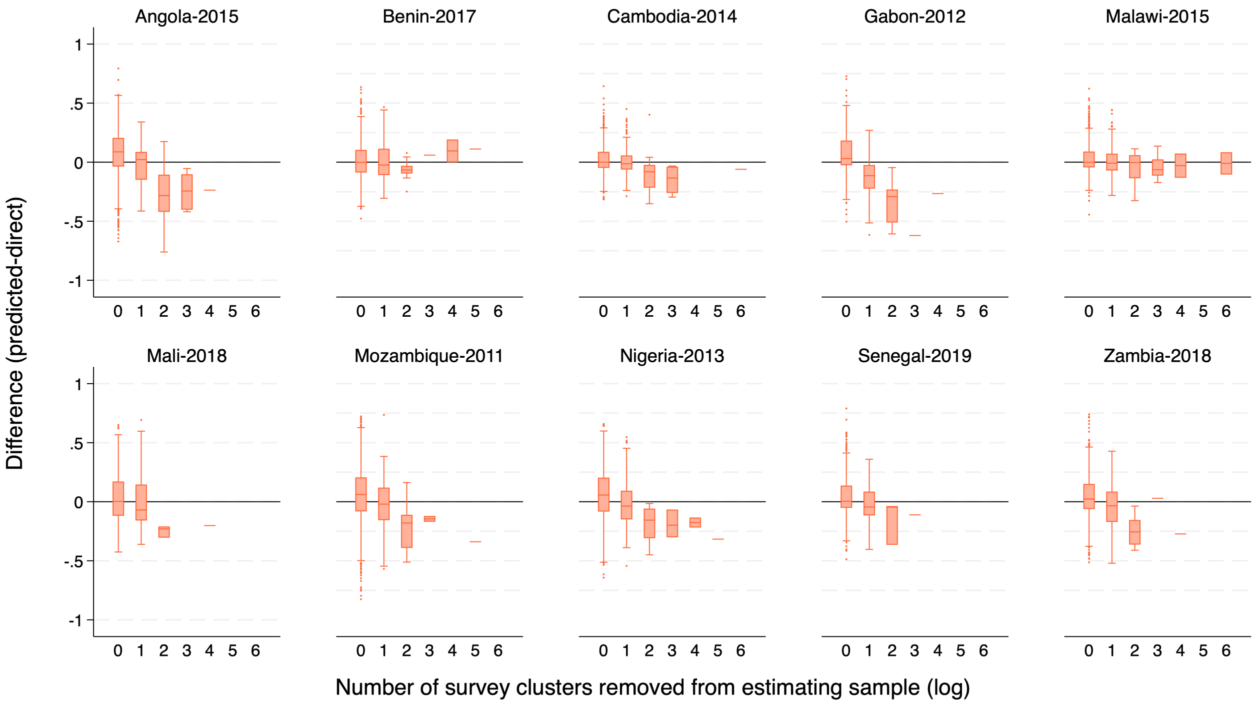


**Supplementary Figure 20.** Difference between LIDW-based Out-Of-Sample *predictions* and DHS *direct* estimates) for the indicator Access to Improved Water Source across the 10 countries by number of *survey clusters* excluded from the estimation sample (log).

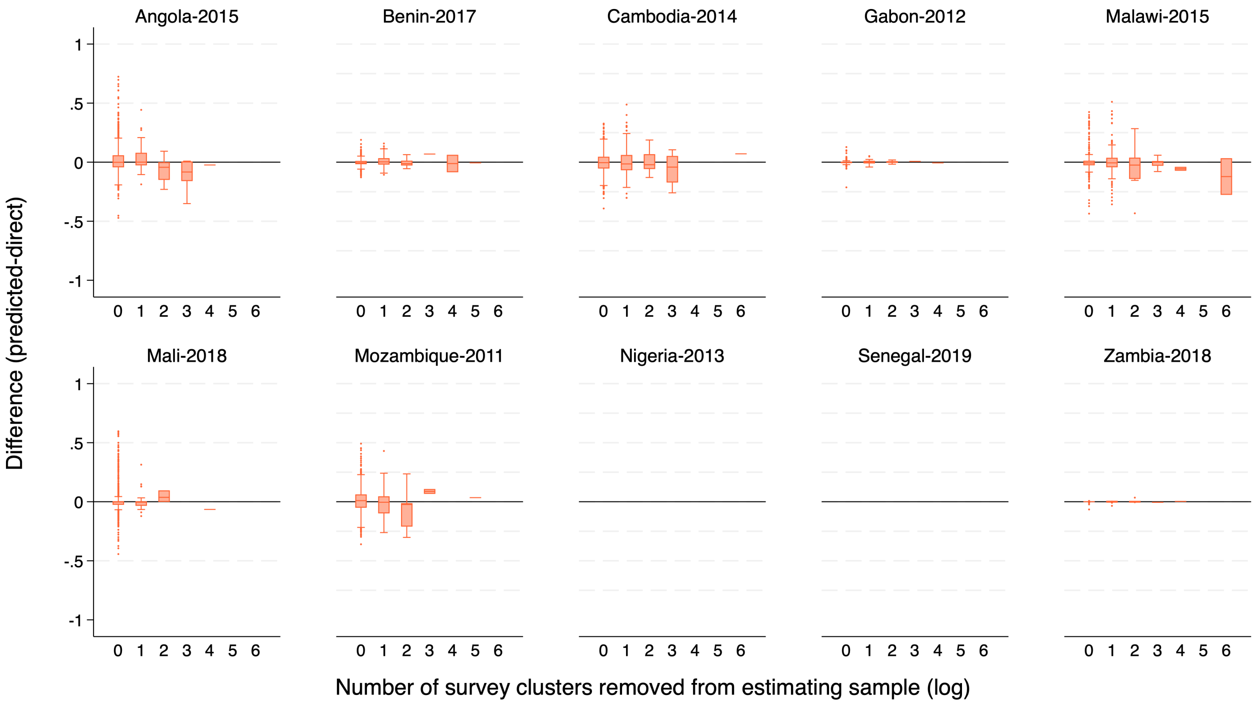


**Supplementary Figure 21.** Difference between LIDW-based Out-Of-Sample *predictions* and DHS *direct* estimates) for the indicator Iodized Salt Intake in Household across the 10 countries by number of *survey clusters* excluded from the estimation sample (log).

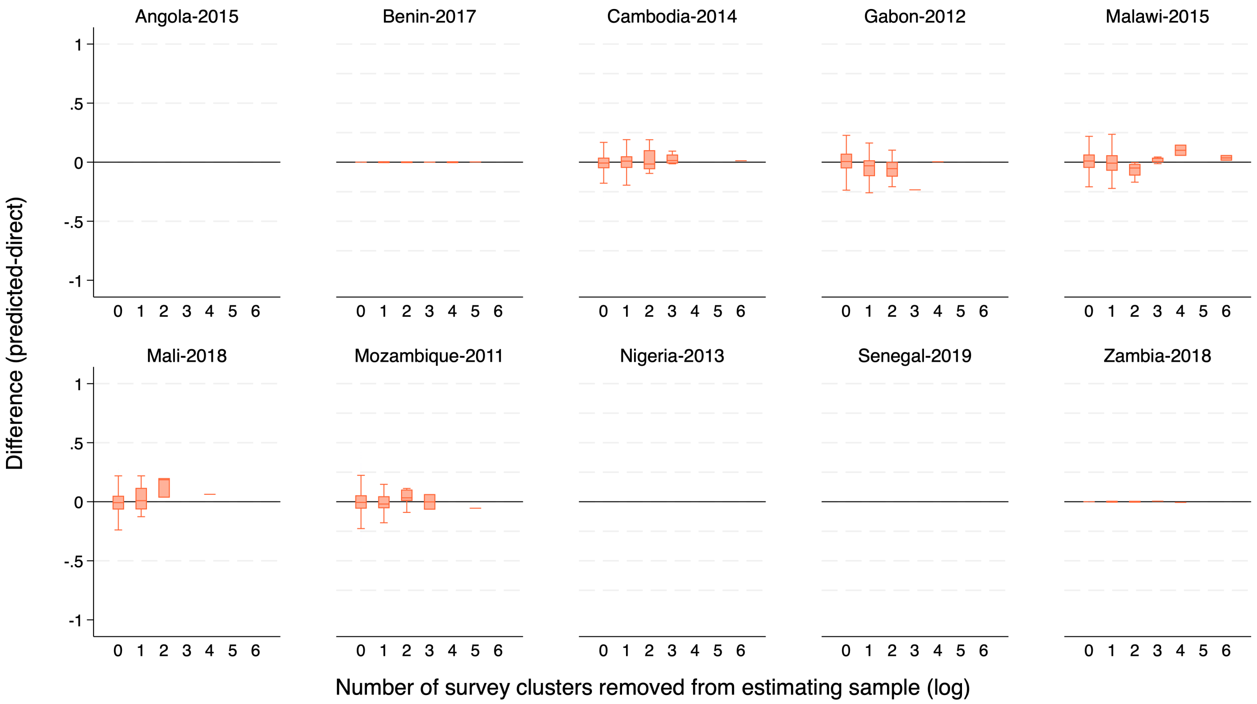


**Supplementary Figure 22.** Difference between LIDW-based Out-Of-Sample *predictions* and DHS *direct* estimates) for the indicator Prevalence of any Anemia in Women across the 10 countries by number of *survey clusters* excluded from the estimation sample (log).

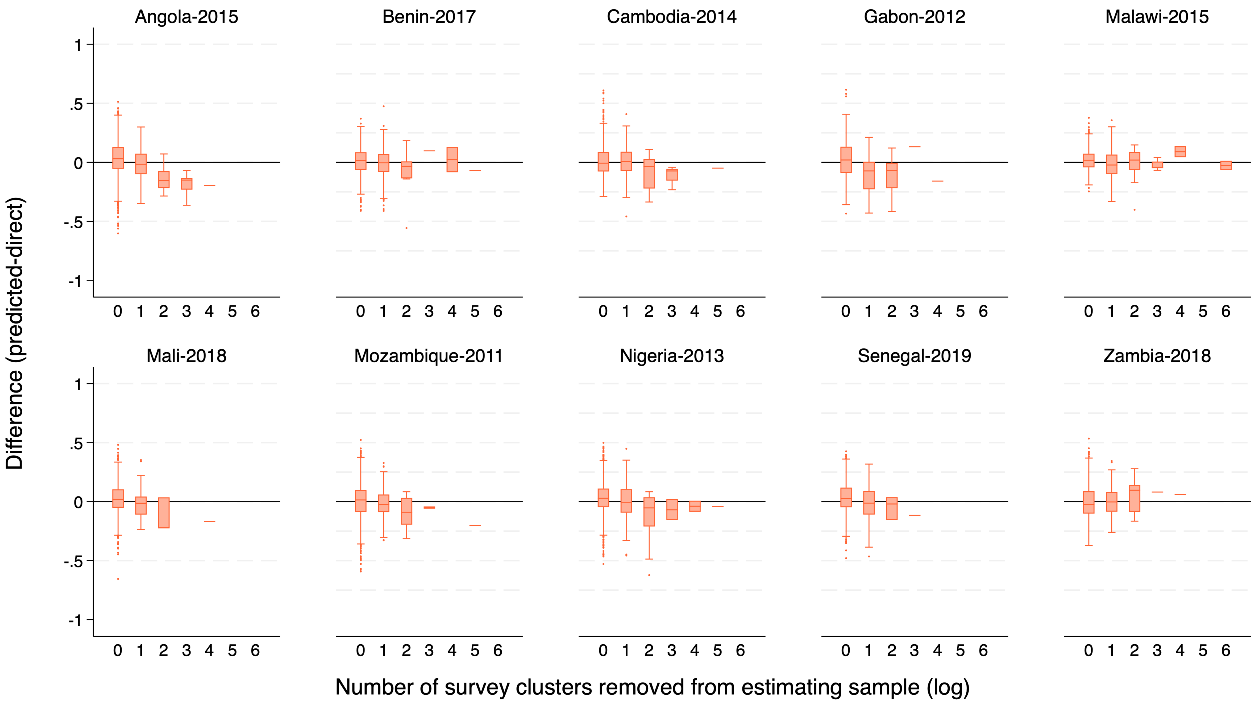


**Supplementary Figure 23.** Difference between LIDW-based Out-Of-Sample *predictions* and DHS *direct* estimates) for the indicator Antenatal Care Visits (4+) during Pregnancy across the 10 countries by number of *survey clusters* excluded from the estimation sample (log).

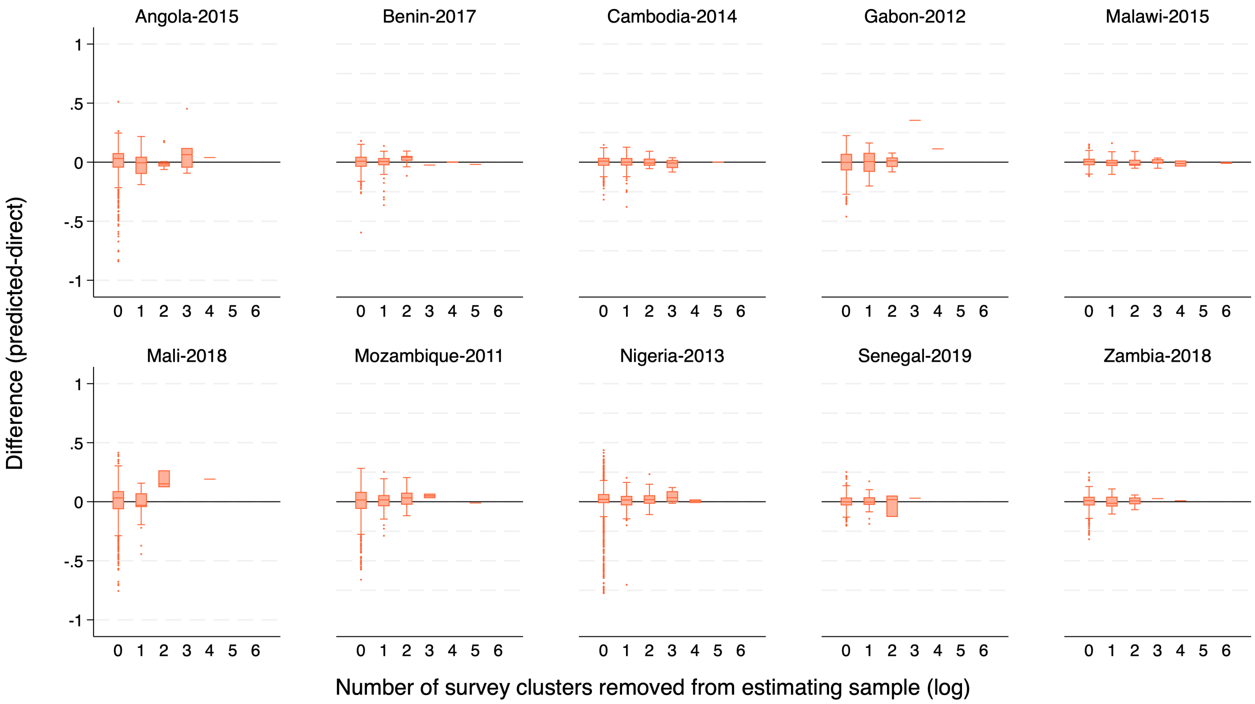


**Supplementary Figure 24.** Difference between LIDW-based Out-Of-Sample *predictions* and DHS *direct* estimates) for the indicator Low Birth Weight Prevalence across the 10 countries by number of *survey clusters* excluded from the estimation sample (log).

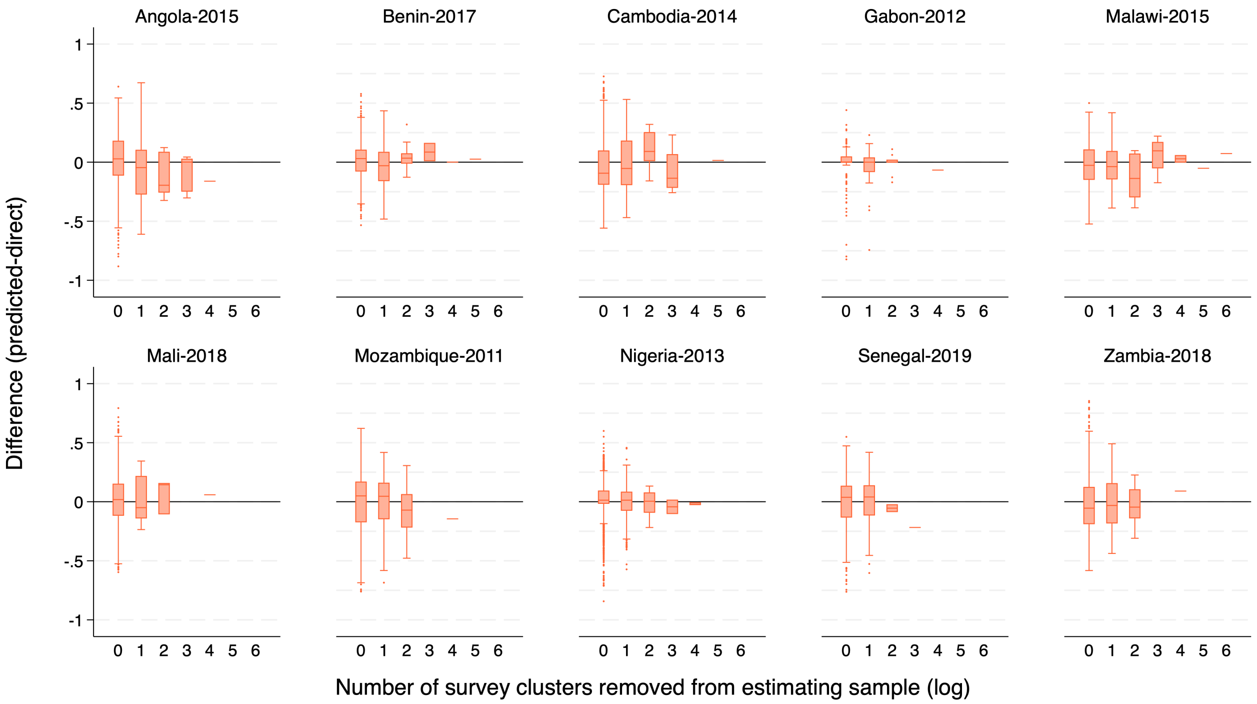


**Supplementary Figure 25.** Difference between LIDW-based Out-Of-Sample *predictions* and DHS *direct* estimates) for the indicator Exclusive Breastfeeding (0-6 months) across the 10 countries by number of *survey clusters* excluded from the estimation sample (log).

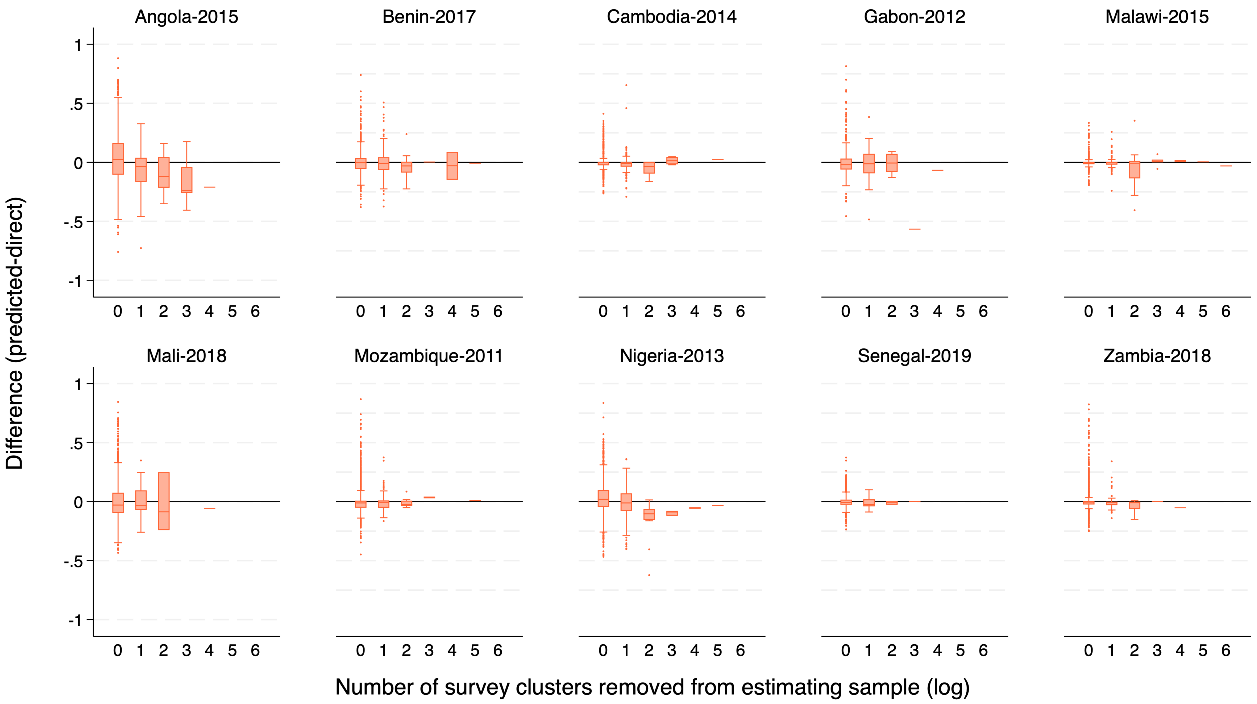


**Supplementary Figure 26.** Difference between LIDW-based Out-Of-Sample *predictions* and DHS *direct* estimates) for the indicator BCG Immunization (12-23 months) across the 10 countries by number of *survey clusters* excluded from the estimation sample (log).

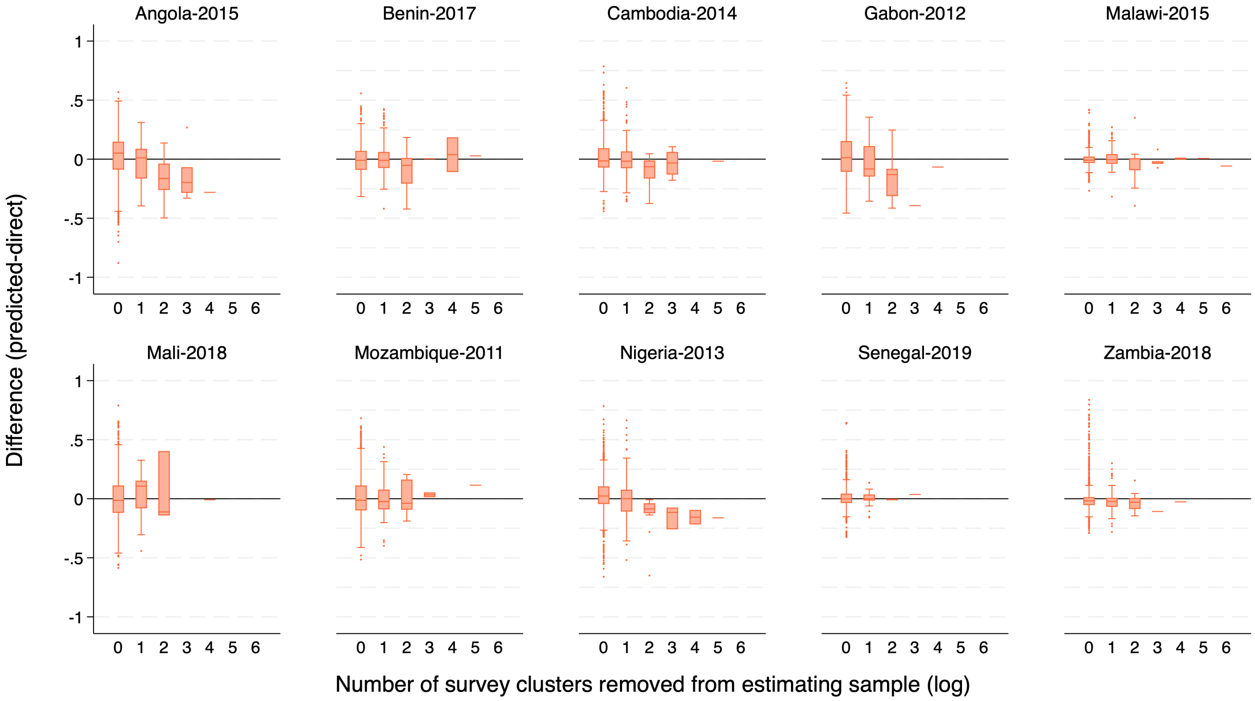


**Supplementary Figure 27.** Difference between LIDW-based Out-Of-Sample *predictions* and DHS *direct* estimates) for the indicator DPT3 Immunization (12-23 months) across the 10 countries by number of *survey clusters* excluded from the estimation sample (log).

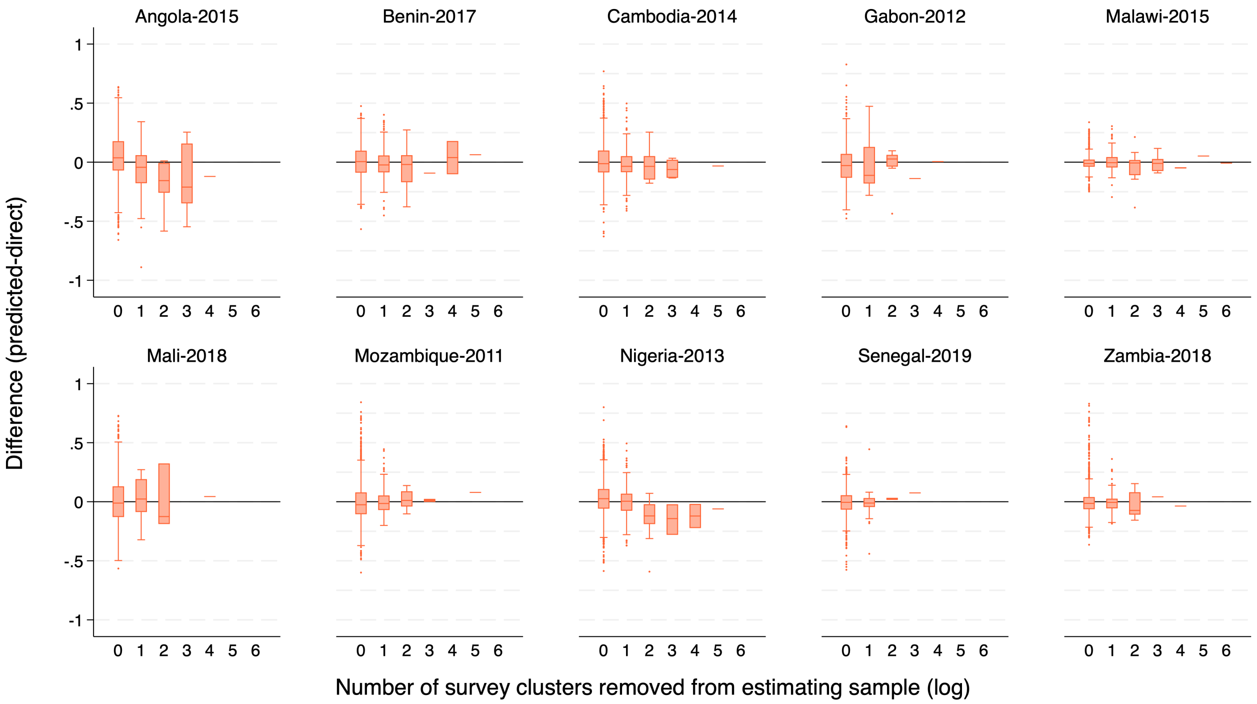


**Supplementary Figure 28.** Difference between LIDW-based Out-Of-Sample *predictions* and DHS *direct* estimates) for the indicator Measles Immunization (12-23 months) across the 10 countries by number of *survey clusters* excluded from the estimation sample (log).

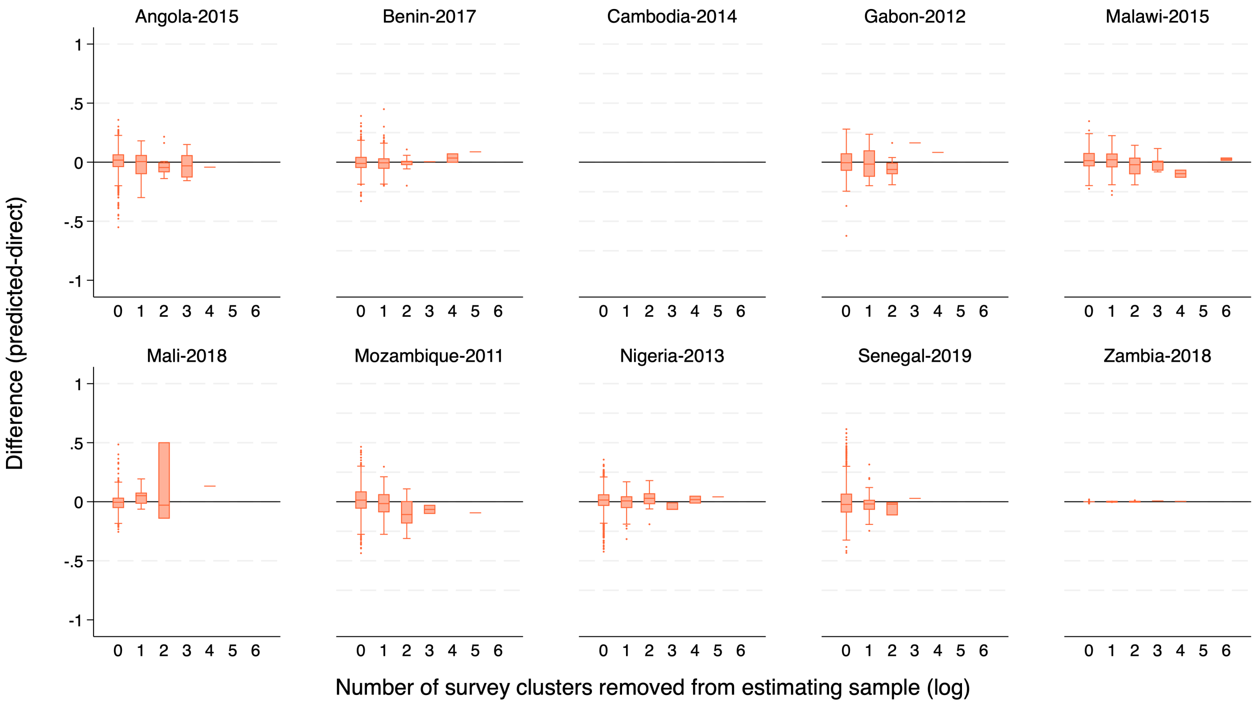


**Supplementary Figure 29.** Difference between LIDW-based Out-Of-Sample *predictions* and DHS *direct* estimates) for the indicator Use of Mosquito Nets among Children (0-59 months) across the 10 countries by number of *survey clusters* excluded from the estimation sample (log).

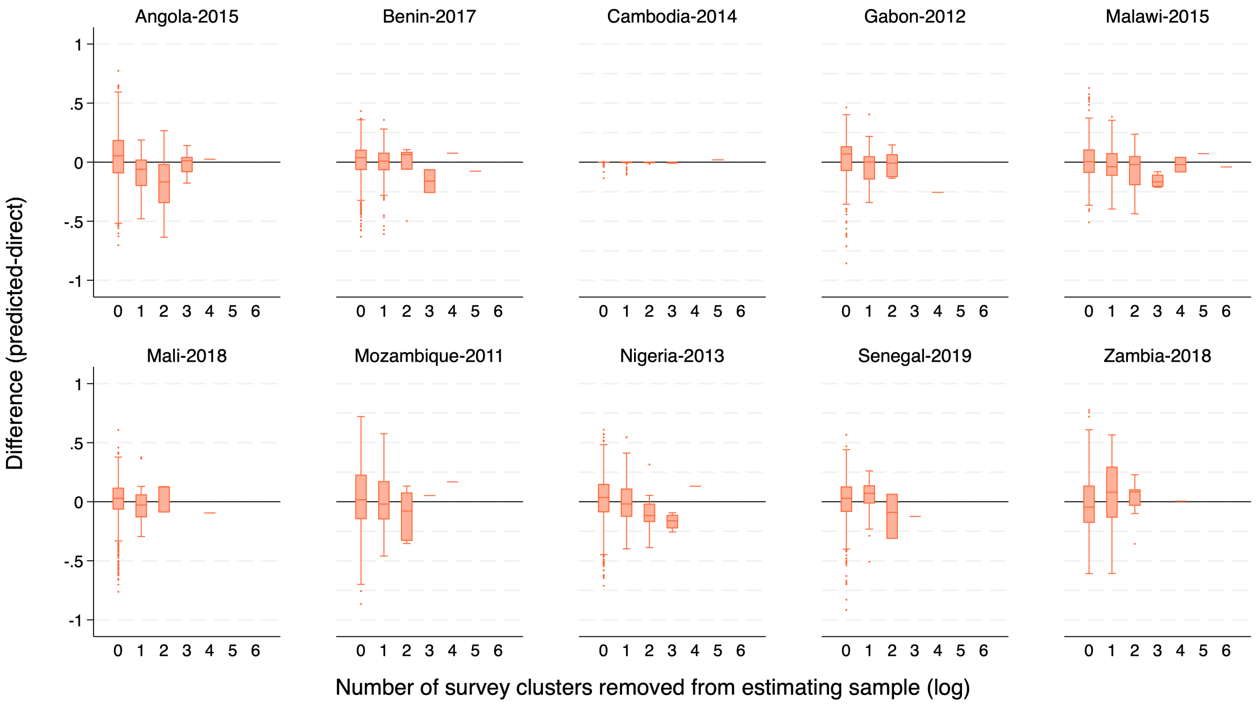


**Supplementary Figure 30.** Difference between LIDW-based Out-Of-Sample *predictions* and DHS *direct* estimates) for the indicator Diarrhea Treatment with ORS among children (0-59 months) across the 10 countries by number of *survey clusters* excluded from the estimation sample (log).

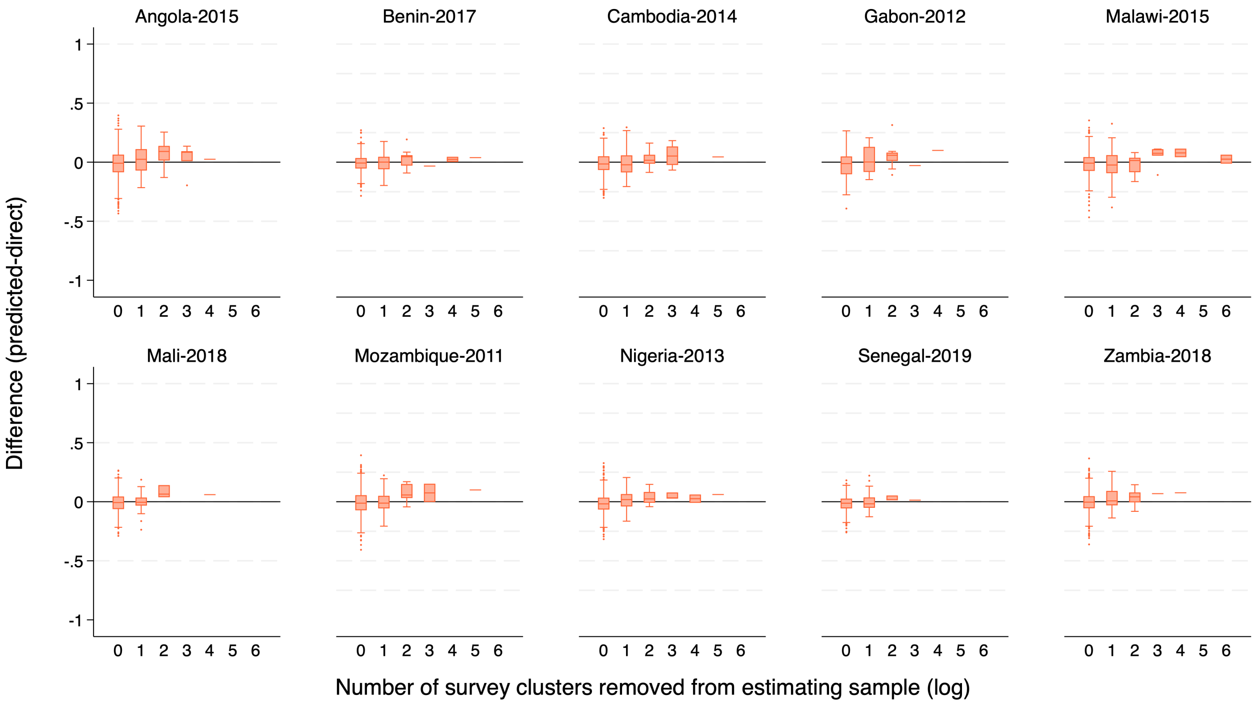


**Supplementary Figure 31.** Difference between LIDW-based Out-Of-Sample *predictions* and DHS *direct* estimates) for the indicator Stunting Prevalence among children (0-59 months) across the 10 countries by number of *survey clusters* excluded from the estimation sample (log).

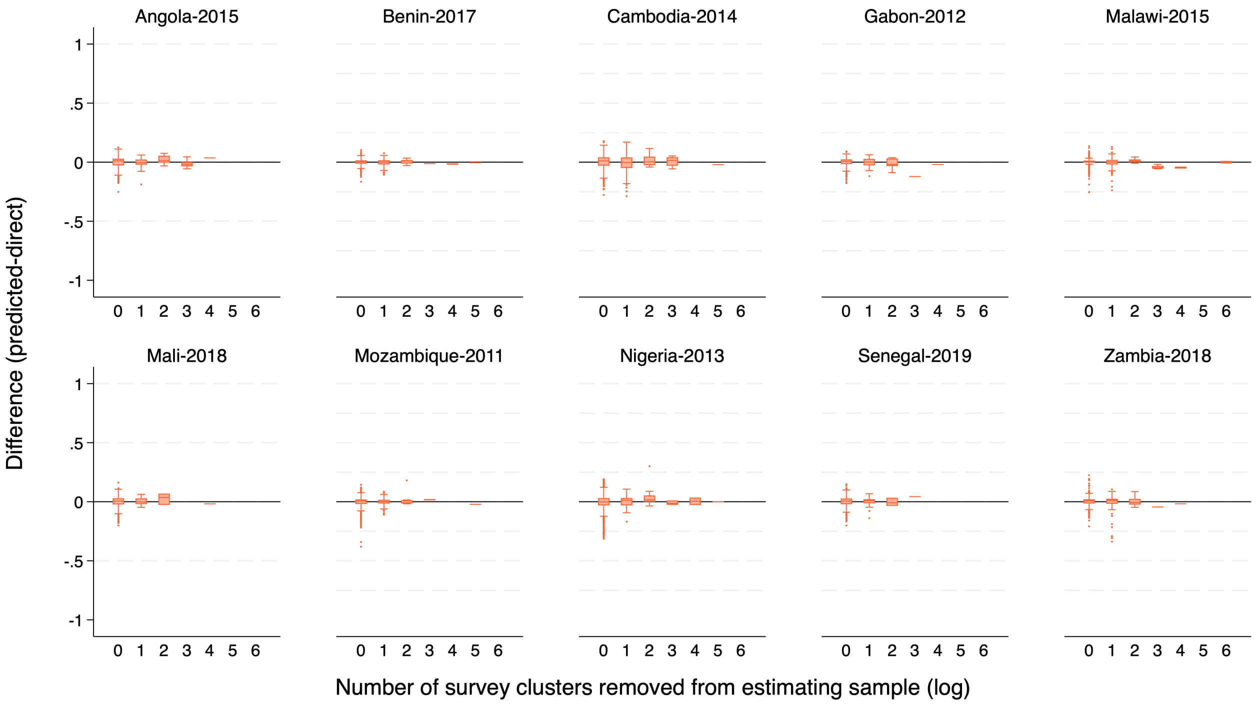


**Supplementary Figure 32.** Difference between LIDW-based Out-Of-Sample *predictions* and DHS *direct* estimates) for the indicator Wasting Prevalence among children (0-59 months) across the 10 countries by number of *survey clusters* excluded from the estimation sample (log).

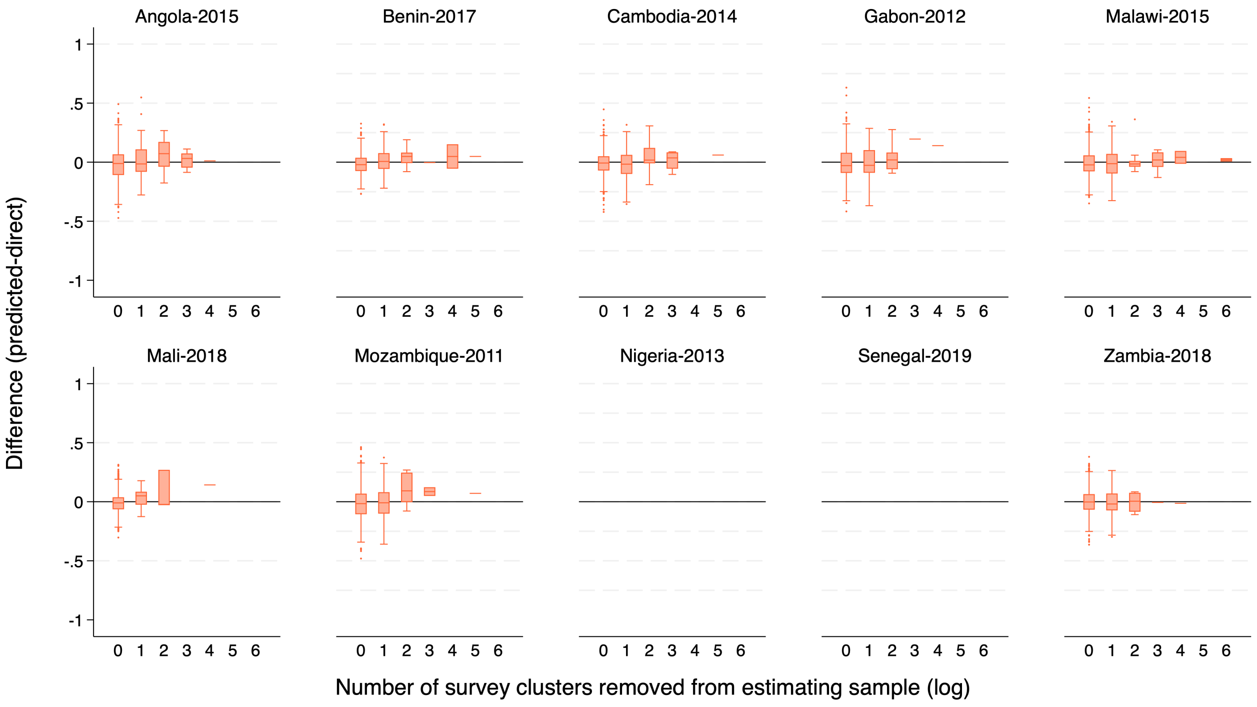


**Supplementary Figure 33.** Difference between LIDW-based Out-Of-Sample *predictions* and DHS *direct* estimates) for the indicator Prevalence of Anemia in Children (6-59 months) across the 10 countries by number of *survey clusters* excluded from the estimation sample (log).

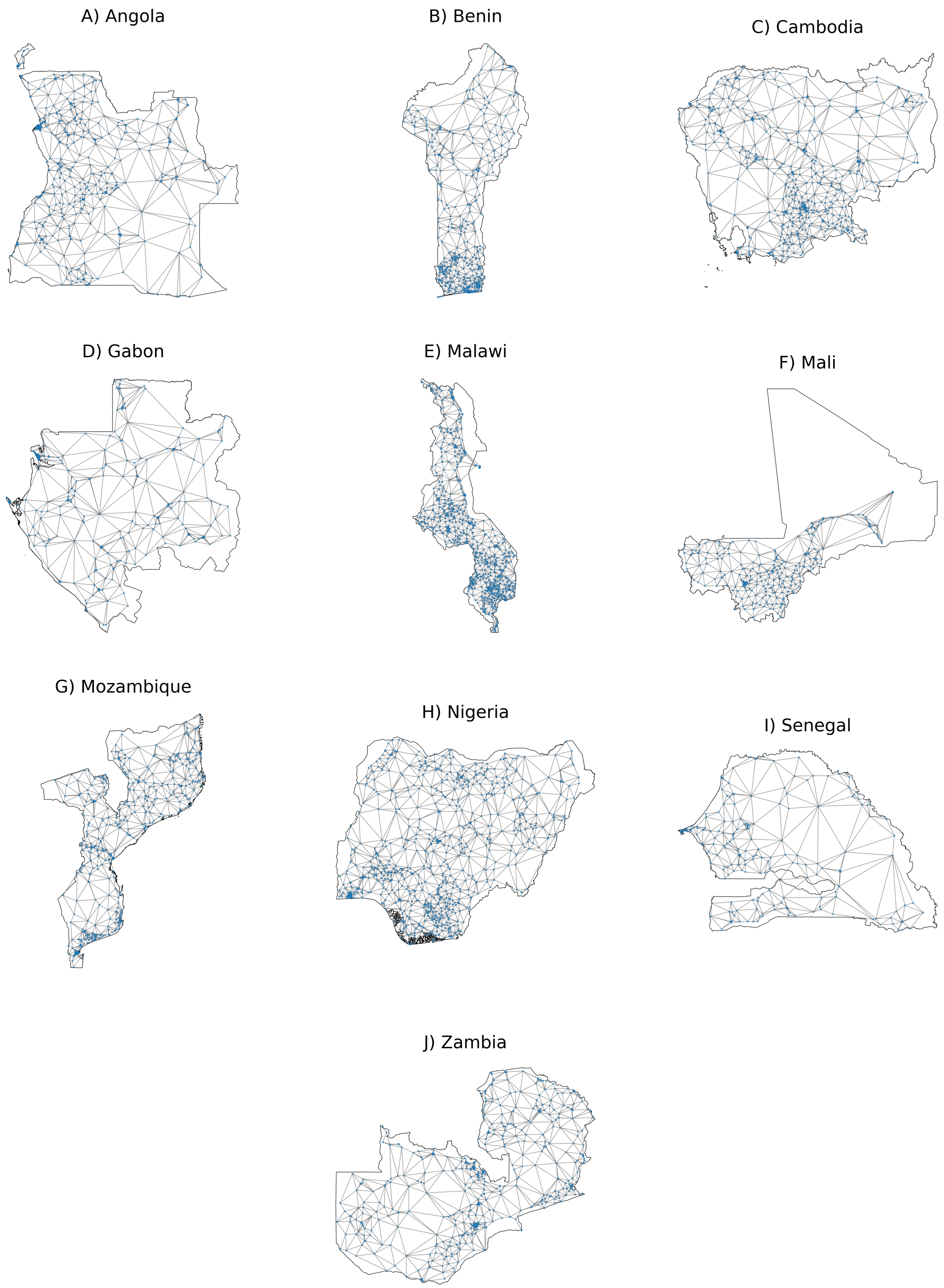


**Supplementary Figure 34.** DHS cluster network used for *indirect* estimates. Map scales are not uniform for illustrative purposes.

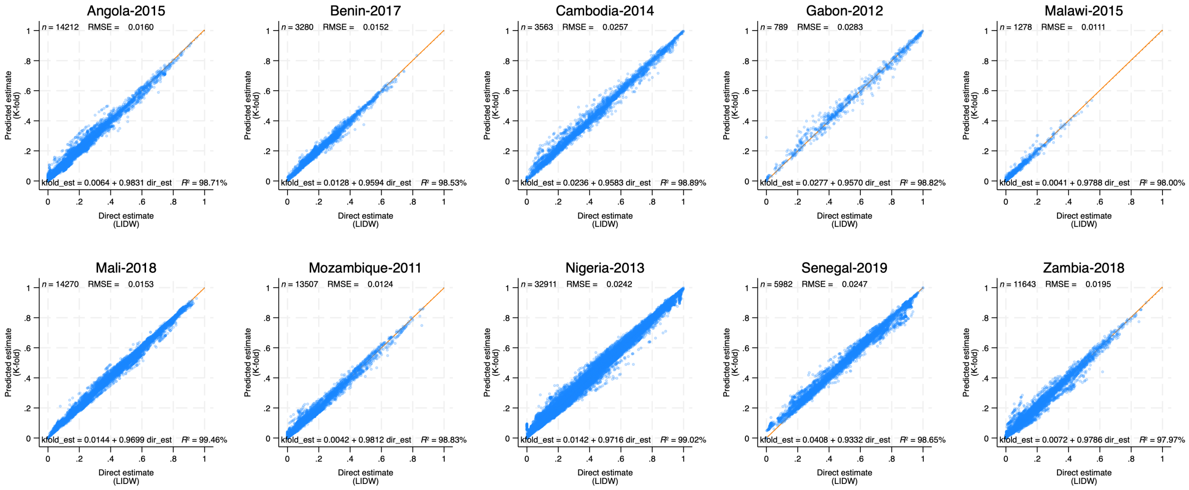


**Supplementary Figure 35.** K-fold validation results; comparison of *predicted* and *direct* estimates within each settlement. Each circle corresponds to one settlement. The solid line corresponds to the linear prediction. The corresponding equation is shown within each plot in addition to the number of settlements included in the validation, the RMSE estimate, and the $R^2$. Results for indicator Households with access to Electricity.

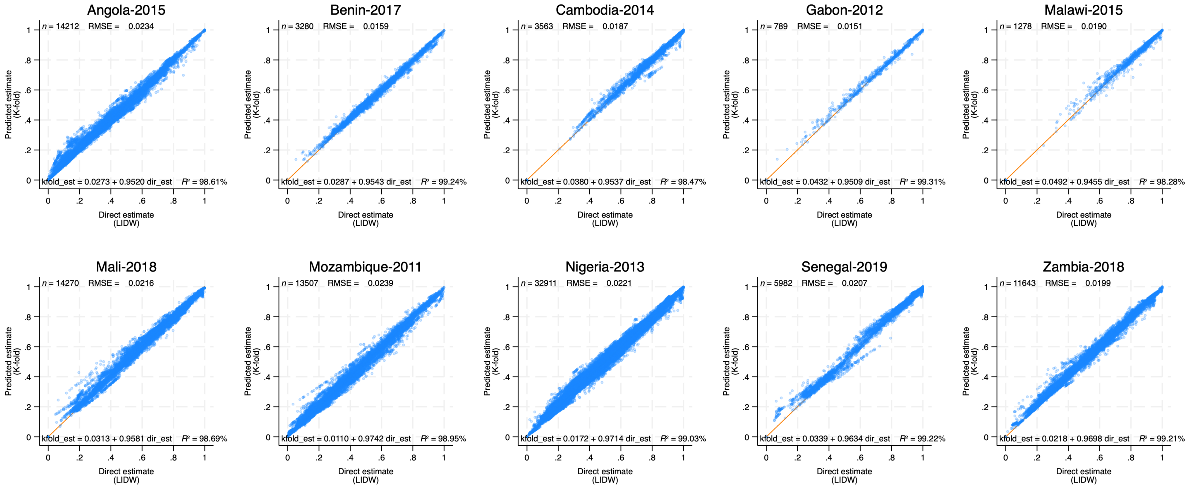


**Supplementary Figure 36.** K-fold validation results; comparison of *predicted* and *direct* estimates within each settlement. Each circle corresponds to one settlement. The solid line corresponds to the linear prediction. The corresponding equation is shown within each plot in addition to the number of settlements included in the validation, the RMSE estimate, and the $R^2$. Results for indicator Access to Improved Water Source.

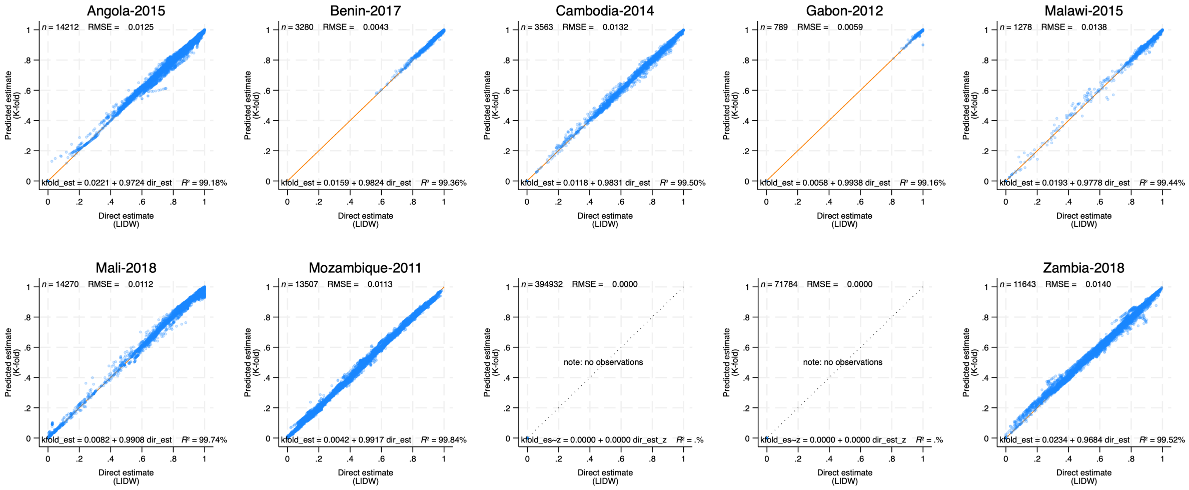


**Supplementary Figure 37.** K-fold validation results; comparison of *predicted* and *direct* estimates within each settlement. Each circle corresponds to one settlement. The solid line corresponds to the linear prediction. The corresponding equation is shown within each plot in addition to the number of settlements included in the validation, the RMSE estimate, and the $R^2$. Results for indicator Iodized Salt Intake in Household.

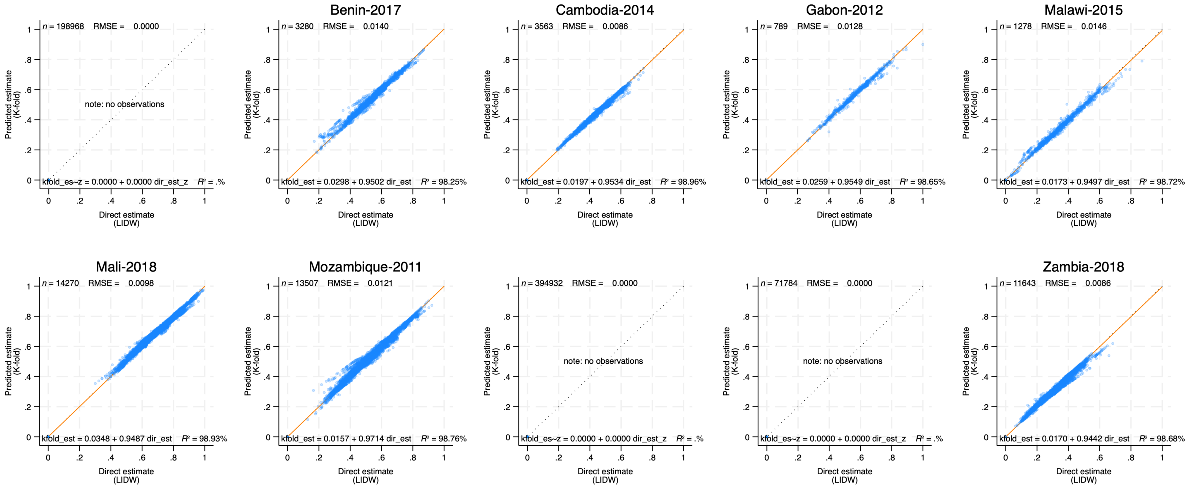


**Supplementary Figure 38.** K-fold validation results; comparison of *predicted* and *direct* estimates within each settlement. Each circle corresponds to one settlement. The solid line corresponds to the linear prediction. The corresponding equation is shown within each plot in addition to the number of settlements included in the validation, the RMSE estimate, and the $R^2$. Results for indicator Percentage of women with anemia.

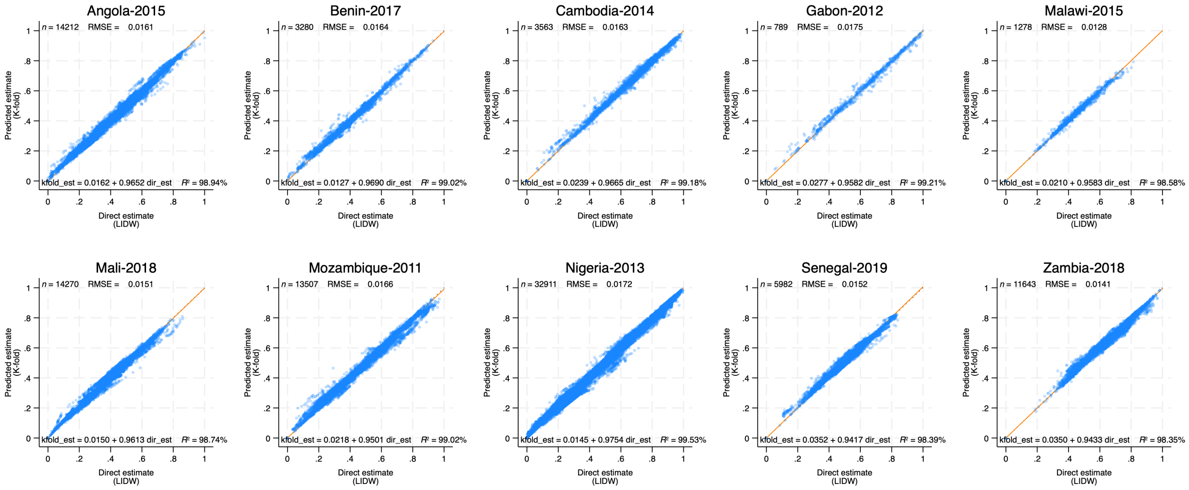


**Supplementary Figure 39.** K-fold validation results; comparison of *predicted* and *direct* estimates within each settlement. Each circle corresponds to one settlement. The solid line corresponds to the linear prediction. The corresponding equation is shown within each plot in addition to the number of settlements included in the validation, the RMSE estimate, and the $R^2$. Results for indicator Antenatal Care Visits (4+) during Pregnancy.

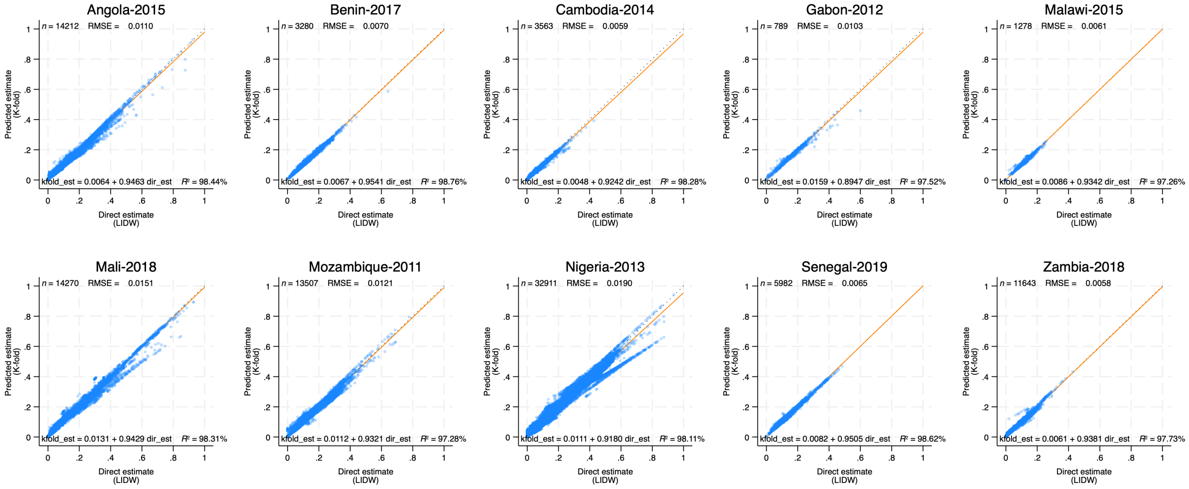


**Supplementary Figure 40.** K-fold validation results; comparison of *predicted* and *direct* estimates within each settlement. Each circle corresponds to one settlement. The solid line corresponds to the linear prediction. The corresponding equation is shown within each plot in addition to the number of settlements included in the validation, the RMSE estimate, and the $R^2$. Results for indicator Low Birth Weight Prevalence.

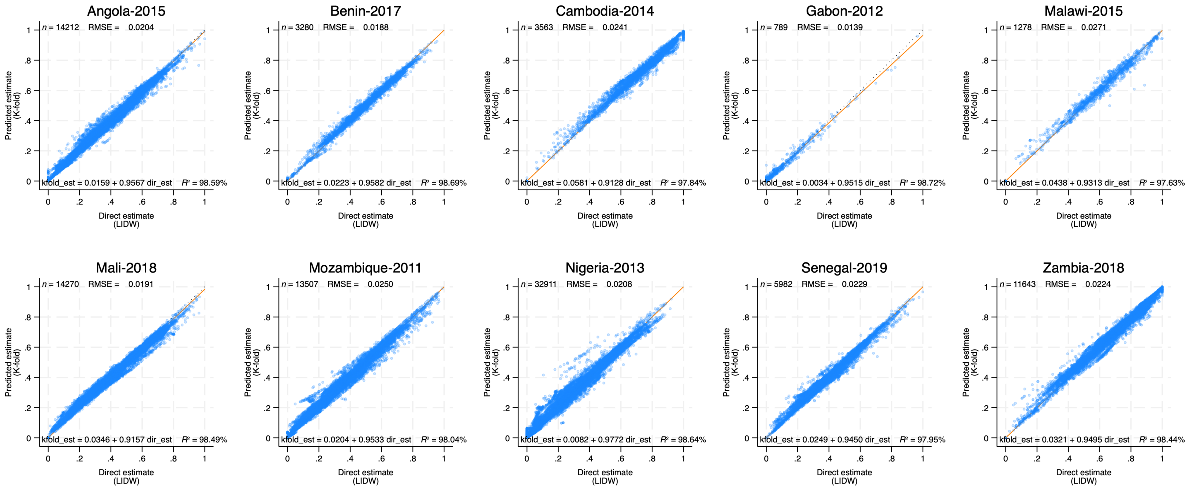


**Supplementary Figure 41.** K-fold validation results; comparison of *predicted* and *direct* estimates within each settlement. Each circle corresponds to one settlement. The solid line corresponds to the linear prediction. The corresponding equation is shown within each plot in addition to the number of settlements included in the validation, the RMSE estimate, and the $R^2$. Results for indicator Exclusive Breastfeeding (0-6 months).

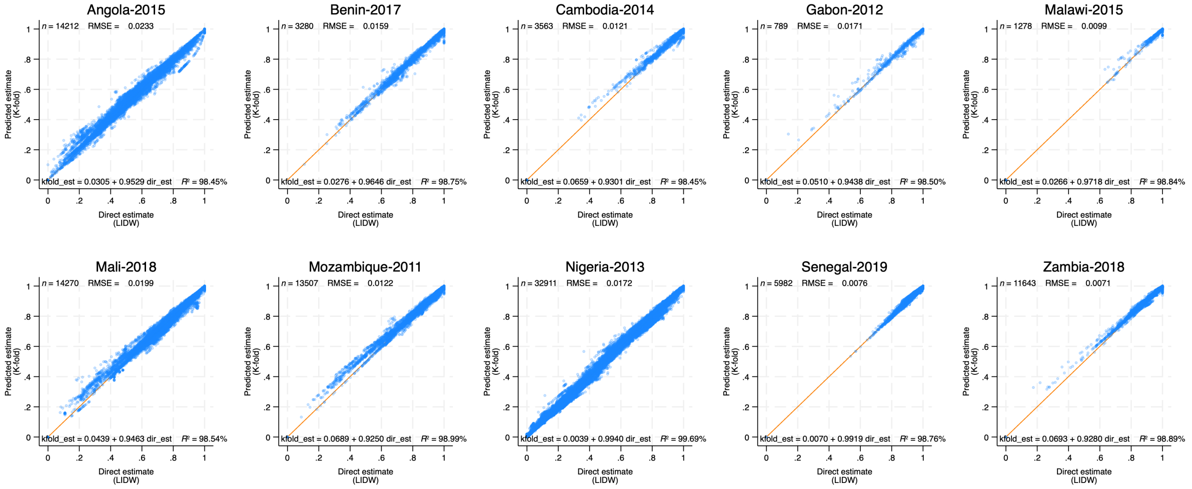


**Supplementary Figure 42.** K-fold validation results; comparison of *predicted* and *direct* estimates within each settlement. Each circle corresponds to one settlement. The solid line corresponds to the linear prediction. The corresponding equation is shown within each plot in addition to the number of settlements included in the validation, the RMSE estimate, and the $R^2$. Results for indicator BCG Immunization (12-23 months).

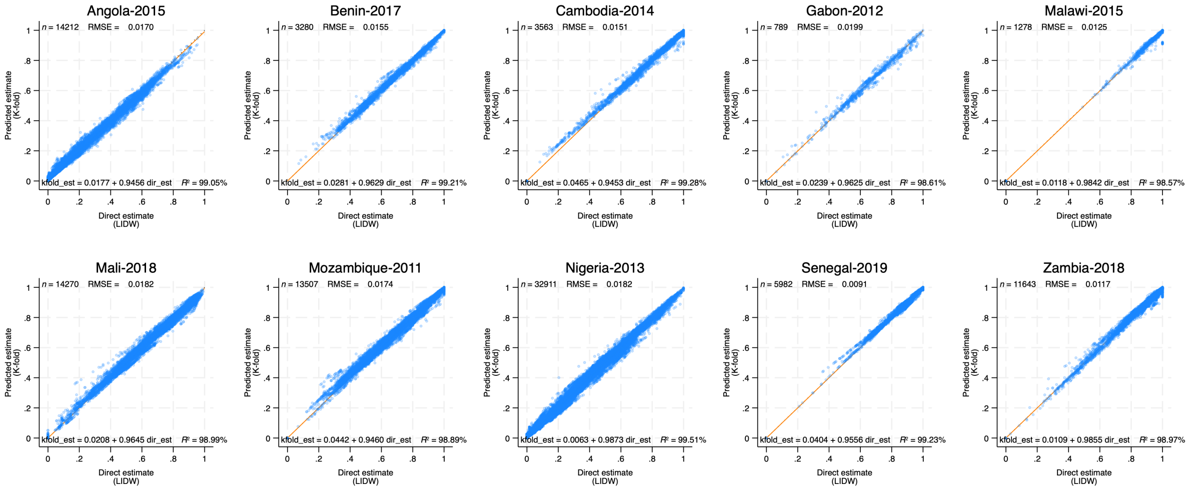


**Supplementary Figure 43.** K-fold validation results; comparison of *predicted* and *direct* estimates within each settlement. Each circle corresponds to one settlement. The solid line corresponds to the linear prediction. The corresponding equation is shown within each plot in addition to the number of settlements included in the validation, the RMSE estimate, and the $R^2$. Results for indicator DPT3 Immunization (12-23 months).

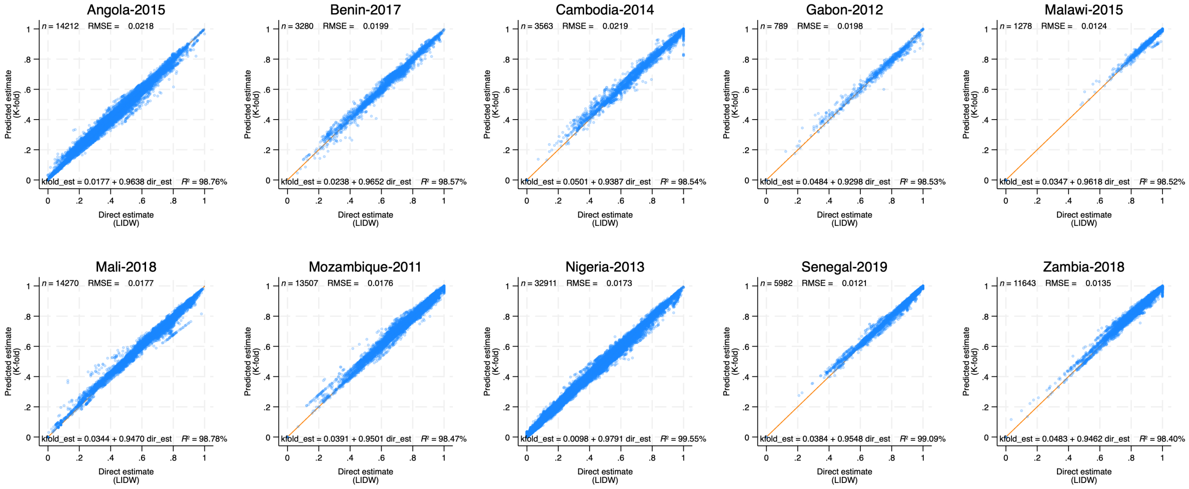


**Supplementary Figure 44.** K-fold validation results; comparison of *predicted* and *direct* estimates within each settlement. Each circle corresponds to one settlement. The solid line corresponds to the linear prediction. The corresponding equation is shown within each plot in addition to the number of settlements included in the validation, the RMSE estimate, and the $R^2$. Results for indicator Measles Immunization (12-23 months).

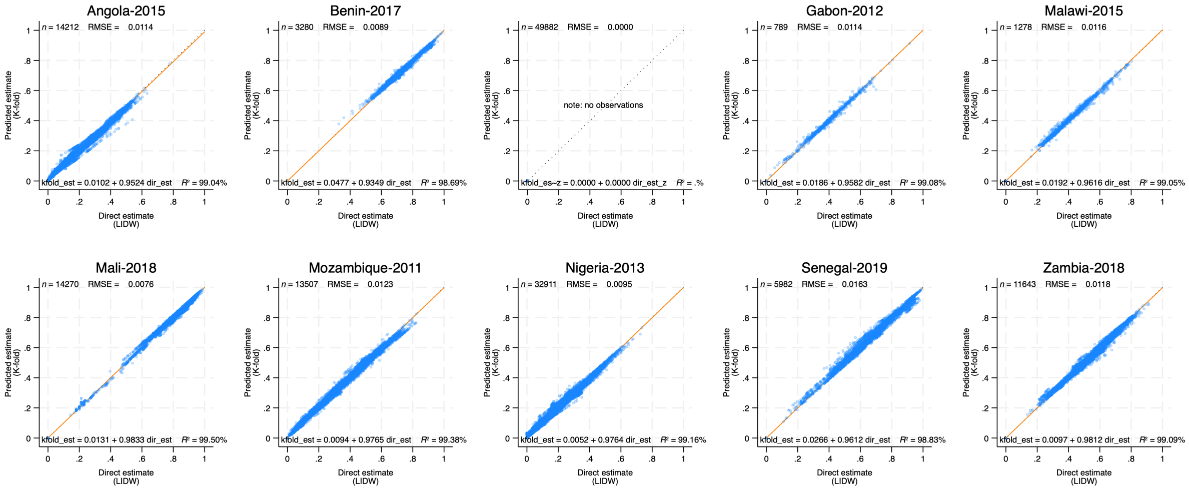


**Supplementary Figure 45.** K-fold validation results; comparison of *predicted* and *direct* estimates within each settlement. Each circle corresponds to one settlement. The solid line corresponds to the linear prediction. The corresponding equation is shown within each plot in addition to the number of settlements included in the validation, the RMSE estimate, and the $R^2$. Results for indicator Children (0-59 months) Slept under Mosquito Net.

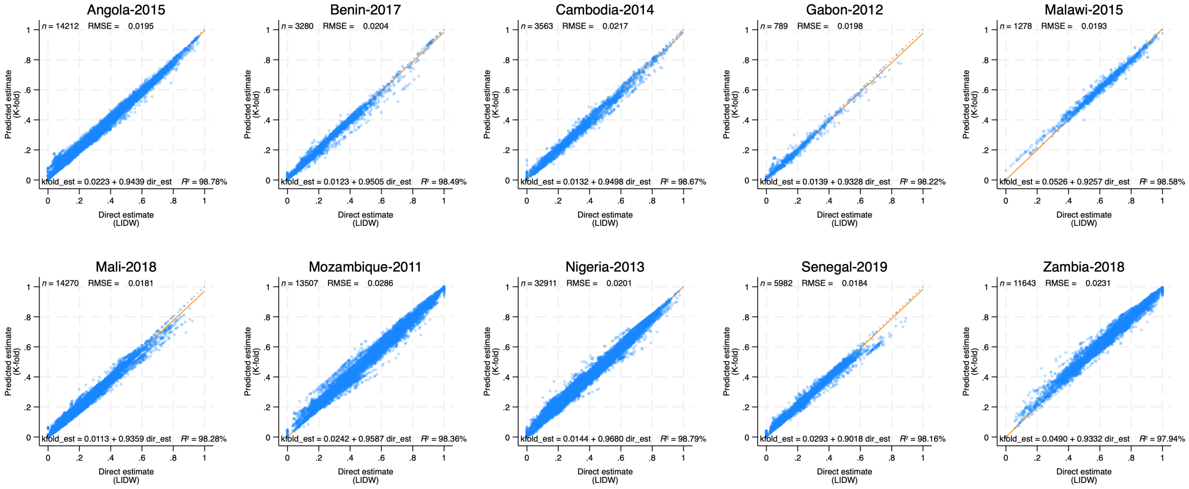


**Supplementary Figure 46.** K-fold validation results; comparison of *predicted* and *direct* estimates within each settlement. Each circle corresponds to one settlement. The solid line corresponds to the linear prediction. The corresponding equation is shown within each plot in addition to the number of settlements included in the validation, the RMSE estimate, and the $R^2$. Results for indicator Diarrhea Treatment with ORS (0-59 months).

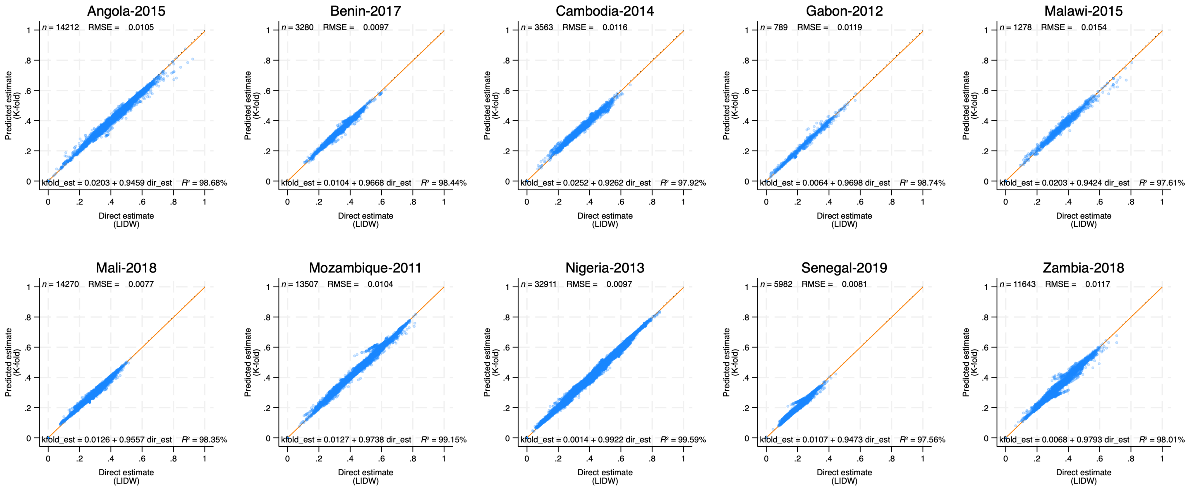


**Supplementary Figure 47.** K-fold validation results; comparison of *predicted* and *direct* estimates within each settlement. Each circle corresponds to one settlement. The solid line corresponds to the linear prediction. The corresponding equation is shown within each plot in addition to the number of settlements included in the validation, the RMSE estimate, and the $R^2$. Results for indicator Stunting Prevalence (0-59 months).

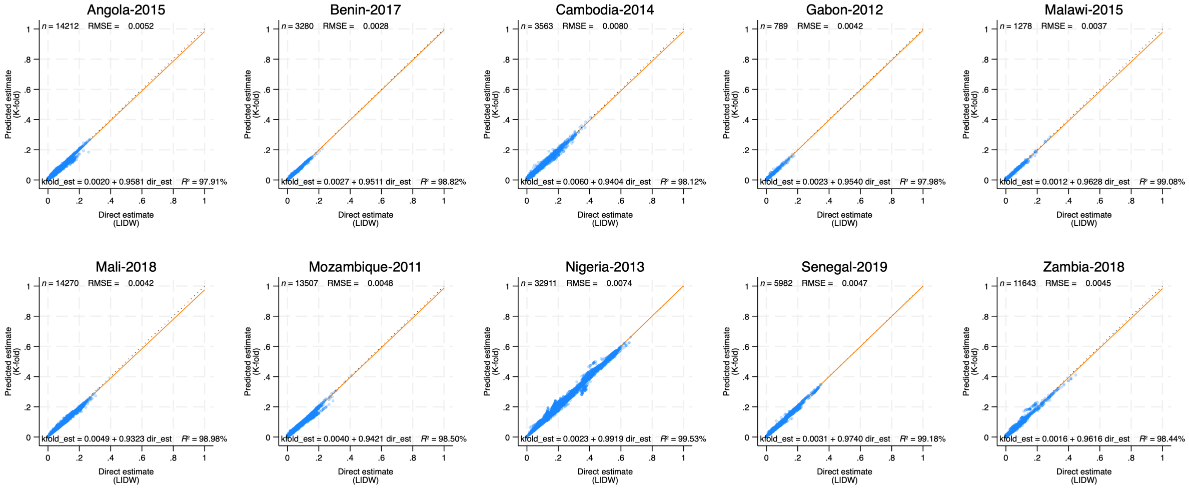


**Supplementary Figure 48.** K-fold validation results; comparison of *predicted* and *direct* estimates within each settlement. Each circle corresponds to one settlement. The solid line corresponds to the linear prediction. The corresponding equation is shown within each plot in addition to the number of settlements included in the validation, the RMSE estimate, and the $R^2$. Results for indicator Wasting Prevalence (0-59 months).

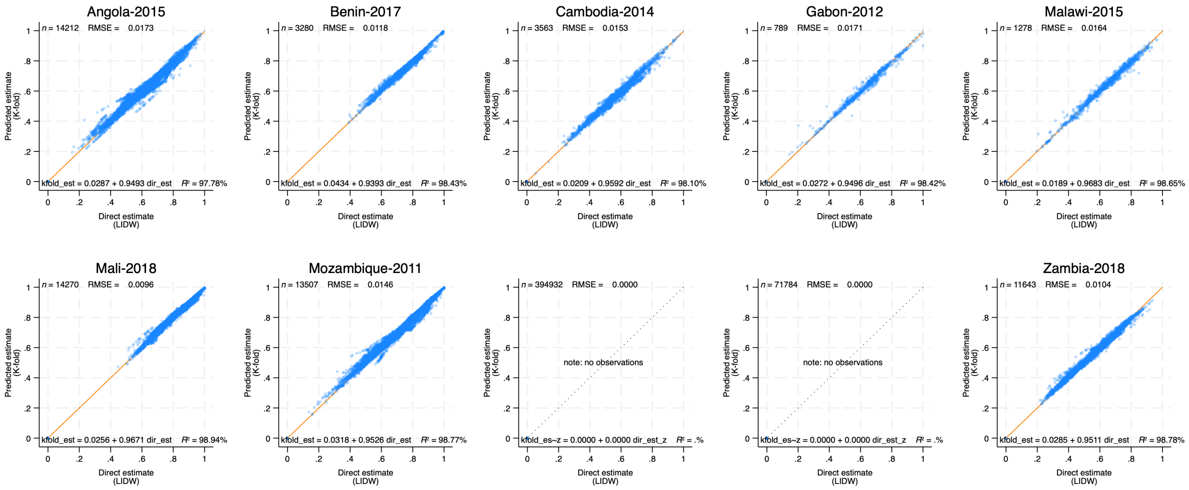


**Supplementary Figure 49.** K-fold validation results; comparison of *predicted* and *direct* estimates within each settlement. Each circle corresponds to one settlement. The solid line corresponds to the linear prediction. The corresponding equation is shown within each plot in addition to the number of settlements included in the validation, the RMSE estimate, and the $R^2$. Results for indicator Prevalence of any Anemia in Children (0-59 months).
